# A multivariate meta-analysis on the relationship between social connectedness and pain

**DOI:** 10.64898/2026.03.24.26349176

**Authors:** Aleksandra Piejka, Sigrid Elsenbruch, Adriane Icenhour, Łukasz Okruszek, Dirk Scheele, Julian Packheiser

## Abstract

**Background:** Social disconnection has emerged as a major public health concern, with health risks comparable to established biomedical factors. At the same time, pain remains the leading cause of years lived with disability worldwide, imposing profound individual and societal costs. Although social factors are increasingly implicated in pain perception and chronification, existing evidence is fragmented across heterogeneous and often conflated constructs of social connectedness. It remains unclear whether subjective experiences such as loneliness or structural factors such as social isolation differentially relate to pain outcomes. A comprehensive synthesis directly comparing these dimensions has been lacking.

**Methods:** We conducted a preregistered multivariate meta-analysis (PROSPERO: CRD420250643896) including 239 studies, 520 effect sizes, and 1,407,803 participants from clinical and non-clinical populations. Pain outcomes encompassed sensory, affective, cognitive, and functional domains. Social connectedness was operationalized as loneliness, social isolation, social support, and social exclusion. Multilevel random-effects models accounted for within-study dependency, with extensive sensitivity analyses and correction for small-study bias.

**Results:** Across populations and social outcomes, greater social connectedness was associated with lower pain (z = −0.09, 95% CI −0.11 to −0.07). Notably, loneliness emerged as the strongest correlate (z = 0.14, 95% CI 0.11 to 0.17). Associations with social isolation were smaller compared to loneliness but were also significant (z = 0.09, 95% CI 0.05 to 0.13). Social support showed modest, significant inverse associations (z = −0.05, 95% CI −0.08 to −0.03), primarily confined to affective and somatic pain components. No reliable association was observed for social exclusion. Associations were consistent across age, sex, and clinical status, and longitudinal evidence supported temporal links between changes in social connectedness and subsequent pain outcomes.

**Conclusions:** This large-scale synthesis identifies subjective social disconnection, particularly loneliness, as a robust correlate of pain across populations and pain dimensions, exceeding the relevance of objective social isolation. Given evidence linking loneliness to increased analgesic and psychotropic medication use, social disconnection may contribute to pharmacological burden and polypharmacy risk in vulnerable populations. Social connectedness emerges as a clinically meaningful, non-pharmacological determinant of pain and a potential target for integrative pain prevention and management strategies.

## Introduction

At a time when social disconnection is rising globally, understanding the health consequences of social ties has become a pressing public health priority. A substantial body of population-based and experimental research has established social connectedness as a central determinant of health. Encompassing both the structural availability of social relationships and the perceived adequacy and reliability of social ties, social connectedness has been linked to morbidity and mortality with effect sizes comparable to major biomedical risk factors [1,47]. Deficits in social connectedness are associated with increased cardiovascular risk, cognitive decline, heightened inflammatory activity, dysregulated stress responses, and vulnerability to mental disorders [2–6]. Increasing recognition of social disconnection as a determinant of health has led national and international bodies to designate it as a priority for research and intervention [7].

Crucially, social connectedness is not a unitary construct. What appears as a single social dimension in public discourse in fact encompasses conceptually and biologically distinct phenomena. Objective social isolation captures the structural thinning of social networks comprising reduced network size and diminished interaction frequency [8]. Subjective dimensions, by contrast, reflect how individuals evaluate their social world. Loneliness denotes the distressing discrepancy between desired and actual relationships [9], whereas perceived social support refers to the belief that emotional, instrumental, or practical assistance is available when needed [10]. Some constructs, such as social support, straddle subjective appraisal and objective receipt of help [11]. Social exclusion, that is the experiences of ostracism or rejection, adds further complexity as it overlaps with but is not reducible to loneliness or isolation and resists clear classification as purely structural or subjective [12,13]. These facets are only moderately correlated [14] and increasingly appear to engage partially distinct biological systems, including inflammatory, neuroendocrine, and neural pathways [15]. Collapsing them into a single construct therefore risks masking meaningful differences in how specific forms of social experience relate to public health.

Pain constitutes one of the most prevalent and debilitating symptoms worldwide, affecting individuals across diagnoses and clinical populations, and is a leading cause of suffering and disability. Epidemiological estimates indicate that 20-30% of adults globally experience some form of pain, with approximately 10% developing chronic pain conditions each year [16,17]. Chronic pain is closely intertwined with impaired mental well-being, reduced social functioning, and diminished occupational capacity [18]. At the societal level, these individual consequences translate into substantial economic costs, amounting to billions of euros annually due to health-care utilization, loss of productivity, and disability benefits [19]. This pervasive burden makes pain a critical focus of biomedical and public health research, independent of its specific etiology.

Contemporary models of pain increasingly conceptualize pain as a multidimensional and context-sensitive experience that emerges from dynamic interactions between biological processes, psychological states, and social environments. Frameworks such as the biopsychosocial model, social communication models of pain, and predictive processing accounts emphasize that pain is not solely a sensory signal but also a socially embedded experience shaped by interpersonal contexts, expectations, and perceived support [20–22]. Within these models, social factors such as social connection or disconnection are understood to influence both acute pain responses, longer-term trajectories of pain persistence, and chronification through behavioral, affective, and neurobiological pathways.

Clinical observations further underscore the relevance of social factors for pain management. Population-based data from older adults show that loneliness is associated with increased use of analgesic and psychotropic medications, including non-opioid pain medication, even after adjustment for age, comorbidity, and functional status [23]. This pattern suggests that socially disconnected individuals may rely more heavily on pharmacological strategies to manage pain and its psychological sequelae, increasing the risk of polypharmacy and medication-related harm in vulnerable populations. At the mechanistic level, converging evidence indicates that social and physical pain share common representational and modulatory components and engage partially overlapping neural systems, particularly within affective pain-processing circuits [24]. Experimental studies show that social rejection activates regions such as the anterior cingulate cortex and anterior insula [25], which are also implicated in the affective dimension of physical pain. In parallel, neurobiological research demonstrates that central oxytocinergic pathways regulate social affiliation, stress responsivity, and affective processing, providing a plausible biological interface through which social connectedness may shape pain experience [26–28]. Together, these findings support a functional overlap between social and physical pain processes and highlight that social experiences may operate as core modulators of pain processing, with direct implications for clinical management and health outcomes.

Despite growing interest in the social modulation of pain, existing research remains fragmented. Most studies have focused on the presumed protective effects of social support, with many reporting inverse associations between support and pain severity in clinical samples [29,30]. However, it remains unclear whether the link between pain and social connectedness is specific for patient populations, or whether it can be similarly observed in healthy samples. Moreover, effect sizes vary widely, and prior meta-analytic work on experimentally induced pain has yielded mixed results depending on the type of support and the relationship to the support provider [31]. In contrast, loneliness and social isolation have not been systematically synthesized in relation to pain despite being well-established risk factors for adverse health outcomes. Although large-scale observational studies increasingly report positive associations between loneliness, isolation, and pain outcomes [32], individual studies are susceptible to bias and effect size inflation [33]. Evidence regarding social exclusion is particularly inconsistent, with laboratory studies reporting both pain sensitization and pain numbing effects [25,34].

This heterogeneity has resulted in a substantial epistemic gap. To date, no meta-analytic synthesis has directly compared the magnitude and robustness of associations between different dimensions of social connectedness and pain outcomes, nor systematically examined key moderators such as pain type, chronicity, population characteristics, or study design. This lack of integration hampers theory development and may contribute to poorly targeted clinical interventions as evidence from experimental and longitudinal research that can provide causal insight into the relationship between social connectedness and pain remains sparse in this field.

The present preregistered multivariate meta-analysis (PROSPERO: CRD420250643896) addresses these limitations by quantitatively synthesizing evidence on the association between pain and multiple dimensions of social connectedness, including loneliness, social isolation, social support, and social exclusion. Specifically, we aimed to (1) estimate the overall association between social connectedness and subjective pain across clinical and non-clinical populations; (2) compare effect sizes across distinct social dimensions; and (3) examine moderation by pain characteristics, demographic factors, study design, and measurement properties. We hypothesized that social connectedness would be significantly associated with pain outcomes. Specifically, we expected loneliness and social isolation to be positively associated with pain, whereas social support would show an inverse association. Given inconsistent prior findings, no directional hypothesis was specified for social exclusion. We further anticipated substantial heterogeneity across studies, reflecting variation in populations, pain characteristics, and methodological approaches.

## Methods

### Search strategy

We conducted a pre-registered systematic literature search in PubMed, Web of Science, and Google Scholar (first 70 pages sorted by relevance) using the following terms: *loneliness* OR *social isolation* OR *social exclusion* OR *ostracism* OR *social withdrawal* OR *social support* OR *social connectedness* OR *social relationship* AND *pain*. Reference lists of relevant review articles were screened to identify additional eligible studies. No restrictions were applied with respect to publication date. Searches were conducted between February 2025 and August 2025. Articles published in English, German, or Polish were eligible; all included studies were published in English.

### Study Selection

#### Eligibility criteria

Studies were eligible for inclusion if they met all of the following criteria:

1. Reported a pain outcome, defined as (a) a self-reported pain measure or (b) inclusion of a sample with a diagnosed pain condition (e.g., chronic low back pain). For clinically diagnosed samples, we included cohorts irrespective of the underlying pain etiology (e.g., primary or secondary pain).
2. Included a measure of social connectedness, operationalized as loneliness, social isolation, social support, or social exclusion.
3. For experimental studies, included an appropriate control condition (e.g., presence vs absence of social support, or clinical vs healthy cohort comparison).

All included studies are listed in Supplementary Table 1. Excluded studies and reasons for exclusion are provided in Supplementary Table 2. Using this open search strategy, eligible studies published between 1976 and 2025 were identified. The flowchart for the search of existing meta-analyses is depicted in Figure 1. Deviations from the pre-registered inclusion criteria can be found under *Open Science practices*.

**Figure 1.**
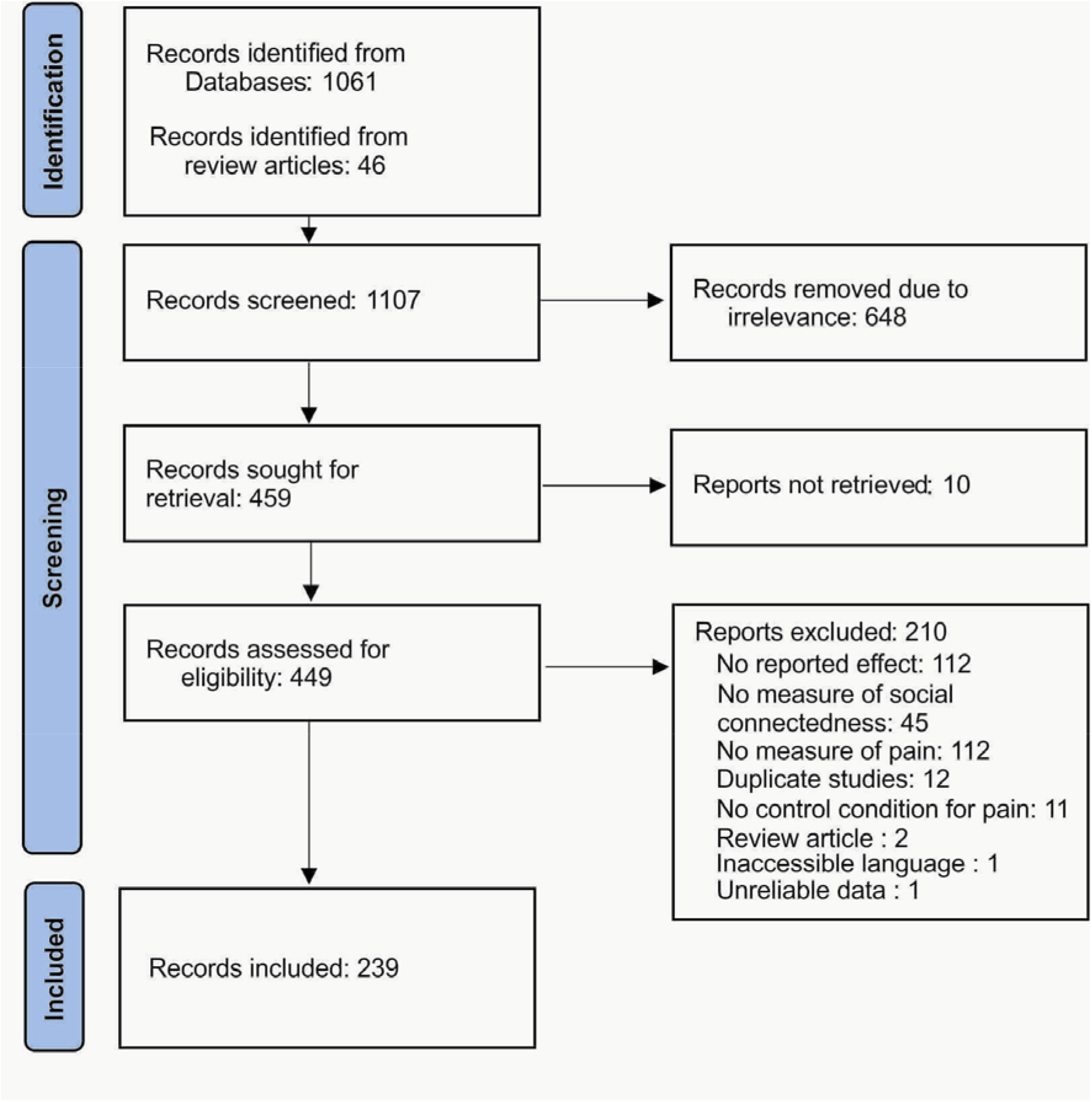
PRISMA 2020 flowchart detailing the identification and screening of identified records for the systematic review and meta-analysis.

### Data extraction and pre-processing

Data extraction was conducted between February and October 2025 by two independent reviewers (AP and JP) using separate extraction files. Title and abstract screening and full-text eligibility assessment were conducted independently by AP and JP; any disagreements were resolved through discussion. Discrepancies in extracted data were resolved by consensus. Following title and abstract screening, full texts were assessed for eligibility.

Effect sizes were extracted or computed and converted to Fisher’s z, which is recommended for meta-analytical synthesis of correlational data [35], using the Campbell Collaboration Effect Size Calculator and the ESCAL platform. Fisher’s z was calculated as:

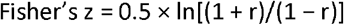

For interpretability, Fisher’s z can be approximated as Pearson’s *r* for values < 0.25 [36]. Odds ratios and Cohen’s d values were converted accordingly. In cases of thresholded statistics (e.g., *p* < .001), conservative estimates were used to avoid effect size inflation. For repeated-measures experimental designs, an assumed within-subject correlation of *r* = 0.5 was applied [37]. When multiple models were reported, the most fully adjusted model was selected.

Effect sizes were coded such that positive values reflected higher pain with higher social disconnectedness (e.g., loneliness), and negative values reflected lower pain with higher social connectedness (e.g., social support). Effect directions were reversed when necessary to ensure consistency. Study variance was derived from sample size. When effect sizes could not be extracted, corresponding authors were contacted.

Pre-registered moderators were extracted as follows:

1. **Social connectedness outcome** (loneliness, social isolation, social support, social exclusion).
2. **Population type** (clinical, healthy). Whenever a chronic pain diagnosis was explicitly mentioned or it was stated that pain was present for more than three months in the sample, the cohort was classified as clinical. Studies were excluded from the analysis if cohorts comprised both healthy individuals and individuals with a clinical diagnosis. Furthermore, exclusion occurred if there was uncertainty of the clinical status.
3. **Demographics** (mean age, sex/gender composition). If multiple cohorts were included in a single effect measure, we computed a weighted mean age across all individuals. The sex/gender ratio was computed by dividing the number of females by the total cohort sample size.
4. **Association type** (cross-sectional, longitudinal for observational studies; within-subject, between-subject for experimental studies). Observational studies were classified as cross-sectional if predictors and outcomes were measured at the same time point. Observational studies with multiple measurement time points were classified as longitudinal. Experimental studies with repeated-measures designs were classified as within-subject. Studies comparing different samples were classified as between-subject.
5. **Pain outcome type** (pain intensity, pain-related disability, pain unpleasantness, pain-related interference, pain frequency, pain duration, pain catastrophizing, pain tolerance, pain threshold). Composite measures (e.g., a questionnaire assessing both intensity and interference) were coded as a separate category and not further analyzed as part of this moderator.
6. **Outcome chronicity** (state vs trait). Outcomes were coded as state measures whenever they referred to an acute evaluation (e.g., pain in this very moment) or for outcomes measured within the last 24h. Measures beyond a 24h timeframe (e.g., painful experiences in the past two weeks) were coded as trait measures.
7. **Measurement type** (questionnaire vs single-item). Studies that did not use a questionnaire or a single item (e.g., a visual analogue scale) to determine social or pain outcomes (e.g., social support being represented through the presence of another human) were excluded from this moderator analysis.

Effect sizes were averaged when multiple assessments could not be meaningfully distinguished (e.g., repeated measurements or conceptually overlapping indicators).

### Statistical analysis and risk of bias assessment

Data were analyzed using R (v. 4.3.3 for Windows) and RStudio (2022.07.2 Build 576) [38] using rma.mv function from the metafor package [39] in a multivariate and multilevel fashion. In addition to our frequentist approach, Bayesian models were fitted using the brms package [40]. We used sigma = 0.1 as default prior (small-to-medium effect size prediction) for the fixed effects but computed models also with small effect size (sigma = 0.05) and medium effect size priors (sigma = 0.15). Effects in the manuscript are reported for sigma = 0.1, the results for the other priors can be found in the Supplementary Material as part of our sensitivity analyses (Supplementary Tables 3-16). In contrast to frequentist statistics, Bayesian statistics provide continuous evidence for either the null or alternative hypothesis. Bayes factors (BF) are reported as complementary continuous measures of evidence. Values of BF_10_ > 1 indicate support for the alternative hypothesis, whereas BF_10_ < 1 indicate support for the null hypothesis (e.g., BF_10_ = 2 or 0.5 reflects twice as much evidence for the alternative or null hypothesis, respectively [41].

To account for the hierarchical nature of the data with individual effects being nested in cohorts being nested in studies, we used a multilevel structure with random effects at the study, cohort and effect level. In addition, we computed the variance-covariance matrix of all data points clustering individual effects at the cohort level. We assumed a correlation of effect sizes of rho = 0.5 to calculate the variance-covariance matrix. In line with previous studies [42,43], we additionally calculated sensitivity analyses to identify the impact of different values of rho (rho = 0.25 and 0.75, see Supplementary Tables 17-30). Result patterns were highly stable across different values of rho and only resulted in rounding differences at the second digit indicating that the results were robust. After the models were computed, we additionally used robust-variance estimation with cluster-robust inference at the cohort level, a recommended step for complex multivariate models to provide more accurate confidence intervals of the meta-analytic effects [44]. Bayesian models were fitted using the raw variance, as the variance–covariance structure could not be specified. Heterogeneity was assessed using Cochran’s Q to test whether effect sizes reflected a common underlying population effect. In addition, variance components (σ^2^) were estimated for each random-effects level to characterize heterogeneity across the multilevel structure. All heterogeneity estimates are reported in Supplementary Table 31.

Prior to moderation analyses, an overall model linking social connectedness to pain was computed. For this purpose, effect sizes estimating the relationship between measures of social disconnectedness (i.e., loneliness, social isolation and social exclusion) and pain were inverted. Due to the inversion, these outcomes reflect measures for the absence of disconnectedness and could thus be jointly analyzed on the same social connectedness scale as social support.

As preregistered, moderator analyses were conducted in separate models and only when statistical power for subgroup analyses was sufficient. Power calculations were based on the median study sample size, a small effect size (Fisher’s z = 0.10) consistent with the overall associations observed, and a moderate level of heterogeneity (I^2^ = 50%). Under these assumptions, at least ten effect sizes per moderator level were required to achieve 80% power (Supplementary Figure 1); moderator analyses were therefore restricted to levels meeting this criterion.

Because social connectedness constituted the primary multivariate outcome, all moderators were modelled as interactions with specific dimensions of social connectedness rather than pooled across dimensions. Pooling was avoided given that constructs such as loneliness and social support are not interchangeable and may reflect opposite ends of a continuum. Random slopes were included at both the study and cohort level to allow moderator effects to vary across the data hierarchy, reducing false-positive findings and improving model fit [45].

Post hoc tests of interactions with categorical moderators were conducted within each social connectedness dimension only (e.g., loneliness in clinical vs non-clinical samples), and not across different dimensions, in line with established meta-analytic practice [42]. Results were visualized using forest and orchard plots for categorical moderators and meta-regression plots with fitted confidence intervals for continuous moderators. Moderator levels included in the models but lacking a comparable secondary level due to data scarcity were not visualized for clarity.

### Risk of bias and study quality

Risk of bias was estimated using pre-specified quality indicators. To assess small study bias, we visually inspected funnel plots. Furthermore, we used precision-effect test and precision-effect estimate with standard errors (PET-PEESE) to test for small study bias by using the standard error as a moderator [46]. When evidence for small-study bias was detected, variance was included as a moderator and the model intercept was used as a bias-corrected effect estimate. PET–PEESE models were fitted using the same multilevel structure as the primary analyses to account for the hierarchical nature of the data.

To further characterize study quality, all included studies were evaluated on pre-registered indicators. For all studies, covariate adjustment, reporting of medication use, power analysis, preregistration, and peer review were coded as present (1) or absent (0). Experimental studies were additionally assessed for randomization, counterbalancing in within-subject designs, blinding, and attrition, with attrition bias coded as present if differential dropout exceeded 10%. Detailed risk-of-bias ratings are provided in Supplementary Table 1. An average quality score was computed and included as a moderator to examine the influence of study quality on the observed associations. Covariate use in particular was analyzed exploratorily as moderator to identify if effect sizes were influenced by covariate inclusion.

### Open science practices

All data and code are accessible under the following link xxx. This meta-analysis was preregistered on PROSPERO (CRD420250643896) prior to data extraction. Below, we report and justify all deviations from the preregistered protocol.

#### Deviation 1

The preregistration specified inclusion of explicit self-reported pain measures. However, behavioral pain outcomes such as pain threshold and tolerance were frequently reported in the original studies and are widely interpreted as reflecting self-reports on pain experience. These measures were elicited via explicit instructions and required participants to consciously evaluate and indicate their pain experience through, for example, hand withdrawal. We therefore included behavioral pain outcomes in the meta-analysis. In addition, studies comparing clinical pain samples with healthy controls were retained, even when explicit pain ratings were not reported.

#### Deviation 2

We were unable to systematically distinguish between between-person (e.g., individuals who are generally lonelier experience more pain) and within-person effects (e.g., momentary increases in an individual’s loneliness relative to their mean predicting pain) due to the limited availability of within-person estimates across all dimensions of social connectedness. We therefore could not further analyze this moderator in the present study and removed it from the list of extracted moderators.

#### Deviation 3

We initially planned to quantify publication bias using Bayesian RoBMA models. Although multivariate extensions of RoBMA have recently been developed, these models were computationally infeasible given the hierarchical complexity and number of effects in the present meta-analysis. Instead, Bayesian models were fitted using the brms package to complement frequentist analyses.

#### Deviation 4

As the initial data extraction took around six months, in July 2025 we ran an additional literature search to update for relevant articles published in 2025, which widened the scope of our initial search beyond the pre-registered 70 pages of Google Scholar.

## Results

### Social connectedness measures and pain

After application of the inclusion and exclusion criteria, data from 239 studies comprising 1,407,803 participants were included [68–314]. Descriptive data for the underlying studies and cohorts can be found in Tables 1 and 2.

**Table 1.** Study level (*n* = 239) characteristics.

| Characteristic | Category | n% (n) |
| --- | --- | --- |
| **Study design** | Experimental | 17.2% (41) |
|  | Observational | 82.8% (198) |
| **Data from observational studies** | Cross-sectional | 85.4% (169) |
|  | Longitudinal | 7.5% (15) |
|  | Both | 7.1% (14) |
| **Pain measurement** | Single item | 50.2% (120) |
|  | Questionnaire | 31.8% (76) |
|  | Both | 10.9% (26) |
|  | Unclear | 7.1% (17) |
| **Social measurement** | Single item | 62.8% (150) |
|  | Questionnaire | 18.0% (43) |
|  | Both | 8.8% (21) |
|  | Unclear | 10.4% (25) |
| **Chronicity measurement** | Trait measure | 75.8% (181) |
|  | State measure | 20.9% (50) |
|  | Both | 2.1% (5) |
|  | Unclear | 1.2% (3) |
| **Number of pain outcomes** | 1 | 71.1% (170) |
|  | 2 | 21.3% (51) |
|  | 3 or more | 7.5% (18) |
| **Number of social outcomes** | 1 | 86.6% (207) |
|  | 2 | 12.1% (29) |
|  | 3 or more | 1.3% (3) |
| **Report of medication use** | Yes | 37.2% (89) |
|  | No | 62.8% (150) |
| **Covariates included in analysis** | Yes | 83.3 % (199) |
|  | No | 16.7% (40) |
| **Preregistration** | Yes | 0.8% (2) |
|  | No | 99.2% (237) |
| **Power analysis** | Yes | 14.2% (34) |
|  | No | 85.8% (205) |
| **Peer-review** | Yes | 97.5% (233) |
|  | No | 2.5% (6) |

**Table 2.** Cohort-level (*j* = 286) characteristics.

| Characteristic | Category | n% (n) |
| --- | --- | --- |
| **Clinical status** | Clinical pain diagnosis | 53.5% (153) |
|  | Healthy cohort | 29.4% (84) |
|  | Mixed cohort / unclear classification | 17.1% (49) |
| **Pain etiology in clinical cohorts** | Primary pain disorder | 9.1% (14) |
|  | Secondary pain disorder | 51.0% (78) |
|  | No identifiable etiology | 39.9% (61) |

To address our primary research question, we first estimated an overall model in which all social variables were combined into a single index of social connectedness; measures of social disconnection (loneliness, social isolation, and social exclusion) were inverted to ensure a common direction of effects. In this model, greater social connectedness was associated with lower pain (*z* = - 0.09, 95% CI = [-0.11;-0.07], *t*_(283)_ = 9.30, *p* < .001, BF_10_ > 100, Figure 2A).

**Figure 2.**
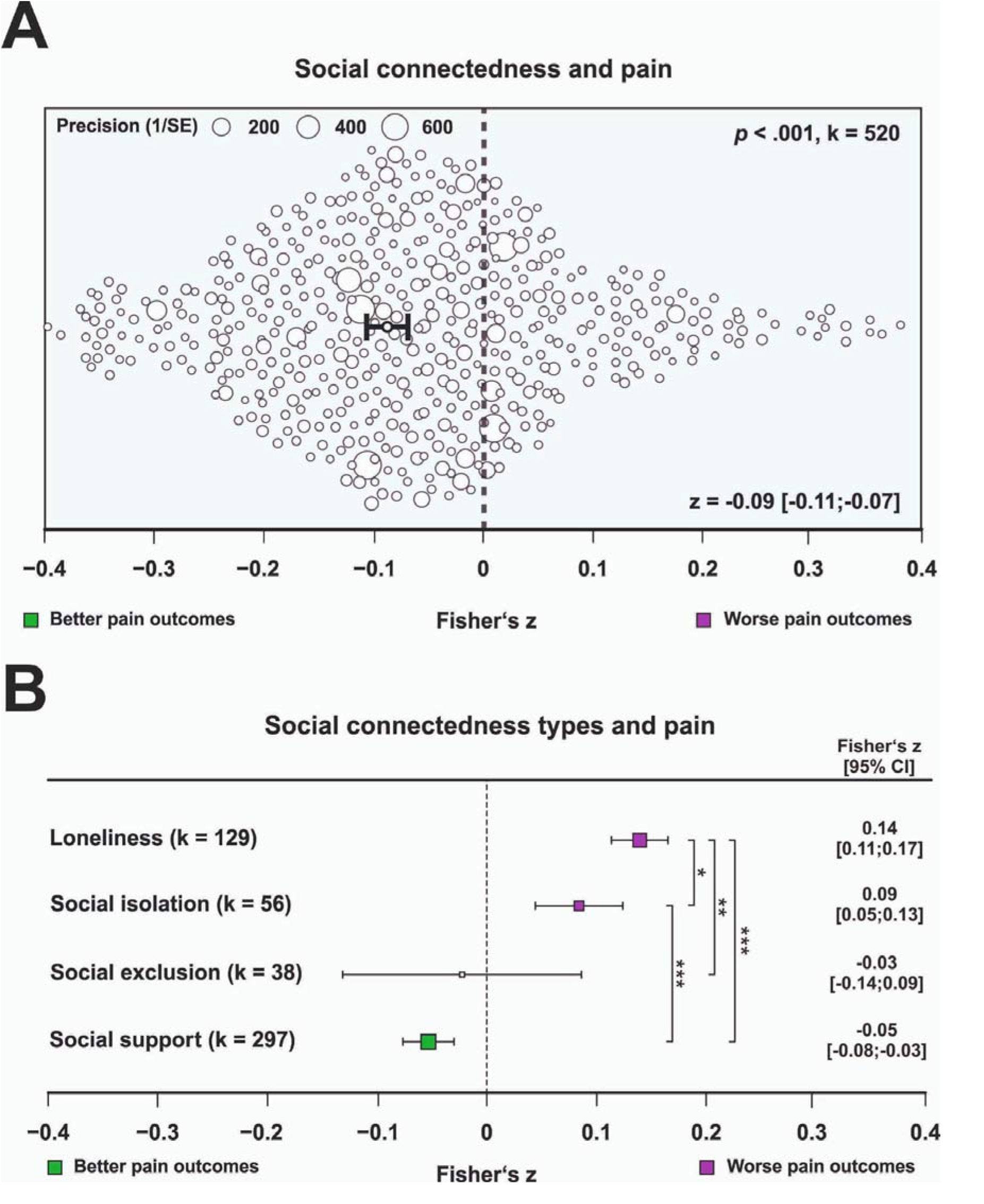
A) Orchard plot illustrating the association between overall social connectedness and pain. Social connectedness comprises loneliness, social isolation, and social exclusion (all inverted) as well as social support. Each dot represents a cohort-level effect size, with dot size reflecting precision. The number of included effects (k) is shown in the upper right. Pooled effects with 95% confidence intervals are displayed in the lower right and indicated by black dots and error bars. B) Forest plot showing associations between individual dimensions of social connectedness (loneliness, social support, social isolation, and social exclusion) and pain (non-inverted). The number of effects (k) contributing to each estimate is shown alongside each category. Pooled effects and 95% confidence intervals are reported numerically. Purple squares indicate significantly worse pain outcomes, green squares indicate significantly better pain outcomes, and grey squares indicate non-significant associations. Significant post hoc comparisons (two-sided *t* tests) are indicated. The corresponding orchard plot is provided in Supplementary Figure 2 (* = *p* < .05, ** = *p* < .01, *** = *p* < .001).

Analyses of specific dimensions revealed a robust positive association between pain and loneliness (*z* = 0.14, 95% CI = [0.11;0.17], *t*_(280)_ = 10.67, *p* < .001, BF_10_ > 100) and a smaller positive association with social isolation (*z* = 0.09 [0.05;0.13], *t*_(280)_ = 4.23, *p* < .001, BF_10_ > 100; Figure 2B). Social support was inversely associated with pain (*z* = -0.05 [-0.08;-0.03], *t*_(280)_ = 4.45, *p* < .001, BF_10_ > 100), whereas no significant association was observed for social exclusion (*z* = -0.03 [-0.14;0.09], *t*_(280)_ = 0.44, *p* = .659, BF_10_ = 0.69). These effects remained robust after adjusting for small study bias, adjusting for the use of covariates, and controlling for study quality. Complete bias assessment and funnel plots are provided in Supplementary results and Supplementary Figures 3–10. Since loneliness, social isolation and social support showed robust associations with pain, we compared them to meta-analytic findings from known biological and lifestyle risk factors affecting pain processing to contextualize their magnitude (Figure 3).

**Figure 3.**
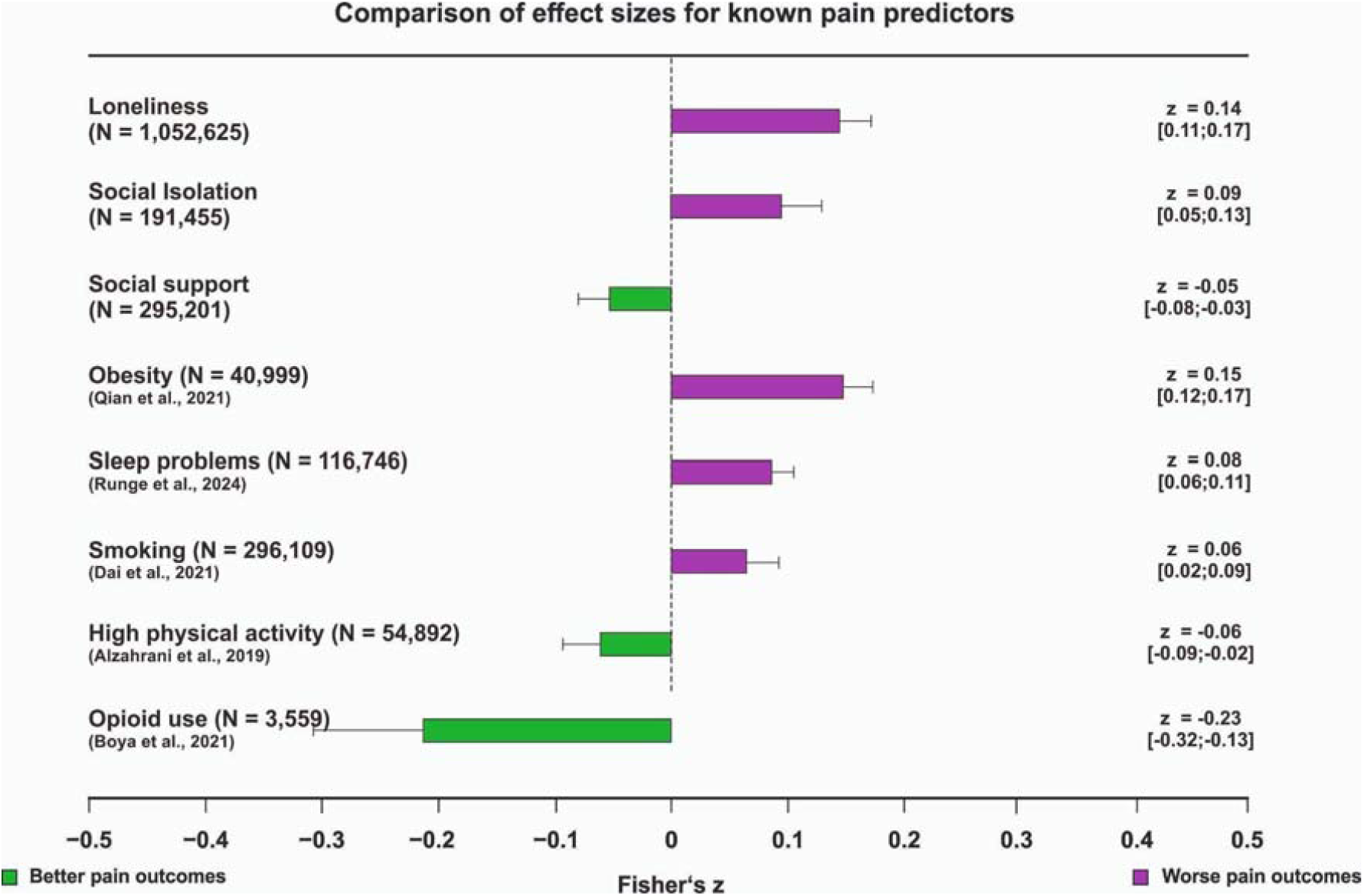
Comparison of the magnitude of associations for loneliness and social support with established biological and lifestyle risk factors for pain. Pooled effect estimates with 95% confidence intervals are shown numerically. Purple bars indicate significantly worse pain outcomes, whereas green bars indicate significantly better pain outcomes. Sample sizes reflect meta-analytic aggregates across studies. Effect estimates for loneliness and social support are derived from the present meta-analysis; estimates from prior meta-analyses were converted to Fisher’s *z* where necessary to ensure comparability. The effect for opioid use reflects placebo-controlled comparisons and represents an aggregate across buprenorphine, hydrocodone, oxycodone, and tramadol plus paracetamol.

Moderator analyses indicated that associations were largely robust across age and sex (Supplementary Figures 11–18). The only exception was social isolation, for which a stronger association with pain was observed in samples with a higher proportion of women (*F*_(1,33)_ = 5.74, *p* = .022, BF_10_ = 1.88). Additional analyses examined methodological factors including construct assessment (single- vs. multi-item measures) and experimental design (within- vs. between-participant), with detailed results reported in Supplementary Results. The corresponding forest and orchard plots for these analyses can be found in Supplementary Figures 19-26.

### Objective vs subjective measures of social connectedness

To enable direct comparisons between measures of social connectedness, effect sizes were again transformed to a common social connectedness metric. The absence of loneliness showed a stronger association with lower pain than the absence of social isolation (*t*_(280)_ = 2.50, *p* = .013, BF_10_ = 1.77) or social exclusion (*t*_(280)_ = 2.72, *p* = .007, BF_10_ = 17.32), and was also more strongly associated with lower pain than the presence of social support (*t*_(280)_ = 5.09, *p* < .001, BF_10_ > 100). This pattern suggests that subjective social disconnection in the form of loneliness is more closely linked to pain than objective isolation or experiences of ostracism. To examine whether the objective physical presence of another individual exerts stronger effects than subjectively perceived social support, we compared effects based on self-reported social support with those based on social presence. Social presence was associated with lower pain but the result for perceived support was marginal (perceived: *z* = -0.03 [-0.06;0.00], *t*_(122)_ = 1.92, *p* = .057, BF_10_ = 1.05, presence: *z* = -0.12 [-0.18;-0.07], *t*_(122)_ = 4.82, *p* < .001, BF_10_ > 100; Supplementary Figures 27A and 28A). The association was significantly stronger for social presence than for perceived support (*t*_(211)_ = 2.96, *p* = .004, BF_10_ = 8.22). Additional subtype analyses distinguished emotional versus instrumental support for perceived support (Supplementary Figures 27B and 28B) and familiar versus unfamiliar presence for social presence (Supplementary Figures 27C and 28C). No significant differences in effect magnitude were observed between these subtypes (emotional vs instrumental: *t*_(62)_ = 0.06, *p* = .956, BF_10_ = 0.31; familiar vs unfamiliar: *t*_(62)_ = 0.83, *p* = .409, BF_10_ = 0.37).

### Clinical vs non-clinical cohorts

If social connectedness represents a general determinant of pain, its associations with pain may extend across both clinical and healthy populations. We therefore compared effect sizes between clinical and non-clinical samples for loneliness and social support, where sufficient data were available (see Figure 4). No significant differences were observed between groups for either loneliness (*t*_(226)_ = 1.53, *p* = .128, BF_10_ = 0.36) or social support (*t*_(226)_ = 0.94, *p* = .347, BF_10_ = 0.27) with Bayes factors providing moderate support for the null hypothesis. Although correlational, these findings suggest that associations between social connectedness and pain are comparable in clinical and healthy populations, underscoring their broad relevance across the population. Within clinical cohorts, we explored whether associations between social connectedness and pain differed between primary and secondary pain disorders. Sufficient data for this comparison were available only for social support (≥10 effect sizes per group; Supplementary Figures 29-30). No significant difference was observed between primary and secondary pain disorders (*t*(84) = 0.19, *p* = .852, BF_10_ = 0.37), with Bayes factors supporting the null hypothesis. These findings suggest that the association between social support and pain does not differ by pain etiology. For that reason, we did not explore differences between clinical and non-clinical cohorts in the subsequent moderator analyses.

**Figure 4.**
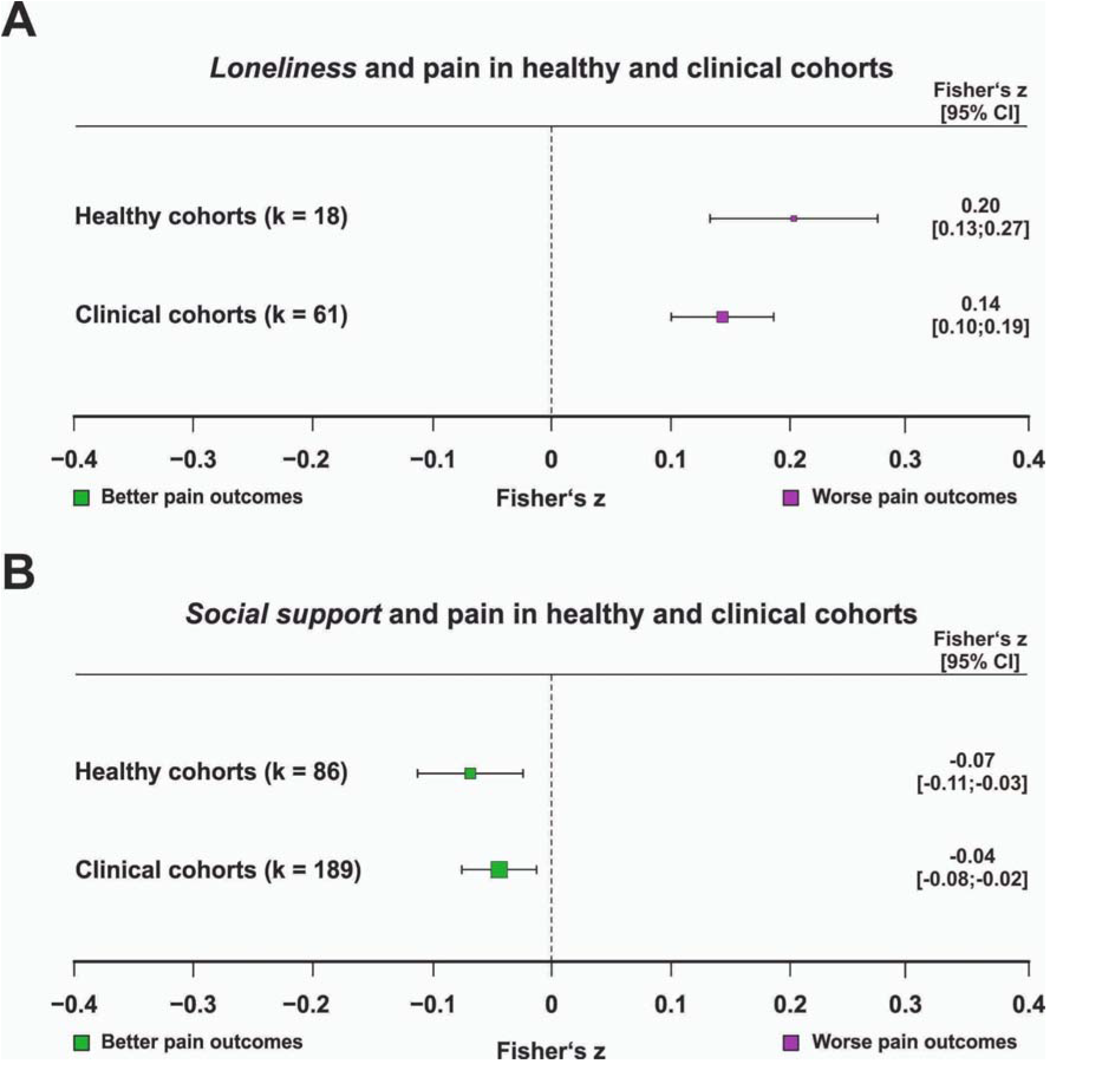
Association between pain and A) loneliness and B) social support in clinical and non-clinical samples. The number of contributing effect sizes (k) for each subgroup is shown; only subgroups with at least ten effect sizes were analyzed. Pooled effect estimates with 95% confidence intervals are reported numerically. Purple squares indicate significantly worse pain outcomes, green squares indicate significantly better pain outcomes, and grey squares indicate non-significant associations. The corresponding orchard plot is provided in Supplementary Figure 31.

### Temporal relationships between connectedness and pain

Most studies examining associations between pain and social connectedness relied on cross-sectional data, precluding inference about temporal directionality which is a necessary, though not sufficient, condition for causal interpretation. For loneliness and social support, however, sufficient data were available to examine longitudinal associations, with social variables predicting subsequent pain outcomes within individuals. Both cross-sectional and longitudinal analyses yielded significant associations for loneliness (cross-sectional: *z* = 0.14 [0.12;0.17], *t*_(211)_ = 10.28, *p* < .001, BF_10_ > 100; longitudinal: *z* = 0.09 [0.01;0.17], *t*_(211)_ = 2.16, *p* = .032, BF_10_ = 3.36; Figure 5A) and social support (cross-sectional: *z* = -0.05 [-0.07;-0.02], *t*_(211)_ = 3.23, *p* = .001, BF_10_ = 10.58; longitudinal: *z* = -0.08 [-0.13;-0.03], *t*_(211)_ = 3.07, *p* = .002, BF_10_ = 2.33; Figure 5B). Effect sizes did not differ significantly between cross-sectional and longitudinal designs for either loneliness (*t*_(211)_ = 1.30, *p* = .197, BF_10_ = 0.71) or social support (*t*_(211)_ = 1.22, *p* = 226, BF_10_ = 0.33), indicating comparable magnitudes across study designs.

**Figure 5.**
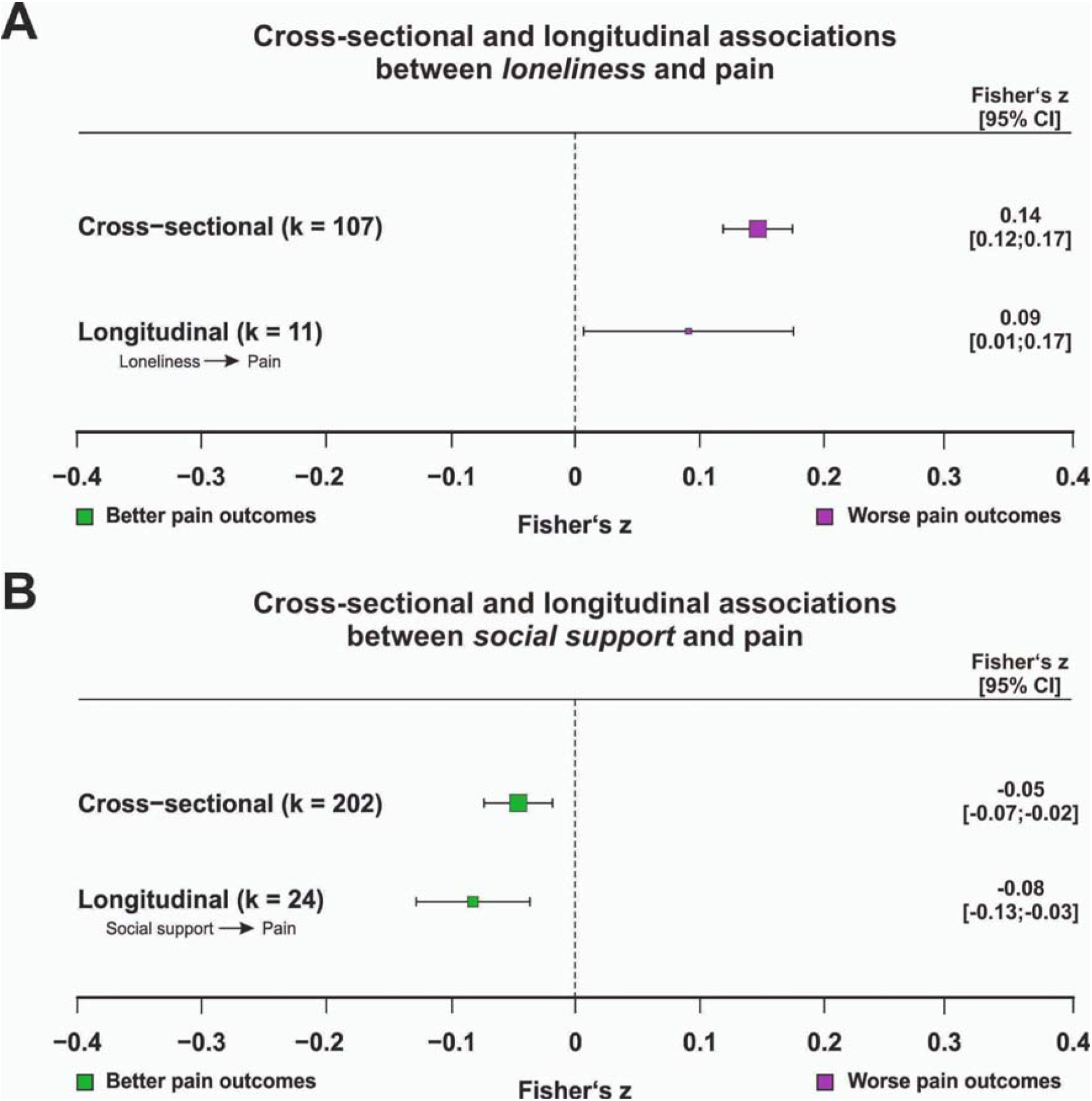
Association between pain and A) loneliness and B) social support in cross-sectional and longitudinal datasets. The number of contributing effect sizes (k) for each subgroup is shown; only subgroups with at least ten effect sizes were analyzed. Pooled effect estimates with 95% confidence intervals are reported numerically. Purple squares indicate significantly worse pain outcomes, green squares indicate significantly better pain outcomes, and grey squares indicate non-significant associations. The corresponding orchard plot is provided in Supplementary Figure 32.

### Specific pain outcomes

Given the multidimensional nature of pain, encompassing somatic, affective, cognitive, and functional components, we examined associations between social connectedness and specific pain outcomes where sufficient data were available. Loneliness was positively associated with pain frequency, a measure of how often painful episodes were experienced (*z* = 0.15 [0.11;0.20], *t*_(179)_ = 6.37, *p* < .001, BF_10_ > 100), pain intensity (*z* = 0.12 [0.09;0.16], *t*_(179)_ = 6.99, *p* < .001, BF_10_ > 100), and pain interference (*z* = 0.21 [0.14;0.27], *t*_(179)_ = 6.36, *p* < .001, BF_10_ > 100; see Figure 6A). Post hoc comparisons indicated that the association with pain interference was significantly stronger than that with pain intensity (*t*_(179)_ = 2.65, *p* = .008, BF_10_ = 2.21). Social isolation was positively associated with pain interference (*z* = 0.18 [0.06;0.30], *t*_(179)_ = 2.92, *p* = .004, BF_10_ = 45.26) and intensity (*z* = 0.09 [0.02;0.16], *t*_(179)_ = 2.60, *p* = .010, BF_10_ = 6.57; Figure 6B). Post hoc comparison indicated that there was no significant difference between the two pain outcomes (*t*_(179)_ = 1.14, *p* = .254, BF_10_ = 0.67). In contrast, social exclusion was not significantly associated with pain threshold (*z* = -0.10 [-0.34;0.13], *t*_(179)_ = 0.88, *p* = .382, BF_10_ = 0.84) or pain tolerance (*z* = -0.08 [-0.35;0.19], *t*_(179)_ = 0.60, *p* = .547, BF_10_ = 0.79; Figure 6C), consistent with the overall null effect observed for this dimension. For social support, sufficient data were available for six pain outcomes (Figure 6D). Higher social support was associated with lower pain unpleasantness (*z* = -0.11 [-0.16;-0.06], *t*_(179)_ = 4.39, *p* < .001, BF_10_ = 10.69), duration (*z* = -0.09 [-0.17;-0.01], *t*_(179)_ = 2.17, *p* = .032, BF_10_ = 1.90) and intensity (*z* = -0.05 [-0.08;-0.02], *t*_(179)_ = 3.00, *p* = .003, BF_10_ = 4.95). In contrast, social support was not significantly related to pain catastrophizing (*z* = -0.05 [-0.13;0.02], *t*_(179)_ = 1.49, *p* = .137, BF_10_ = 0.65), pain-related disability (*z* = -0.05 [-0.12;0.01], *t*_(179)_ = 1.62, *p* = .107, BF_10_ = 0.70), or pain interference (*z* = 0.01 [-0.04;0.06], *t*_(179)_ = 0.45, *p* = .655, BF_10_ = 0.28). Post hoc tests revealed a stronger negative association for pain unpleasantness compared to both pain intensity (*t*_(179)_ = 3.23, *p* = .002, BF_10_ = 0.95) and pain interference (*t*_(179)_ = 4.63, *p* < .001, BF_10_ = 5.93).

**Figure 6.**
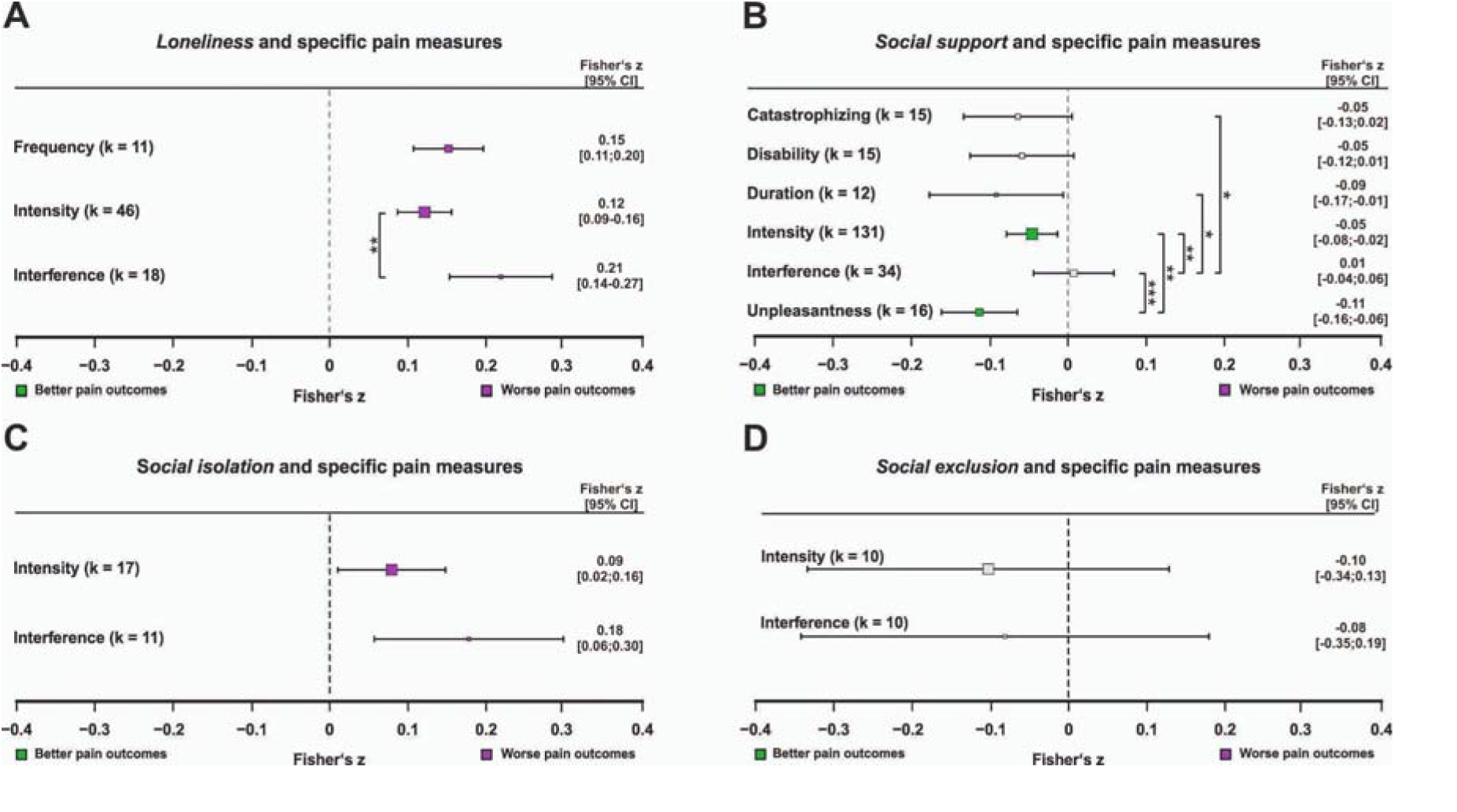
Association between A) loneliness, B) social support, C) social isolation, and D) social exclusion and specific pain outcomes. The number of contributing effect sizes (k) for each outcome is shown. Only outcomes with at least ten effect sizes were analyzed. Pooled effect estimates with 95% confidence intervals are displayed numerically. Purple squares indicate significantly worse pain outcomes, green squares indicate significantly better pain outcomes, and grey squares indicate non-significant associations. Significant post hoc comparisons (two-sided *t* tests) are indicated (* = *p* < .05, ** = *p* < .01, *** = *p* < .001). The corresponding orchard plot is provided in Supplementary Figure 33.

### The role of outcome chronicity

To examine the influence of pain outcome chronicity, we compared state (momentary) and trait-like (longer-term) assessments of pain. Sufficient data for both assessment types were available for loneliness and social support. Across assessment types, associations mirrored the overall pattern, with loneliness showing positive associations with pain (state: *z* = 0.13 [0.07;0.20], *t*_(272)_ = 3.91, *p* < .001, BF_10_ = 2.36; trait-like: *z* = 0.15 [0.12;0.17], *t*_(272)_ = 10.27, *p* < .001, BF_10_ > 100; Supplementary Figures 34A and 35A) and social support showing negative associations (state: *z* = -0.09 [-0.14;-0.05], *t*_(277)_ = 4.06, *p* < .001, BF_10_ = 6.51; trait-like: *z* = -0.05 [-0.07;-0.02], *t*_(277)_ = 3.47, *p* < .001, BF_10_ = 19.15; Supplementary Figures 34B and 35B). No significant differences were observed between state and trait-like assessments for either loneliness (*t*_(272)_ = 0.30, *p* = .765, BF_10_ = 0.58) or social support (*t*_(272)_ = 1.85, *p* = .065, BF_10_ = 0.45) . Analyses based on the chronicity of social outcomes yielded comparable results, reflecting the frequent alignment of assessment timeframes for pain and social outcomes (Supplementary Figure 19-20).

## Discussion

This meta-analysis provides a comprehensive quantitative synthesis of how multiple dimensions of social connectedness (i.e., loneliness, social support, social isolation, and social exclusion) relate to the experience of pain. Integrating frequentist and Bayesian evidence from 239 studies comprising 1,407,803 participants, we found robust evidence for an overall negative association between social connectedness and pain. However, dimension-specific analyses revealed substantial heterogeneity. Even if adjusted for publication bias, loneliness, social isolation and social support remained reliably associated with pain, with loneliness emerging as the strongest correlate. Associations for loneliness and social support did not differ between clinical and non-clinical samples and were not moderated by age or sex suggesting broad public health relevance. Furthermore, these associations were stable independent of study protocol with converging evidence from cross-sectional, longitudinal and experimental studies suggesting that the link between social connectedness and pain is robust.

A central aim of this meta-analysis was to distinguish between dimensions of social connectedness and their associations with pain experience. Our findings indicate that loneliness, a subjective appraisal of social disconnection, showed robust associations with pain, whereas social isolation, an objective structural indicator, exhibited weaker yet still significant effects. This pattern suggests that the experiential quality of social relationships may be more tightly coupled to pain than their structural characteristics, potentially reflecting dynamic and reciprocal processes between social appraisal and pain. It aligns with broader health literature demonstrating partially independent effects of subjective and objective social factors [47] and suggesting that perceived relationship quality may be more consequential for health than network size or contact frequency [48,49]. Interestingly, the opposite pattern emerged for social support. Objective social presence, that was typically examined in experimental paradigms, showed stronger pain-reducing effects than self-reported perceived support. While this might suggest a different relation for social presence compared to perceived support, this discrepancy should be interpreted cautiously, as it possibly reflects methodological rather than substantive differences. Perceived social support was predominantly assessed in observational studies, whereas social presence was exclusively studied in experimental settings involving acute pain induction, where immediate social cues may exert particularly strong modulatory effects.

When examined on the same underlying dimension of social connectedness, loneliness showed a significantly stronger association with pain than social support, consistent with the notion that these constructs capture related but distinct facets of social connectedness [47,50]. It should however be noted that a number of studies have reported positive associations between social support and pain. This pattern is consistent with operant and interpersonal models of pain, which propose that certain supportive responses, such as reassurance or solicitous attention, may inadvertently reinforce pain behaviors or intensify emotional pain communication, thereby contributing to heightened pain reports [51]. This could potentially explain why social support showed higher variability in its relationship with pain and a generally weaker association overall as its effects may depend more strongly on the interpersonal context [29]. A similar contextual dependency may underlie the null effect observed for social exclusion in the present meta-analysis. Prior research suggests that the direction of social exclusion effects on pain may depend on intensity, with milder exclusion increasing sensitivity and more severe exclusion producing numbing responses [52]. However, the limited number of studies precluded formal subgroup analyses to test this possibility. Interpretation of this finding is further limited by pronounced heterogeneity in how exclusion was operationalized, ranging from laboratory paradigms (e.g., Cyberball) to structural deprivation or self-reported ostracism.

A further key aim of the present study was to examine whether the association between social connectedness and pain varies across demographic and clinical subgroups, thereby informing its potential public health relevance. When comparing clinical and non-clinical samples, the evidence did not indicate systematic differences between these groups. This finding is notable given well-established differences in pain prevalence, severity, and underlying mechanisms across populations [53]. One might have anticipated that social connectedness would relate differently to pain among individuals with diagnosed pain conditions than in community samples, as chronic pain is frequently accompanied by maladaptive cognitive patterns, including negative appraisal and catastrophizing [54], which overlap with cognitive features observed in chronic loneliness [55,56]. Our findings suggest that social connectedness is relevant not only for pain management in clinical settings but also for the prevention of pain-related problems in the general population. This pattern supports the view that links between social experience and pain reflect fundamental biopsychosocial processes rather than effects confined to specific diagnostic categories. The broad public health relevance of these associations is further underscored by the absence of moderation by key demographic factors such as age and sex. Although age and sex are well-established determinants of chronic pain prevalence [57], the association between social connectedness and pain appeared consistent across these groups. This stability suggests that strengthening social connection may confer benefits across the lifespan and across sexes. Given the high prevalence of both chronic pain and loneliness, interventions targeting social connectedness may therefore represent a scalable and widely applicable public health strategy.

Translating these findings into intervention strategies requires evidence that social connectedness exerts a causal influence on pain, beyond their observed co-variation. Although causal inference is inherently constrained by the predominance of observational studies in our meta-analysis, longitudinal evidence provides important support for temporal ordering: earlier measures of loneliness and social support predict later pain outcomes, with effect sizes comparable to cross-sectional associations. Prospective designs suggest that social isolation more often precedes increases in pain interference than vice versa [2]. Temporal precedence, while necessary but not sufficient for causal inference, nonetheless strengthens the plausibility of directional effects. Converging evidence from experimental studies further underscores the interventional potential of social connectedness, illustrating that social support can reduce pain under controlled conditions. Thus, at least for the subset of experimental studies in our meta-analysis, a causal relationship is demonstrable. Together, these findings suggest that social connectedness is not merely correlated with pain but may represent a modifiable target for intervention, with the capacity to influence both acute pain responses and potentially longer-term pain trajectories.

Although a direct causal relationship between social connectedness and pain processing appears plausible, the underlying mechanisms remain incompletely understood. Affective factors such as depression and anxiety likely represent partial mediators, as loneliness and low social support are consistently associated with elevated affective symptomatology, which in turn modulates pain perception [58,59]. Evidence from chronic pain populations further suggests that anxiety may partially mediate the association between social connectedness and pain outcomes [60]. However, social disconnection cannot be fully reduced to generalized affective distress. Neurobiological evidence demonstrates a dissociation between loneliness and social anxiety at both behavioral and neural levels, indicating distinct underlying mechanisms despite phenotypic overlap [61]. Consistent with this view, we did not observe systematic differences between studies adjusting for psychiatric covariates and those that did not, arguing against a purely indirect or fully mediated association. Future longitudinal and experimental research is needed to formally test affective mediation models and clarify causal pathways linking social connectedness and pain.

Importantly, the magnitude of the loneliness–pain association was comparable to that reported for established biological and lifestyle risk factors such as obesity [62], and exceeded associations observed for sleep disturbances [63] and smoking [64], underscoring its clinical relevance. Associations involving social support were smaller but robust, approximating effect sizes reported for high physical activity [65]. Although pharmacological treatments, particularly opioids, produce substantially larger analgesic effects [66], such comparisons provide an incomplete benchmark. Our findings indicate that social connectedness constitutes a meaningful non-pharmacological determinant of pain, operating at an effect size comparable to other widely targeted lifestyle and psychosocial factors. These results support a stronger integration of social factors into multimodal pain management approaches, including routine assessment of patients’ perceived social connection and, where indicated, interventions aimed at strengthening supportive relationships or addressing deficits in desired social engagement.

Finally, this meta-analysis sought to differentiate the associations of distinct social connectedness constructs with specific pain outcomes. Loneliness was consistently associated with all examined pain outcomes for which sufficient data was available, whereas social support demonstrated more selective associations. This pattern may suggest that loneliness relates to pain across multiple domains, while associations involving social support appear more outcome-specific. Such a distribution is in line with broader evidence linking loneliness to adverse health indicators across diverse domains [67]. At the same time, this distinction should be interpreted cautiously as social support was operationalized heterogeneously across studies, whereas loneliness was typically assessed using conceptually coherent instruments. Measurement heterogeneity may therefore partly explain the more selective associations observed for social support.

Several considerations should inform the interpretation of these findings. Randomized controlled trials targeting loneliness or social isolation remain scarce, limiting translational inference for a causal role of these factors. Experimental studies investigating the role of social support have often involved relatively small samples and were rarely pre-registered, and reporting of factors such as medication use has not always been comprehensive. In addition, evidence of publication bias was detected. Variability in the operationalization of social support, together with incomplete reporting of psychiatric comorbidities, particularly depression, introduces further uncertainty in estimating the precision of observed associations. Finally, although moderator analyses were conducted in accordance with pre-registered power assumptions, these did not explicitly incorporate the additional complexity of multilevel random-slope models, which may have reduced statistical power for certain subgroup analyses approaching the minimum threshold.

## Conclusions

This meta-analysis provides converging evidence that social connectedness is reliably associated with the subjective experience of pain, with loneliness emerging as the strongest correlate and pointing to the central relevance of subjective social appraisals relative to structural social circumstances. The associations generalized across populations, demographics, and pain characteristics, highlighting broad public health relevance. Although the available evidence is predominantly correlational, experimental findings support a causal role for social support in modulating pain. Adequately powered, theory-driven interventions are now needed to determine whether improving social functioning, particularly reducing loneliness, can yield meaningful benefits across sensory, affective, cognitive, and functional dimensions of pain.

## Data Availability

Data availability statement
All data are available under the following link: https://osf.io/6gkxj/
Code availability statement
All code is available under the following link: https://osf.io/6gkxj/

https://osf.io/6gkxj/

## Data availability statement

All data are available under the following link: https://osf.io/6gkxj/

## Code availability statement

All code is available under the following link: https://osf.io/6gkxj/

## Author contributions statement

A.P.: Conceptualization, Data curation, Formal analysis, Funding acquisition, Investigation, Methodology, Project administration, Visualization, Writing - original draft, Writing - review & editing

S.E.: Conceptualization, Investigation, Writing - review & editing

A.I.: Conceptualization, Investigation, Writing - review & editing

L.O.: Conceptualization, Investigation, Writing - review & editing

D.S.: Conceptualization, Funding acquisition, Investigation, Project administration, Supervision, Visualization, Writing - review & editing

J.P.: Conceptualization, Data curation, Formal analysis, Investigation, Methodology, Project administration, Supervision, Visualization, Writing - original draft, Writing - review & editing

