## Supplementary Materials for "A multivariate meta-analysis on the relationship between social connectedness and pain"

**Table of Contents – Supplementary Figures**

| Supplementary Figure 1. Preregistered power analysis for moderator and subgroup analyses. | 5 |
| --- | --- |
| Supplementary Figure 2. Overall associations between dimensions of social connectedness and pain (orchard plot). | 6 |
| ***Publication Bias and Study Quality*** | |
| Supplementary Figure 3. Funnel plot meta-regression (standard error) for loneliness. | 7 |
| Supplementary Figure 4. Funnel plot meta-regression (standard error) for social support. | 8 |
| Supplementary Figure 5. Funnel plot meta-regression (standard error) for social isolation. | 9 |
| Supplementary Figure 6. Funnel plot meta-regression (standard error) for social exclusion. | 10 |
| Supplementary Figure 7. Meta-regression of study quality and the loneliness–pain association. | 11 |
| Supplementary Figure 8. Meta-regression of study quality and the social support–pain association. | 12 |
| Supplementary Figure 9. Meta-regression of study quality and the social isolation–pain association. | 13 |
| Supplementary Figure 10. Meta-regression of study quality and the social exclusion–pain association. | 14 |
| ***Demographic moderators***  Supplementary Figure 11. Meta-regression of cohort mean age and the loneliness–pain association. | 15 |
| Supplementary Figure 12. Meta-regression of cohort mean age and the social support–pain association. | 16 |
| Supplementary Figure 13. Meta-regression of cohort mean age and the social isolation–pain association. | 17 |
| Supplementary Figure 14. Meta-regression of cohort mean age and the social exclusion–pain association. | 18 |
| Supplementary Figure 15. Meta-regression of cohort gender ratio and the loneliness–pain association. | 19 |
| Supplementary Figure 16. Meta-regression of cohort gender ratio and the social support–pain association. | 20 |
| Supplementary Figure 17. Meta-regression of cohort gender ratio and the social isolation–pain association. | 21 |
| Supplementary Figure 18. Meta-regression of cohort gender ratio and the social exclusion–pain association. | 22 |
| ***State vs. Trait Social Measurement***  Supplementary Figure 19. Social support associations with pain for state vs. trait social support measures (forest plot).  Supplementary Figure 20. Same comparisons as Supplementary Figure 19 (orchard plot). | 23  24 |

| ***Construct Measurement*** | |
| --- | --- |
| Supplementary Figure 21. Associations between pain and loneliness, social support, and social isolation for single-item vs. multi-item pain measures (forest plot). | 25 |
| Supplementary Figure 22. Same comparisons as Supplementary Figure 21 (orchard plot). | 26 |
| Supplementary Figure 23. Associations between pain and loneliness, social support, and social isolation for single-item vs. multi-item social measures (forest plot). | 27 |
| Supplementary Figure 24. Same comparisons as Supplementary Figure 23 (orchard plot). | 28 |
| ***Experimental Design*** | |
| Supplementary Figure 25. Social support–pain associations in within-participant vs. between-participant designs (forest plot). | 29 |
| Supplementary Figure 26. Same comparisons as Supplementary Figure 25 (orchard plot). | 30 |

| ***Operationalisation of Social Support*** | | |
| --- | --- | --- |
| Supplementary Figure 27. Pain associations for perceived vs. present support, emotional vs. instrumental support, and familiar vs. unfamiliar presence (forest plot).  Supplementary Figure 28. Same comparisons as Supplementary Figure 27 (orchard plot).  ***Clinical characteristics*** | | 31  32 |
| Supplementary Figure 29. Social support–pain associations in primary vs. secondary pain disorders (forest plot). | 32 | |
| Supplementary Figure 30. Social support–pain associations in primary vs. secondary pain disorders (orchard plot).  Supplementary Figure 31. Associations between loneliness, social support, and pain in clinical vs. non-clinical samples (orchard plot).  ***Temporal Directionality***  Supplementary Figure 32. Loneliness and social support associations with pain in cross-sectional vs. longitudinal studies (orchard plot).  ***Pain Outcome Specificity*** | 33  34  35 | |
| Supplementary Figure 33. Associations between social connectedness dimensions and specific pain outcomes (orchard plot). | 36 | |
| ***State vs. Trait Pain Measurement*** | | |
| Supplementary Figure 34. Loneliness and social support associations with pain for state vs. trait pain measures (forest plot). | 37 | |
| Supplementary Figure 35. Same comparisons as Supplementary Figure 34 (orchard plot). | 38 | |

**Table of Contents – Supplementary Tables**

| ***Studies information*** | |
| --- | --- |
| Supplementary Table 1. Study quality assessment ratings for all included studies. | 40 |
| Supplementary Table 2. Studies excluded from the meta-analysis with reasons for exclusion. | 50 |
| ***Bayes Factor Analysis*** | |
| Supplementary Table 3. Bayes Factor analysis for overall effect. | 59 |
| Supplementary Table 4. Bayes Factor analysis by social connectedness dimension. | 60 |
| Supplementary Table 5. Bayes Factor analysis for social connectedness dimension by pain type interaction. | 61 |
| Supplementary Table 6. Bayes Factor analysis for perceived vs. present social support. | 62 |
| Supplementary Table 7. Bayes Factor analysis for social support types. | 63 |
| Supplementary Table 8. Bayes Factor analysis for social connectedness by clinical sample interaction. | 64 |
| Supplementary Table 9. Bayes Factor analysis for social connectedness by pain classification interaction. | 65 |
| Supplementary Table 10. Bayes Factor analysis for social connectedness by association type interaction. | 66 |
| Supplementary Table 11. Bayes Factor analysis for social connectedness by study design interaction. | 67 |
| Supplementary Table 12. Bayes Factor analysis for social connectedness by pain measurement interaction. | 68 |
| Supplementary Table 13. Bayes Factor analysis for social connectedness by social measurement interaction. | 69 |
| Supplementary Table 14. Bayes Factor analysis for social connectedness by trait/state pain interaction. | 70 |
| Supplementary Table 15. Bayes Factor analysis for social connectedness by trait/state social measurement interaction. | 71 |
| Supplementary Table 16. Bayes Factor analysis for publication bias tests and continuous moderators. | 72 |
| ***Sensitivity Analysis*** | |
| Supplementary Table 17. Sensitivity analysis for overall effect across different correlation assumptions (ρ = 0.25, 0.5, 0.75). | 73 |
| Supplementary Table 18. Sensitivity analysis by social connectedness dimension across different correlation assumptions. | 74 |
| Supplementary Table 19. Sensitivity analysis for social connectedness by pain type interaction across different correlation assumptions. | 75 |
| Supplementary Table 20. Sensitivity analysis for perceived vs. present social support across different correlation assumptions. | 77 |
| Supplementary Table 21. Sensitivity analysis for social support types across different correlation assumptions. | 78 |
| Supplementary Table 22. Sensitivity analysis for social connectedness by clinical sample interaction across different correlation assumptions. | 79 |
| Supplementary Table 23. Sensitivity analysis for social connectedness by pain classification interaction across different correlation assumptions.  Supplementary Table 24. Sensitivity analysis for social connectedness by outcome chronicity interaction across different correlation assumptions. | 80  81 |
| Supplementary Table 25. Sensitivity analysis for social connectedness by study design interaction across different correlation assumptions. | 82 |
| Supplementary Table 26. Sensitivity analysis for social connectedness by pain measurement interaction across different correlation assumptions. | 83 |
| Supplementary Table 27. Sensitivity analysis for social connectedness by social measurement interaction across different correlation assumptions. | 84 |
| Supplementary Table 28. Sensitivity analysis for social connectedness by trait/state pain interaction across different correlation assumptions. | 85 |
| Supplementary Table 29. Sensitivity analysis for social connectedness by trait/state social measurement interaction across different correlation assumptions. | 86 |
| Supplementary Table 30. Sensitivity analysis for publication bias tests and continuous moderators across different correlation assumptions. | 87 |
| Supplementary Table 31. Heterogeneity estimates for all models. | 88 |

**Table of Contents – Supplementary Results**

| Bias Assessment | 89 |
| --- | --- |
| Moderator analyses of methodological factors | 89 |

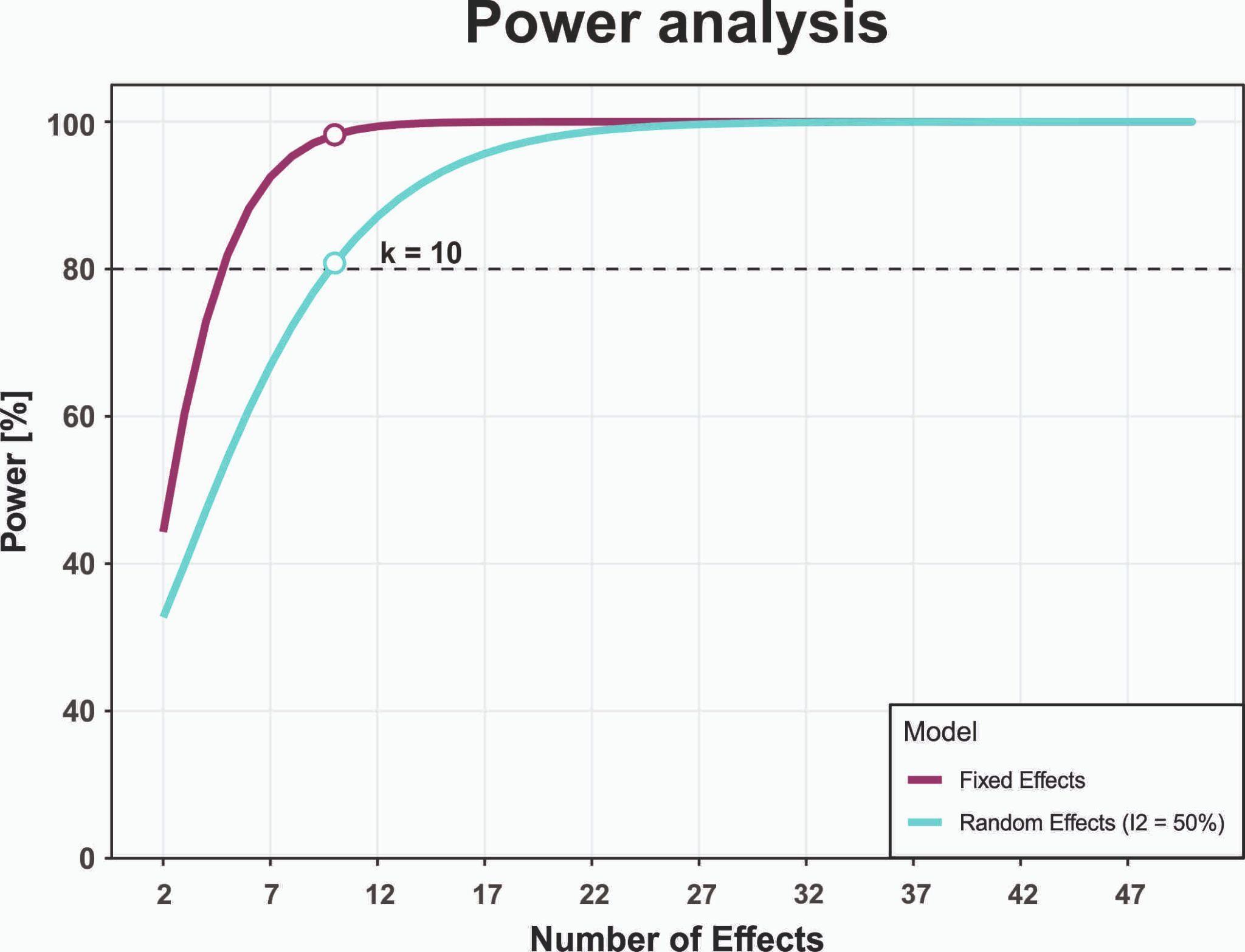

**Supplementary Figure 1.** Preregistered power analysis. Power calculations were based on the median study sample size (n = 168) and a small expected effect size (r = 0.10). Assuming moderate heterogeneity (I² = 50%), a minimum of 10 effect sizes per moderator level was required to achieve 80% statistical power.

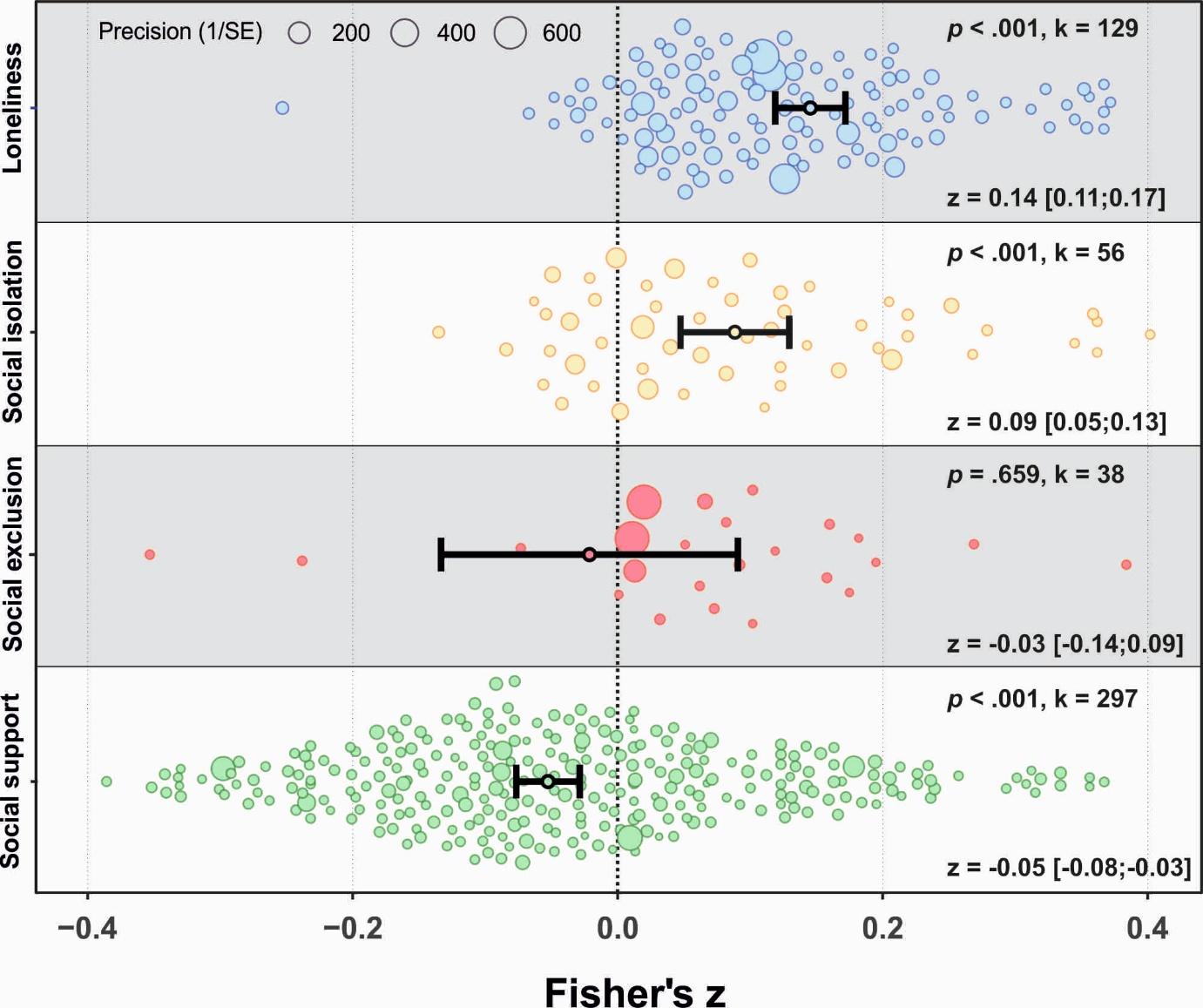
 **Supplementary Figure 2.** Orchard plot illustrating associations between dimensions of social connectedness and pain. Each dot represents a cohort-level effect size, with dot size reflecting precision. The number of included effect sizes (k) is shown in the upper right of each panel. Pooled effects with 95% confidence intervals are displayed in the lower right and indicated by black dots and error bars.

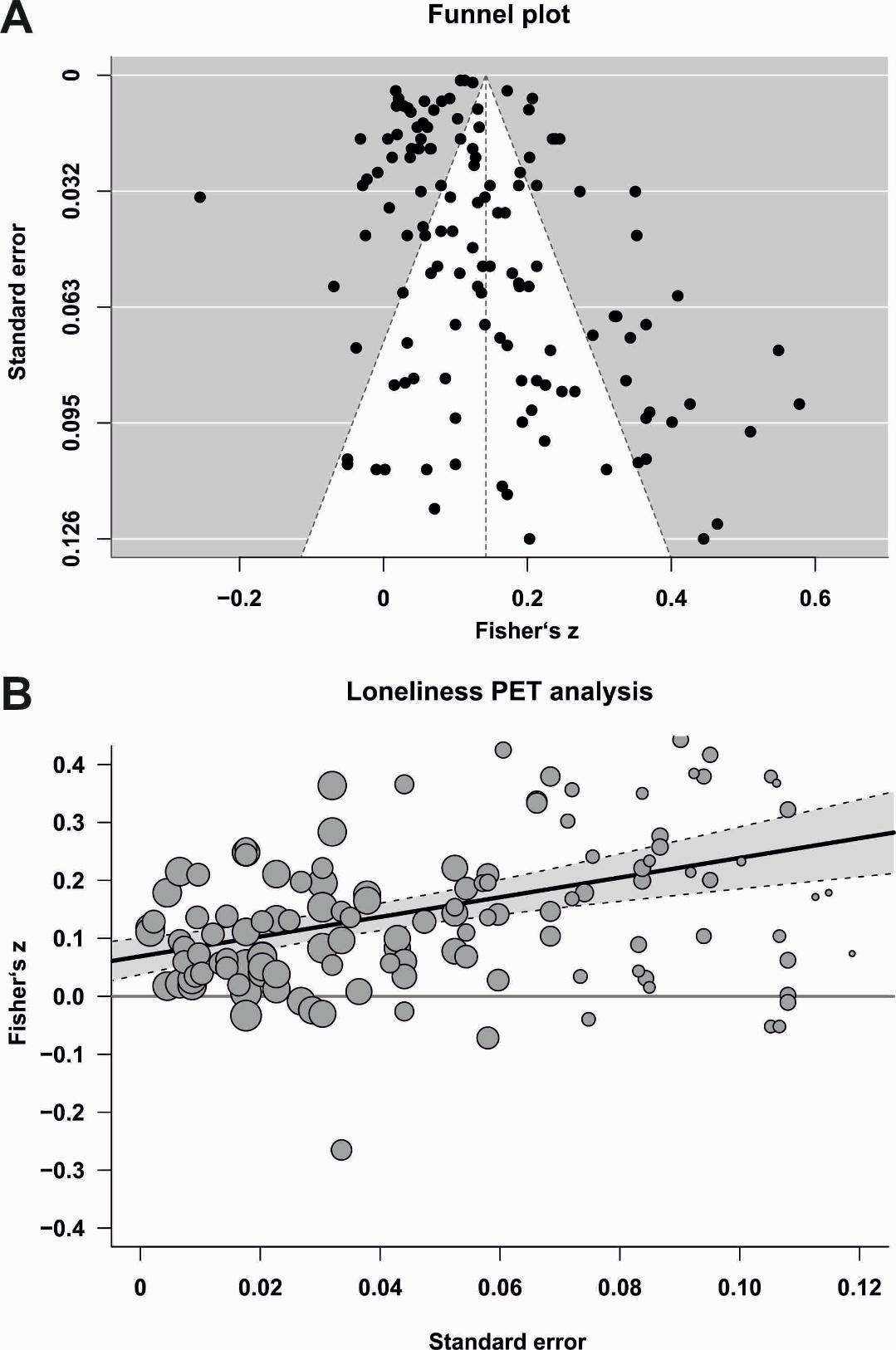

**Supplementary Figure 3.** Meta-regression depicting the relationship between effect size magnitude and standard error for loneliness. Each dot represents an effect size from an individual dataset, with dot size indicating study precision. The shaded band denotes the 95% confidence interval around the regression line.

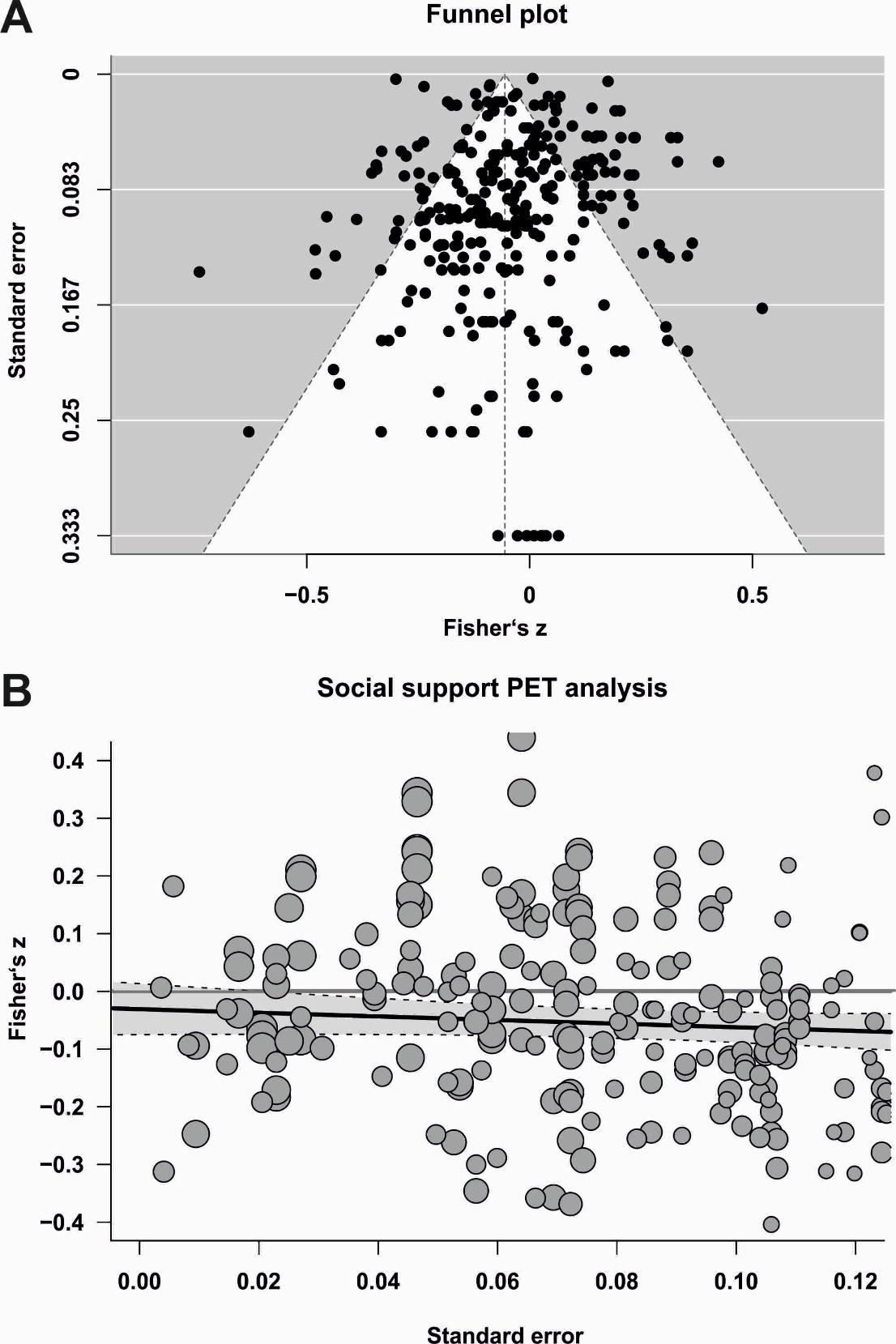

**Supplementary Figure 4.** Meta-regression showing the relationship between effect size magnitude and standard error for social support. Each dot represents an effect size from an individual dataset, with dot size indicating study precision. The shaded band denotes the 95% confidence interval around the regression line.

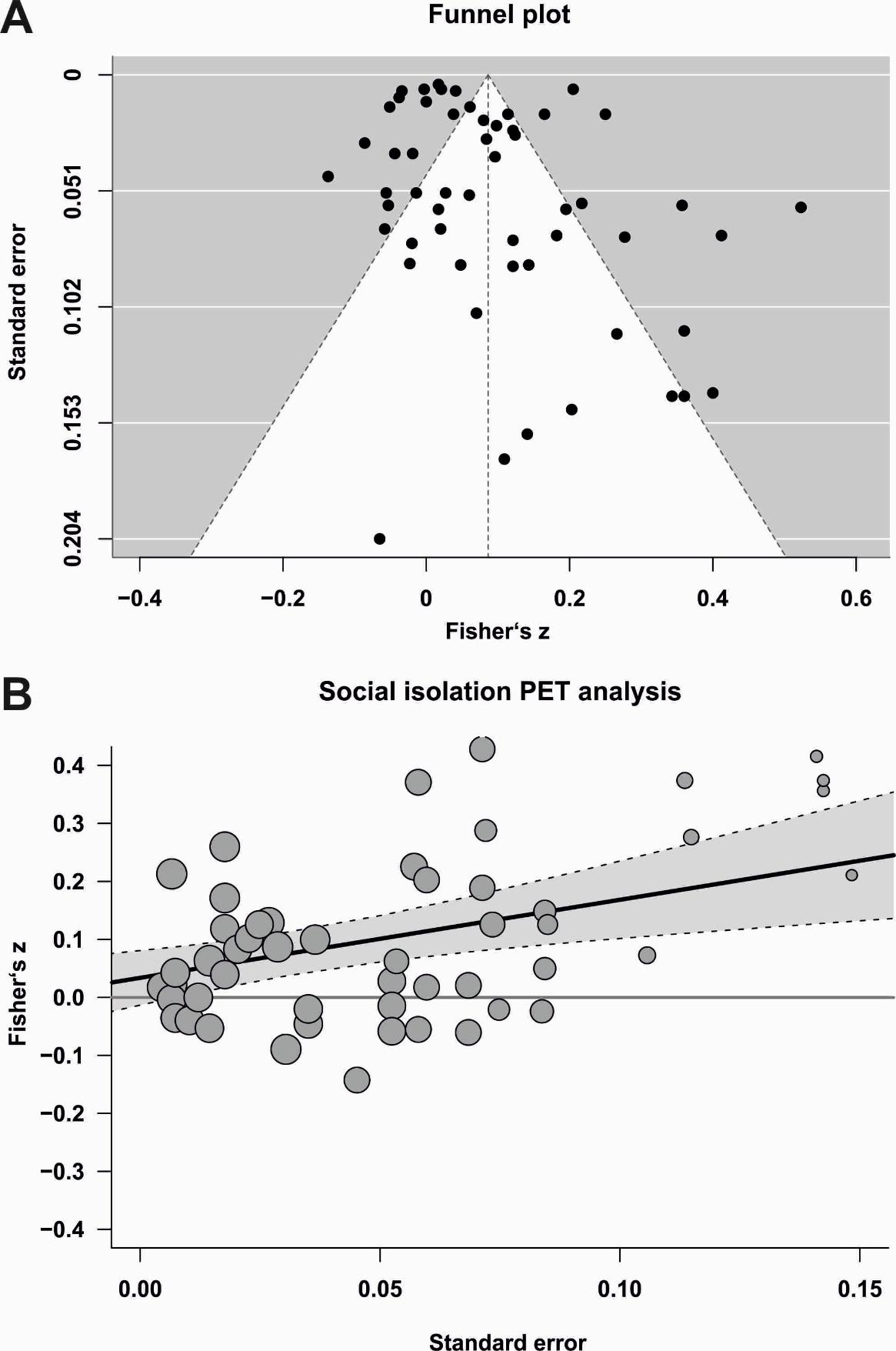

**Supplementary Figure 5.** Meta-regression showing the relationship between effect size magnitude and standard error for social isolation. Each dot represents an effect size from an individual dataset, with dot size indicating study precision. The shaded band denotes the 95% confidence interval around the regression line..

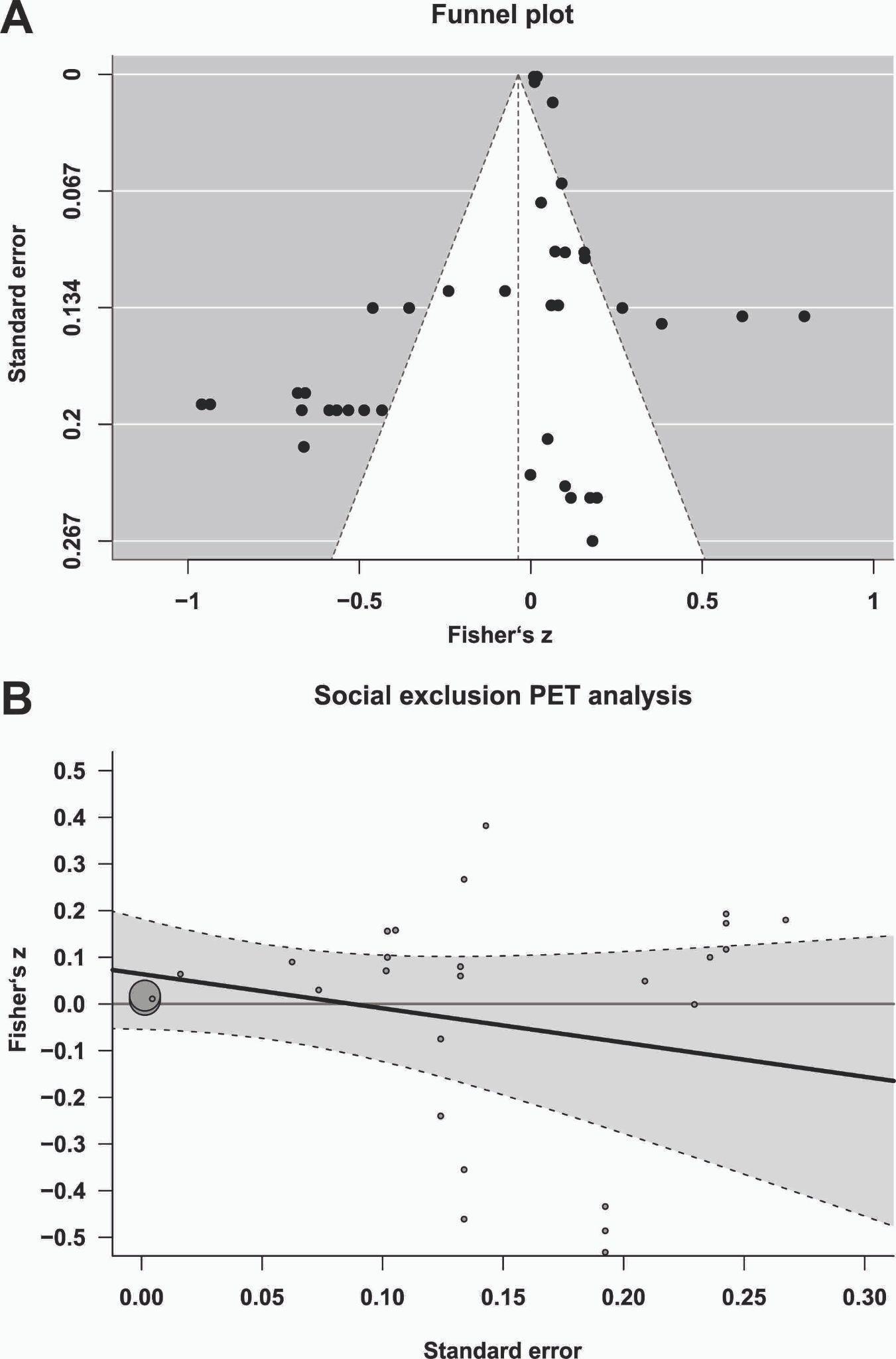

**Supplementary Figure 6.** Meta-regression showing the relationship between effect size magnitude and standard error for social exclusion. Each dot represents an effect size from an individual dataset, with dot size indicating study precision. The shaded band denotes the 95% confidence interval around the regression line.

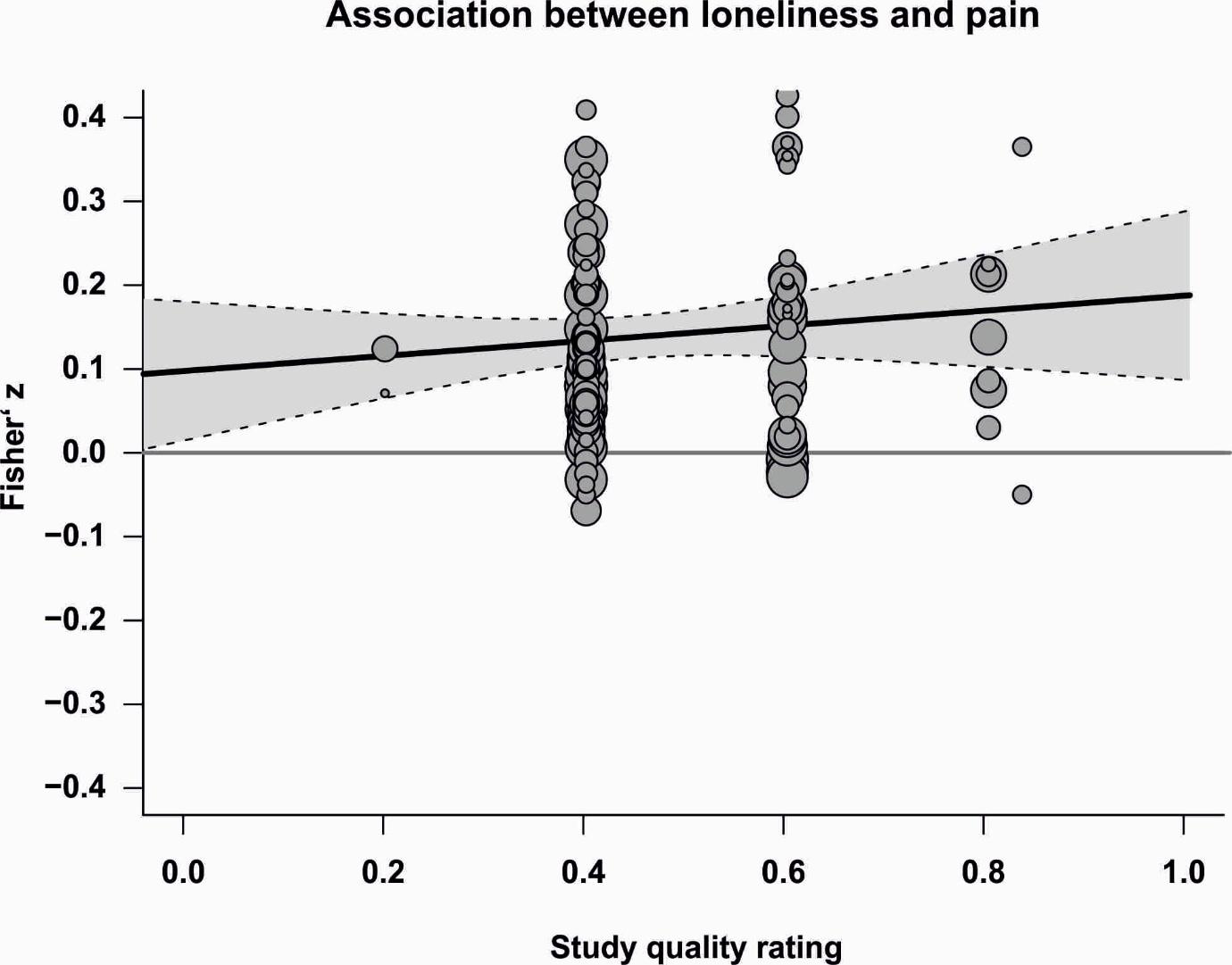

**Supplementary Figure 7.** Meta-regression showing the association between study quality rating and the loneliness–pain effect size. Each dot represents an effect size from an individual dataset, with dot size indicating study precision. The shaded band denotes the 95% confidence interval around the regression line.

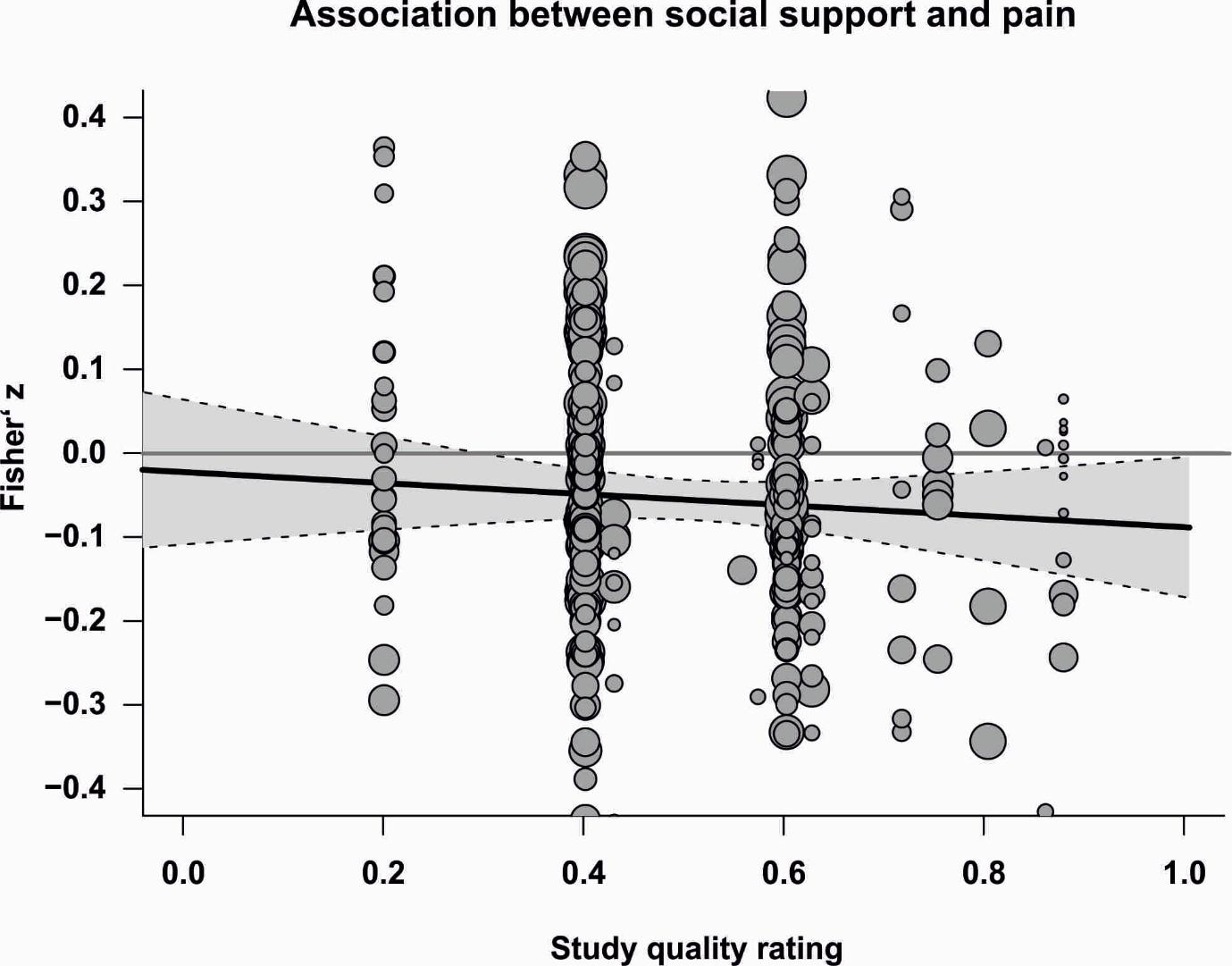

**Supplementary Figure 8.** Meta-regression showing the association between study quality rating and the social support–pain effect size. Each dot represents an effect size from an individual dataset, with dot size indicating study precision. The shaded band denotes the 95% confidence interval around the regression line.

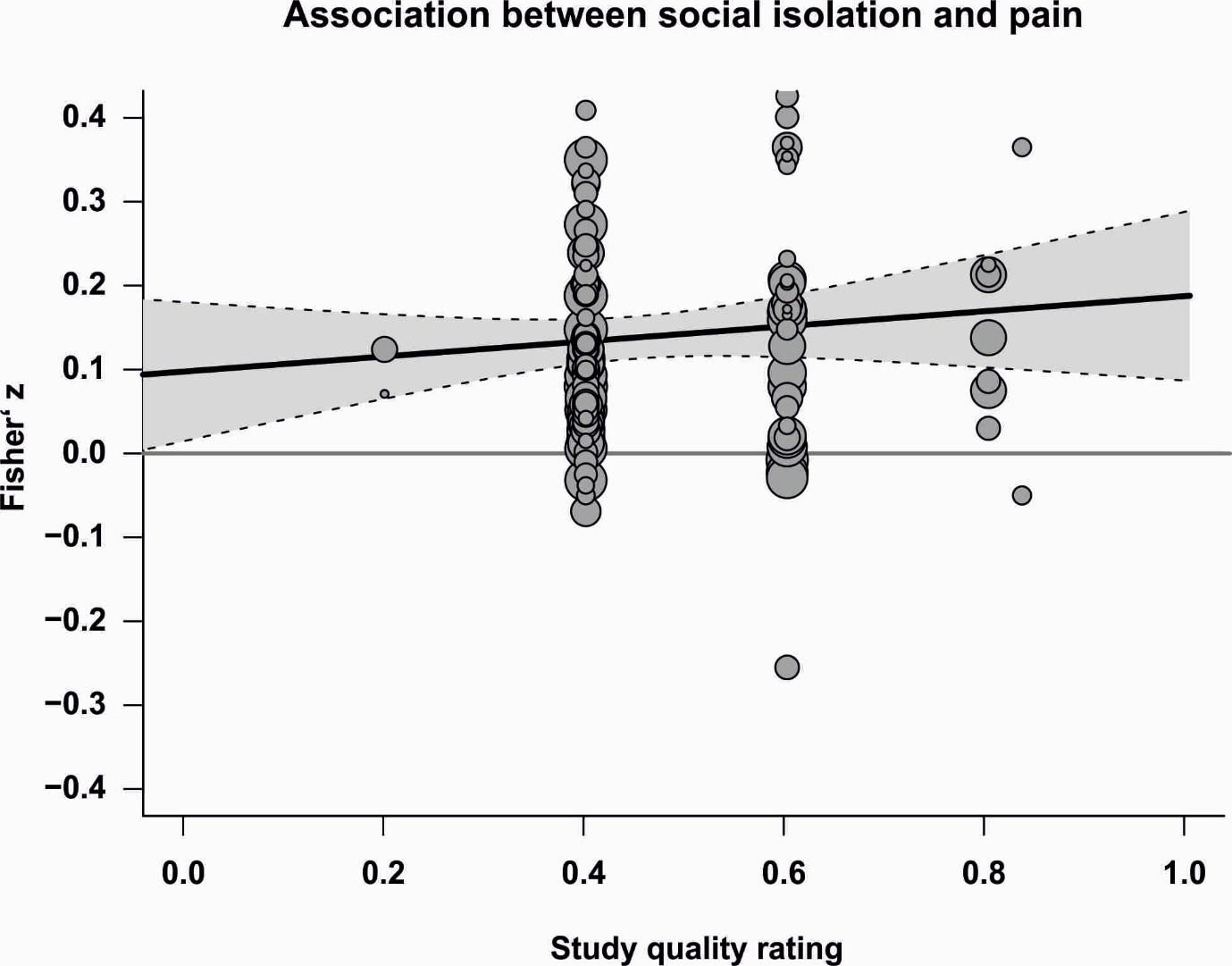

**Supplementary Figure 9.** Meta-regression showing the association between study quality rating and the social isolation–pain effect size. Each dot represents an effect size from an individual dataset, with dot size indicating study precision. The shaded band denotes the 95% confidence interval around the regression line.

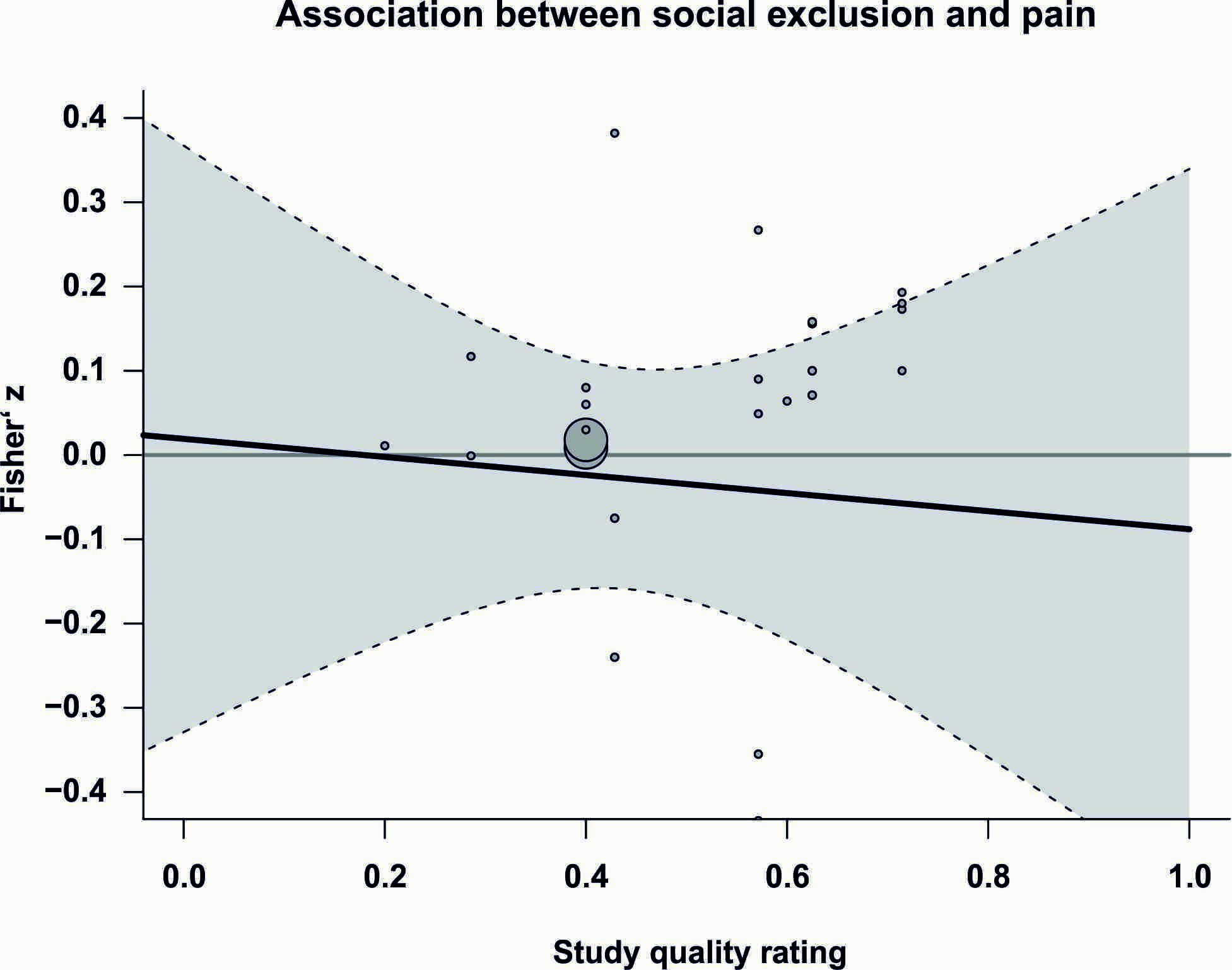

**Supplementary Figure 10.** Meta-regression showing the association between study quality rating and the social exclusion–pain effect size. Each dot represents an effect size from an individual dataset, with dot size indicating study precision. The shaded band denotes the 95% confidence interval around the regression line.

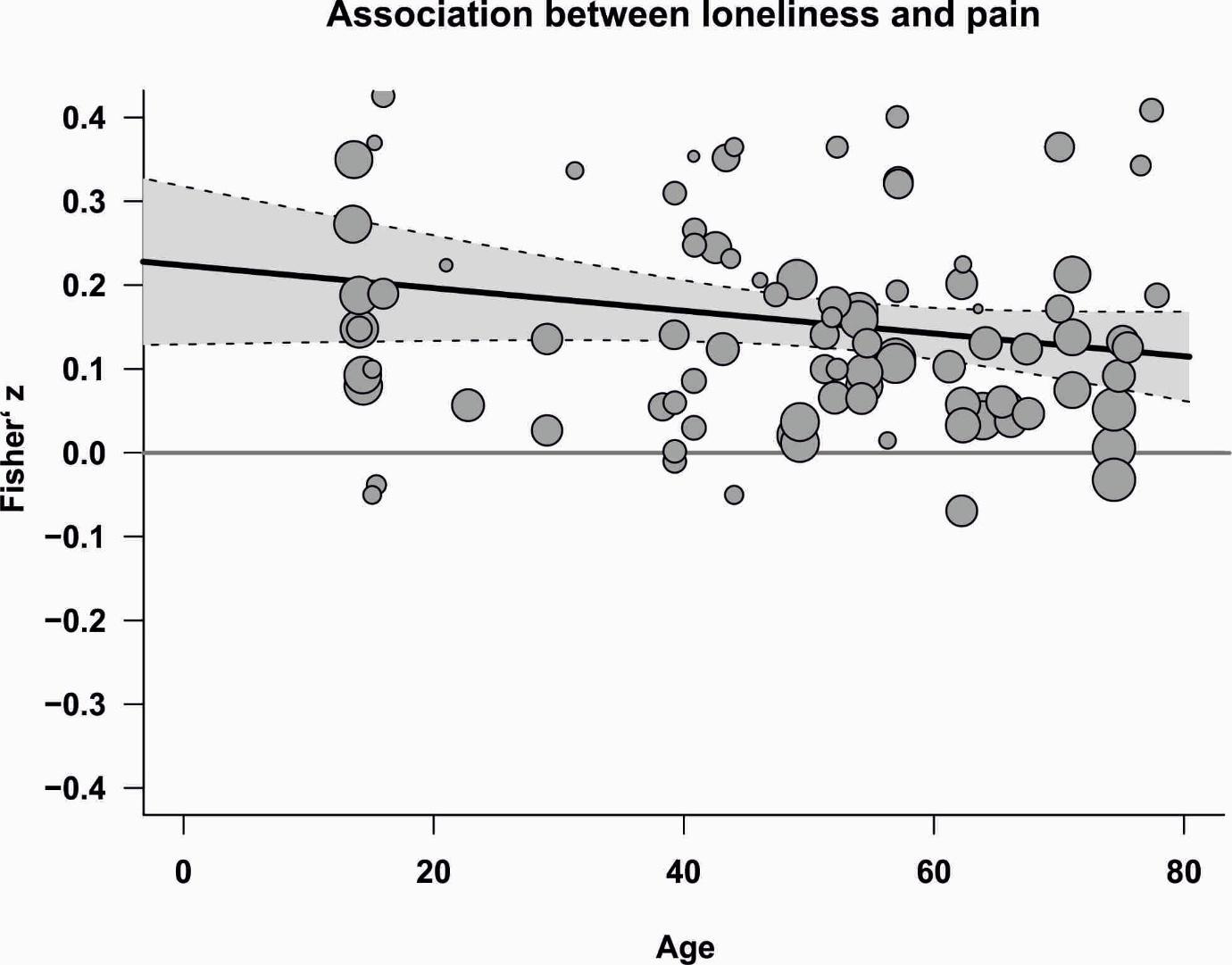

**Supplementary Figure 11.** Meta-regression depicting the association between cohort mean age and the loneliness–pain effect size. Each dot represents an effect size from an individual dataset, with dot size reflecting study precision. The shaded band indicates the 95% confidence interval around the regression line.

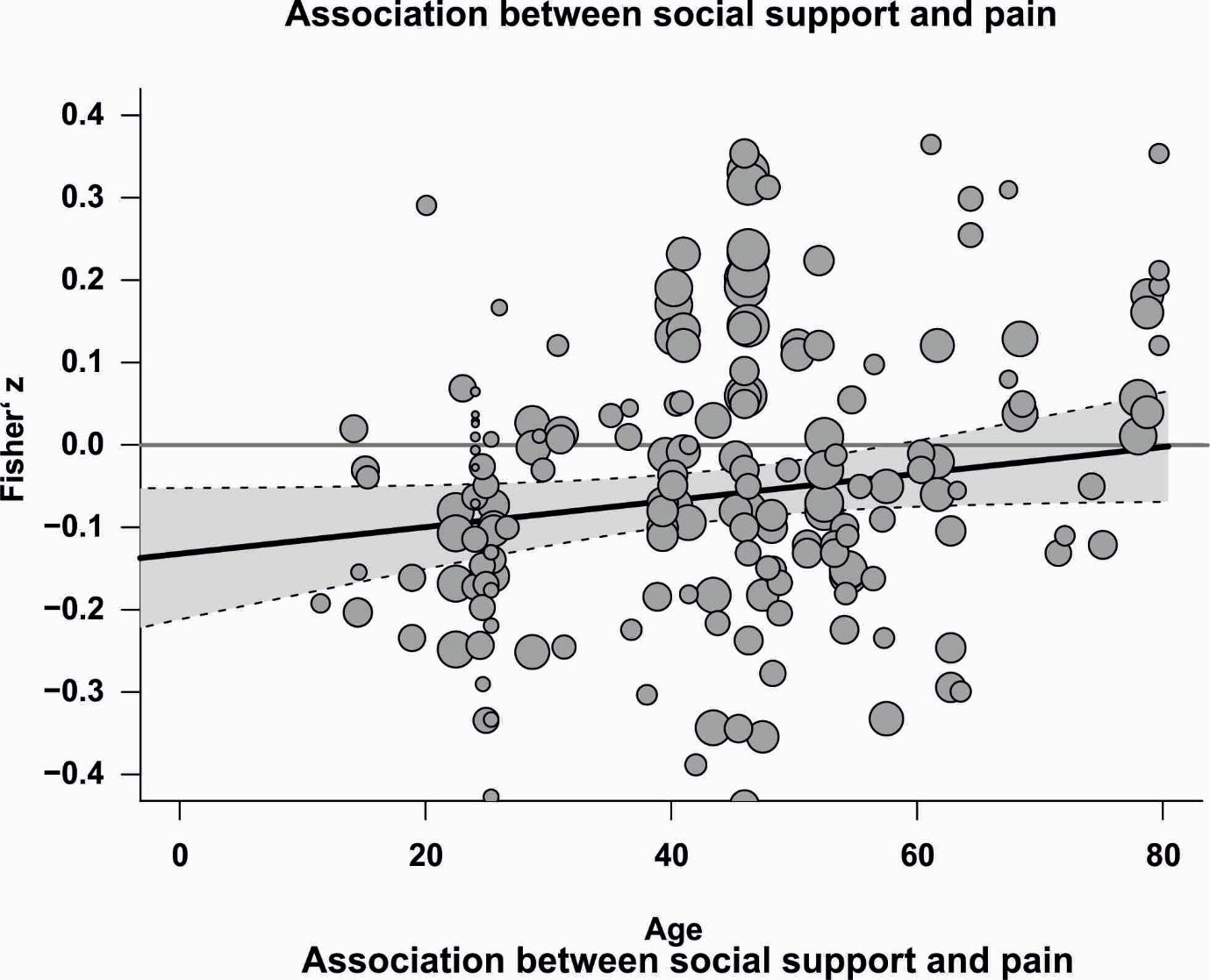

**Supplementary Figure 12.** Meta-regression showing the association between cohort mean age and the social support–pain effect size. Each dot represents an effect size from an individual dataset, with dot size indicating study precision. The shaded band denotes the 95% confidence interval around the regression line.

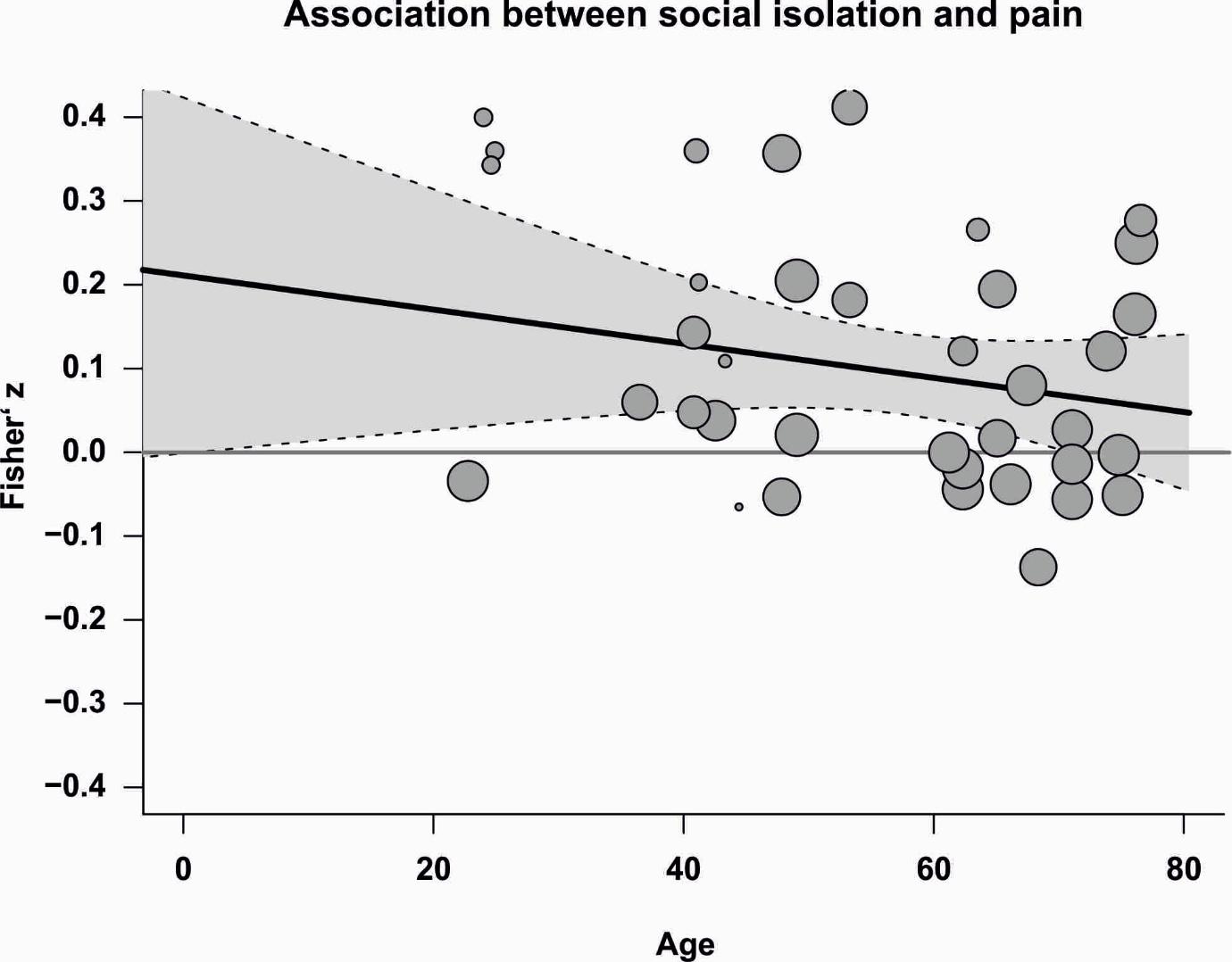

**Supplementary Figure 13.** Meta-regression showing the association between cohort mean age and the social isolation–pain effect size. Each dot represents an effect size from an individual dataset, with dot size indicating study precision. The shaded band denotes the 95% confidence interval around the regression line.

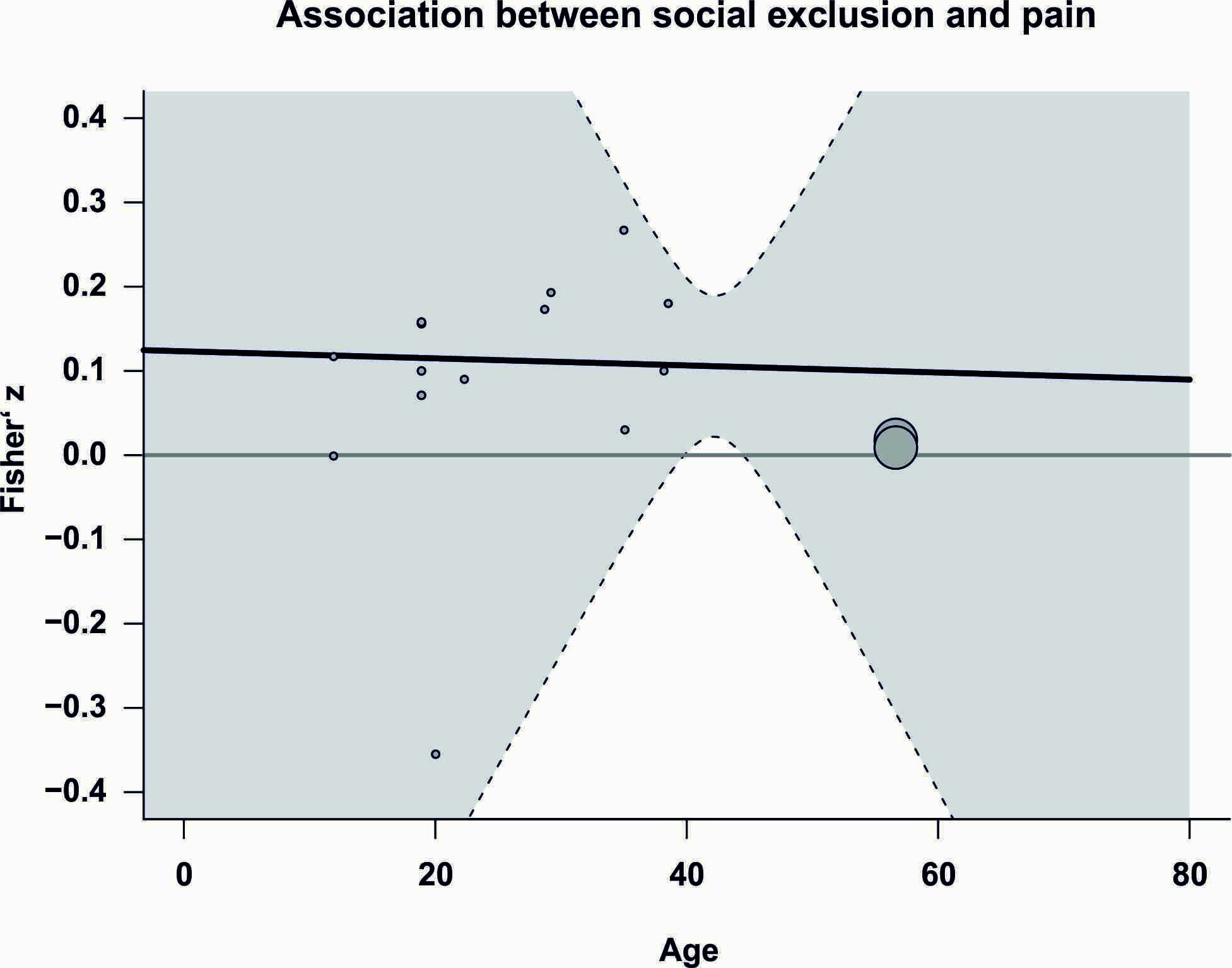

**Supplementary Figure 14.** Meta-regression showing the association between cohort mean age and the social exclusion–pain effect size. Each dot represents an effect size from an individual dataset, with dot size indicating study precision. The shaded band denotes the 95% confidence interval around the regression line.

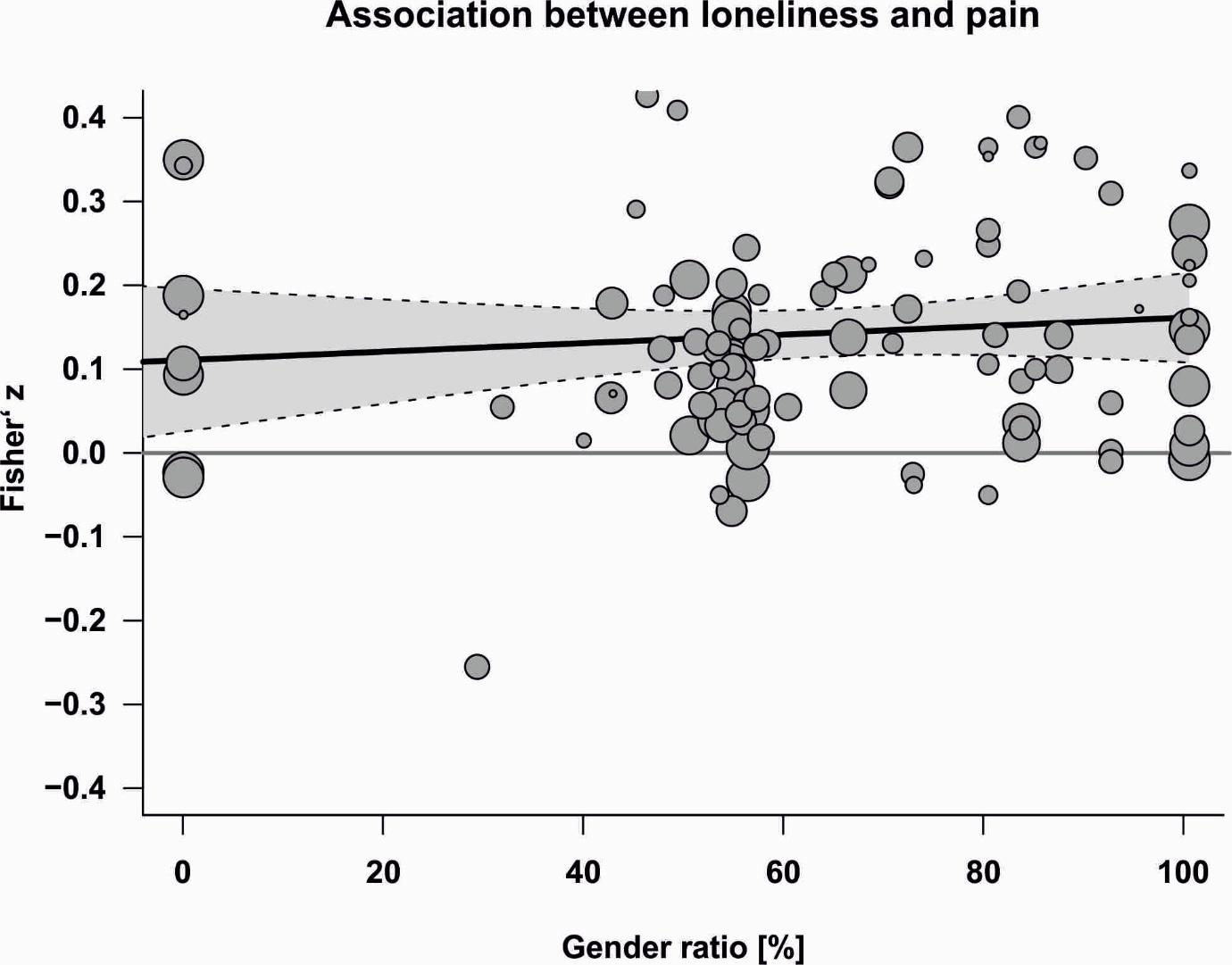

**Supplementary Figure 15.** Meta-regression showing the association between cohort gender ratio (proportion of women) and the loneliness–pain effect size. Each dot represents an effect size from an individual dataset, with dot size indicating study precision. The shaded band denotes the 95% confidence interval around the regression line.

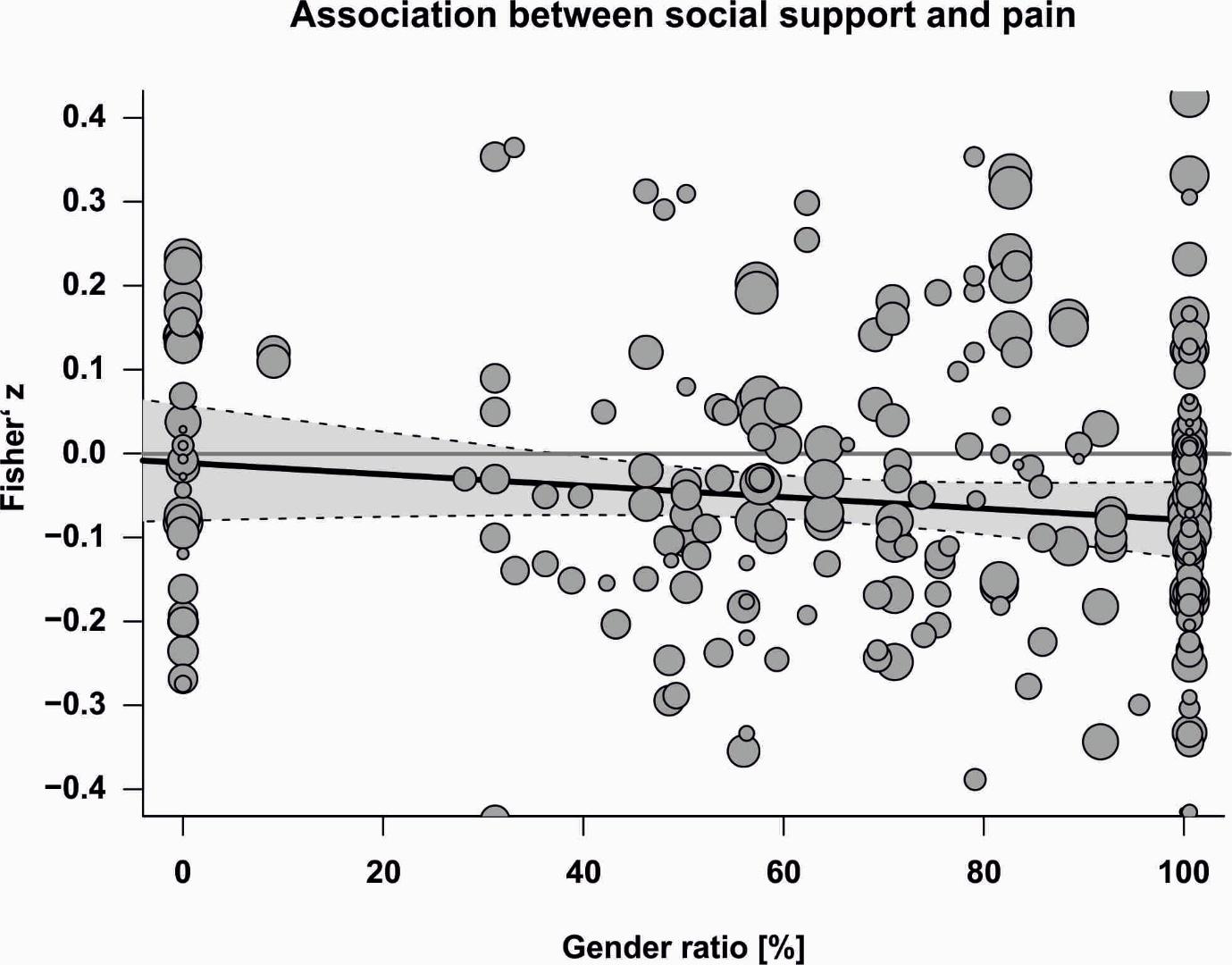

**Supplementary Figure 16.** Meta-regression showing the association between cohort gender ratio (proportion of women) and the social support–pain effect size. Each dot represents an effect size from an individual dataset, with dot size indicating study precision. The shaded band denotes the 95% confidence interval around the regression line.

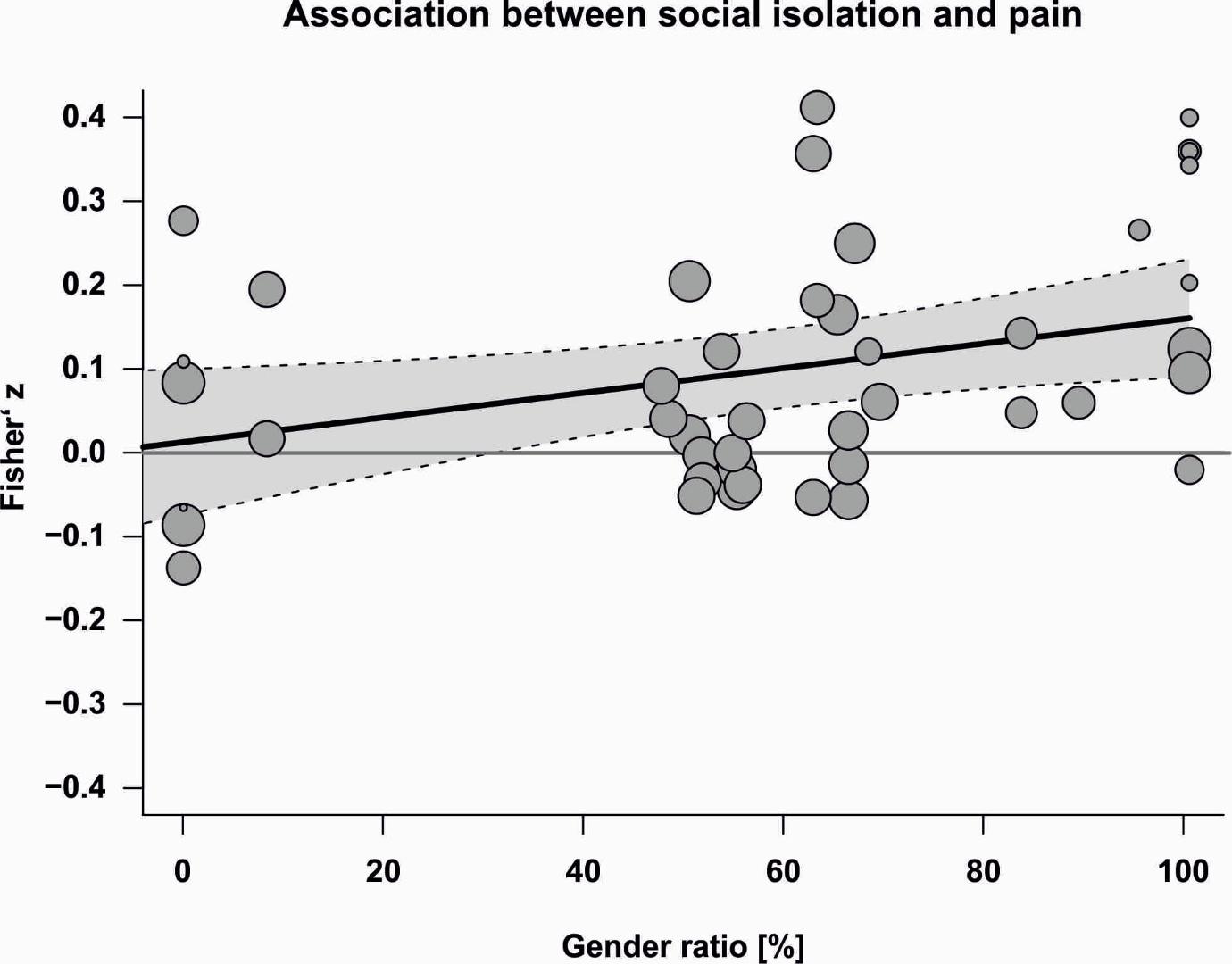

**Supplementary Figure 17.** Meta-regression showing the association between cohort gender ratio (proportion of women) and the social isolation–pain effect size. Each dot represents an effect size from an individual dataset, with dot size indicating study precision. The shaded band denotes the 95% confidence interval around the regression line.

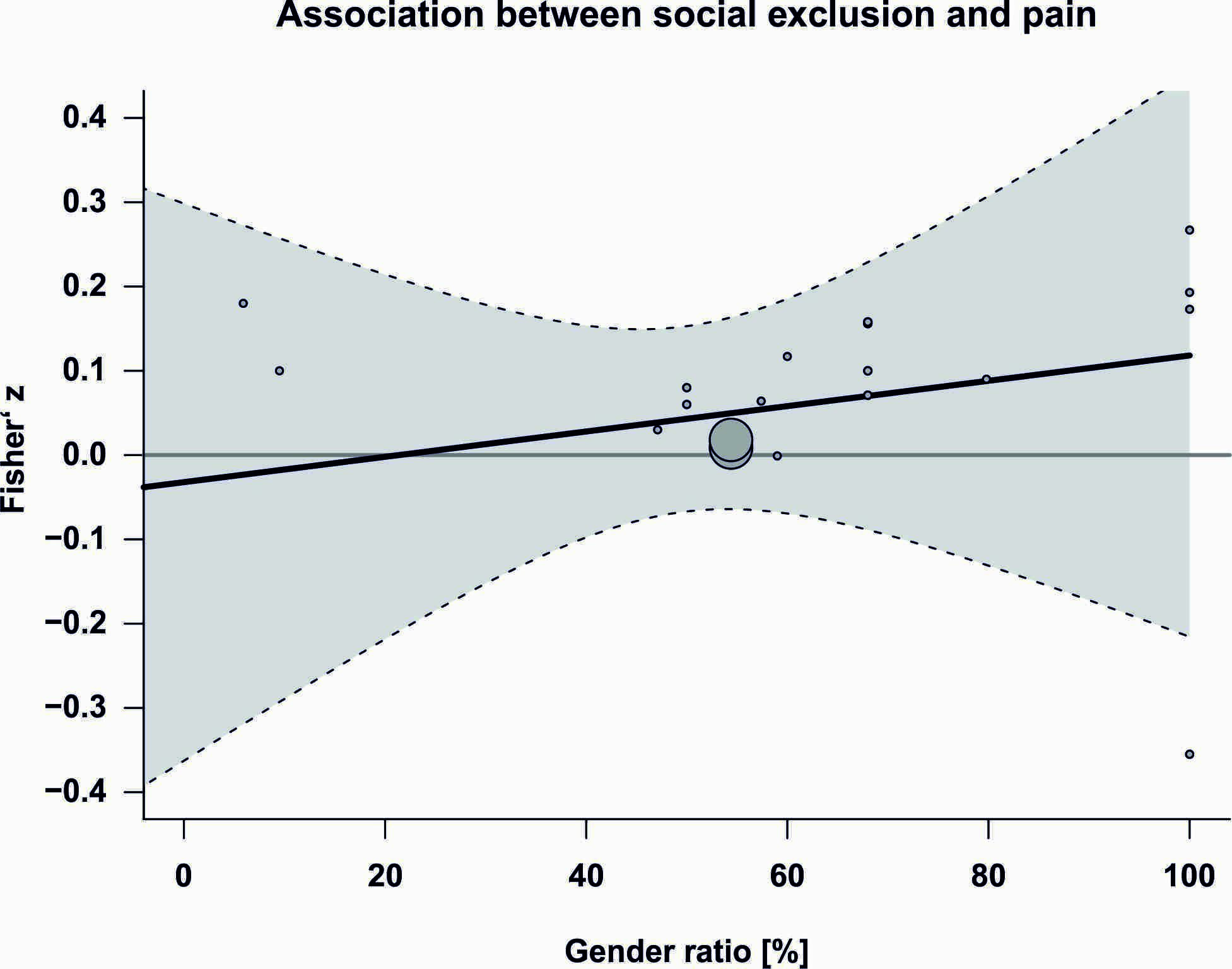

**Supplementary Figure 18.** Meta-regression showing the association between cohort gender ratio (proportion of women) and the social exclusion–pain effect size. Each dot represents an effect size from an individual dataset, with dot size indicating study precision. The shaded band denotes the 95% confidence interval around the regression line.

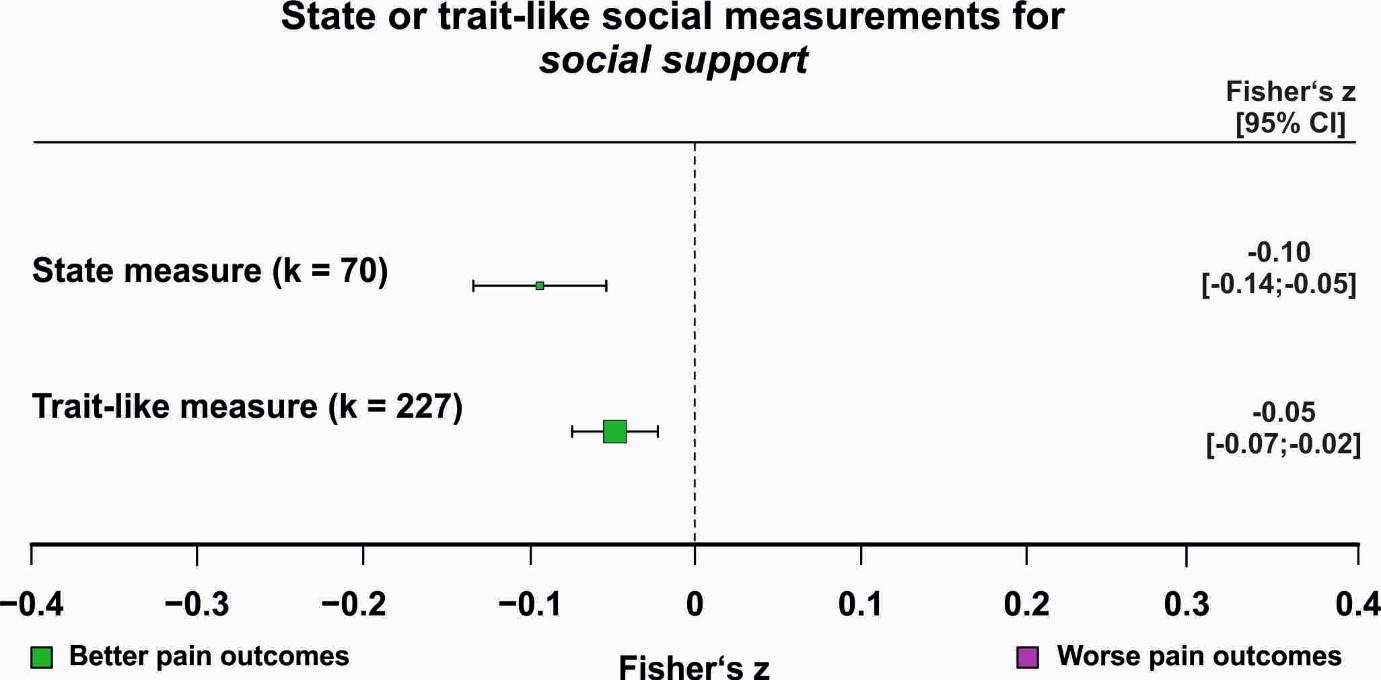

**Supplementary Figure 19.** Association between pain and social support for state versus trait-like social support assessments. The number of contributing effect sizes (k) for each subgroup is shown; only subgroups with at least ten effect sizes were analysed. Pooled effect estimates with 95% confidence intervals are reported numerically. Purple squares indicate significantly worse pain outcomes, green squares indicate significantly better pain outcomes, and grey squares indicate non-significant associations. Significant post hoc comparisons (two-sided *t* tests) are indicated. The corresponding orchard plot is provided in Supplementary Figure 29.

* = *p* < .05, ** = *p* < .01, *** = *p* < .001

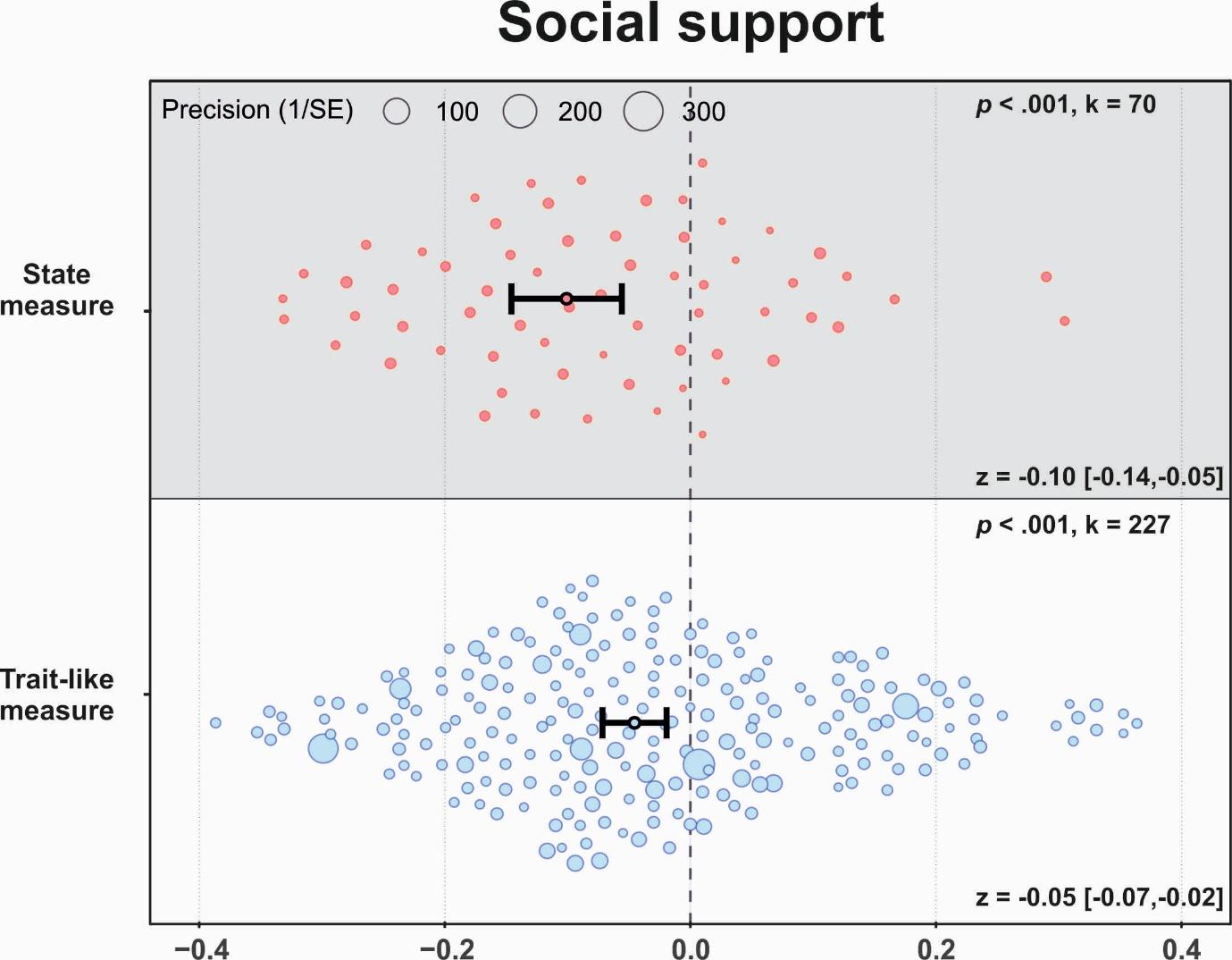

**Supplementary Figure 20.** Orchard plot illustrating associations between social support and pain for state versus trait-like social assessments. Each dot represents a cohort-level effect size, with dot size reflecting precision. The number of included effect sizes (k) is shown in the upper right of each panel. Pooled effects with 95% confidence intervals are displayed in the lower right and indicated by black dots and error bars.

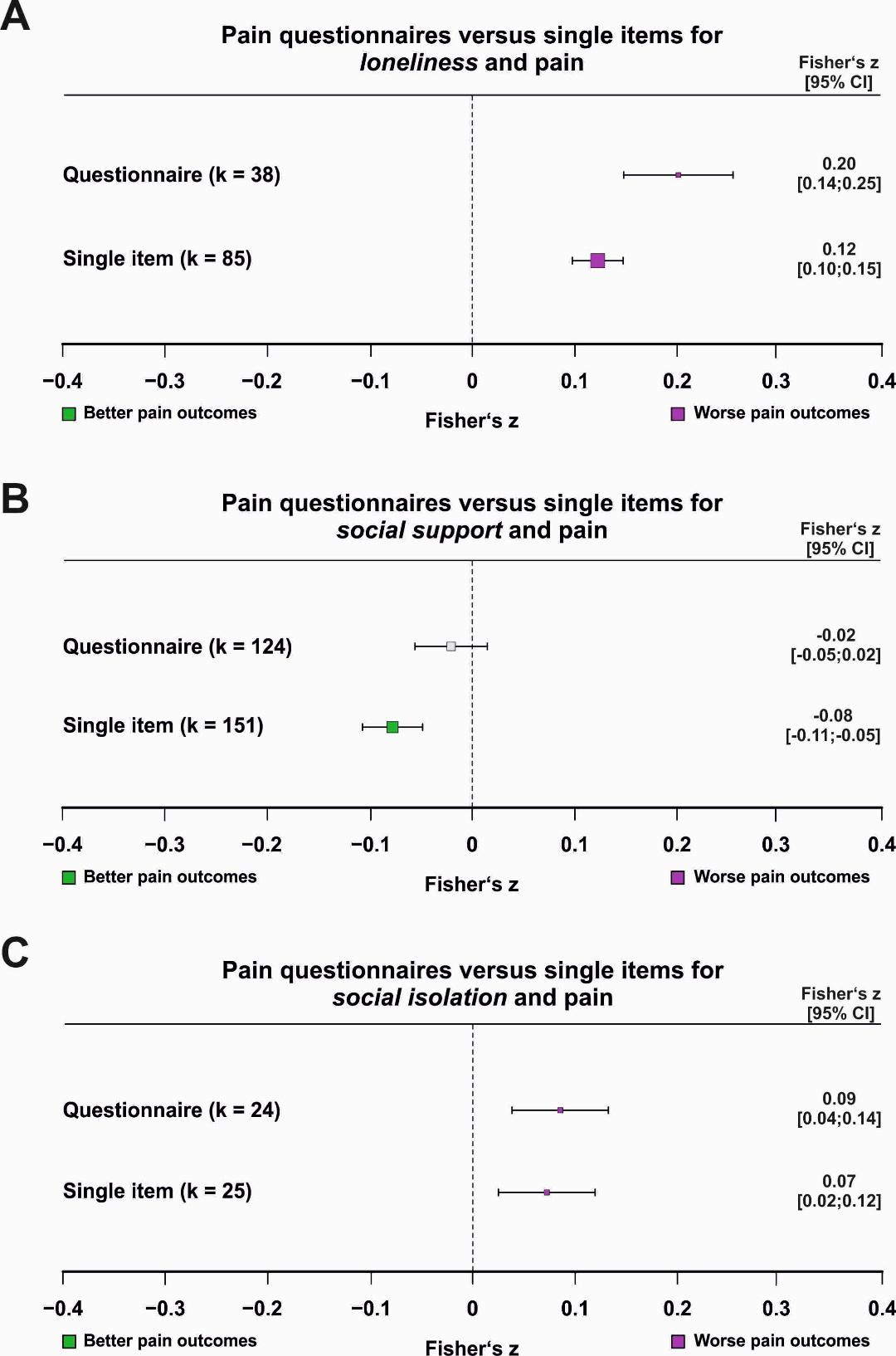

**Supplementary Figure 21.** Associations between pain and **(A)** loneliness, **(B)** social support, and **(C)** social isolation for single-item versus multi-item pain assessments. The number of contributing effect sizes (k) for each subgroup is shown; only subgroups with at least ten effect sizes were analysed. Pooled effect estimates with 95% confidence intervals are reported numerically. Purple squares indicate significantly worse pain outcomes, green squares indicate significantly better pain outcomes, and grey squares indicate non-significant associations. Significant post hoc comparisons (two-sided *t* tests) are indicated. The corresponding orchard plot is provided in Supplementary Figure 31.* = *p* < .05, ** = *p* < .01, *** = *p* < .001

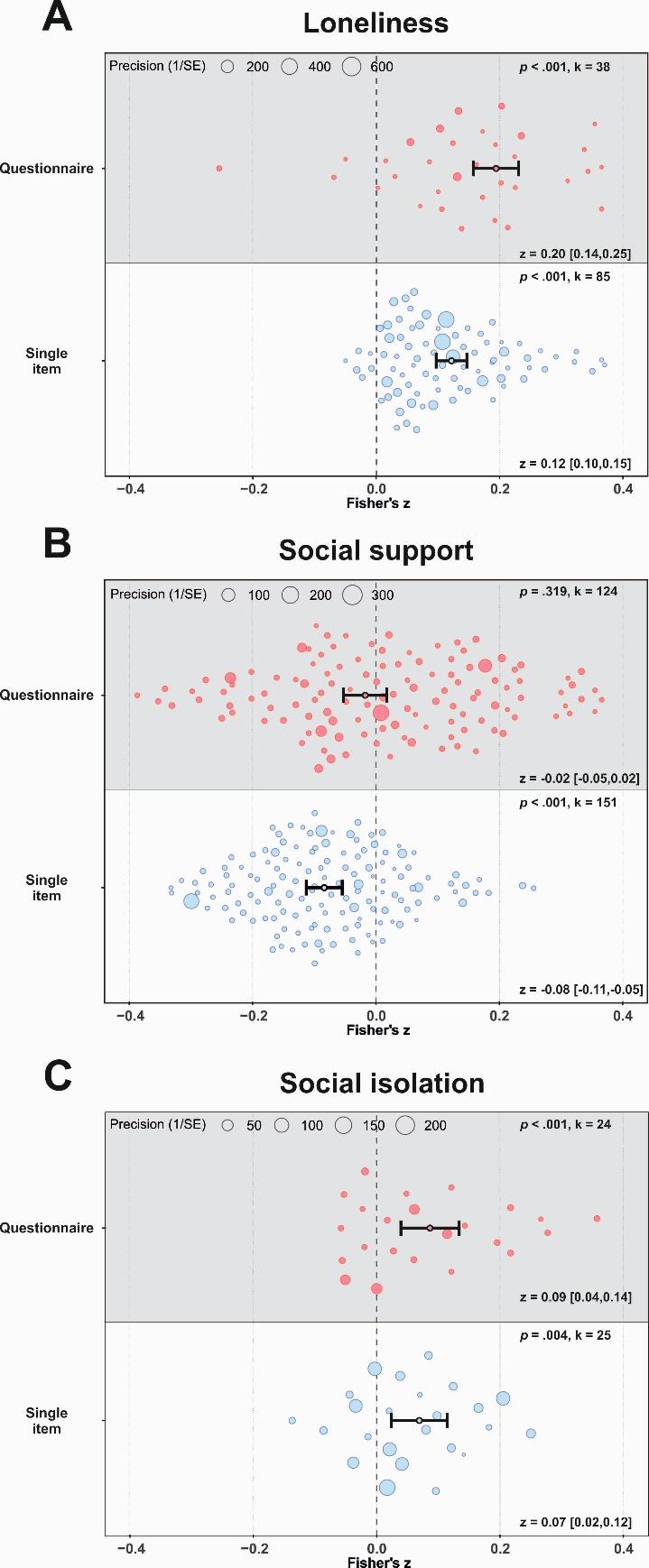

**Supplementary Figure 22.** Orchard plot illustrating associations between **(A)** loneliness, **(B)** social support, and **(C)** social isolation and pain for single-item versus multi-item pain assessments. Each dot represents a cohort-level effect size, with dot size reflecting precision. The number of included effect sizes (k) is shown in the upper right of each panel. Pooled effects with 95% confidence intervals are displayed in the lower right and indicated by black dots and error bars.

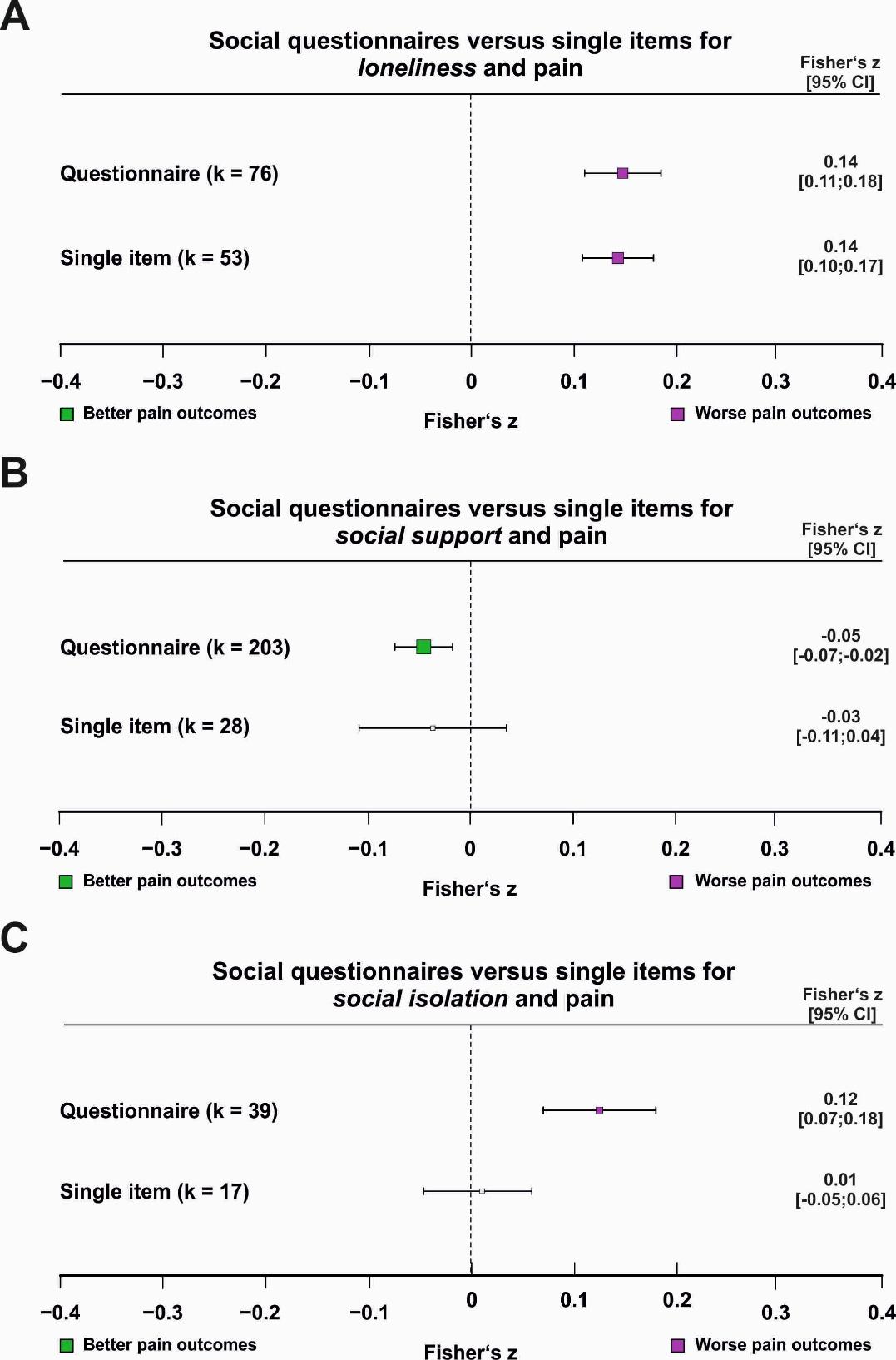

**Supplementary Figure 23.** Associations between pain and **(A)** loneliness, **(B)** social support, and **(C)** social isolation for single-item versus multi-item assessments of the respective social constructs. The number of contributing effect sizes (k) for each subgroup is shown; only subgroups with at least ten effect sizes were analysed. Pooled effect estimates with 95% confidence intervals are reported numerically. Purple squares indicate significantly worse pain outcomes, green squares indicate significantly better pain outcomes, and grey squares indicate non-significant associations. Significant post hoc comparisons (two-sided *t* tests) are indicated. The corresponding orchard plot is provided in Supplementary Figure 33. * = p < .05, ** = p < .01, *** = p < .001

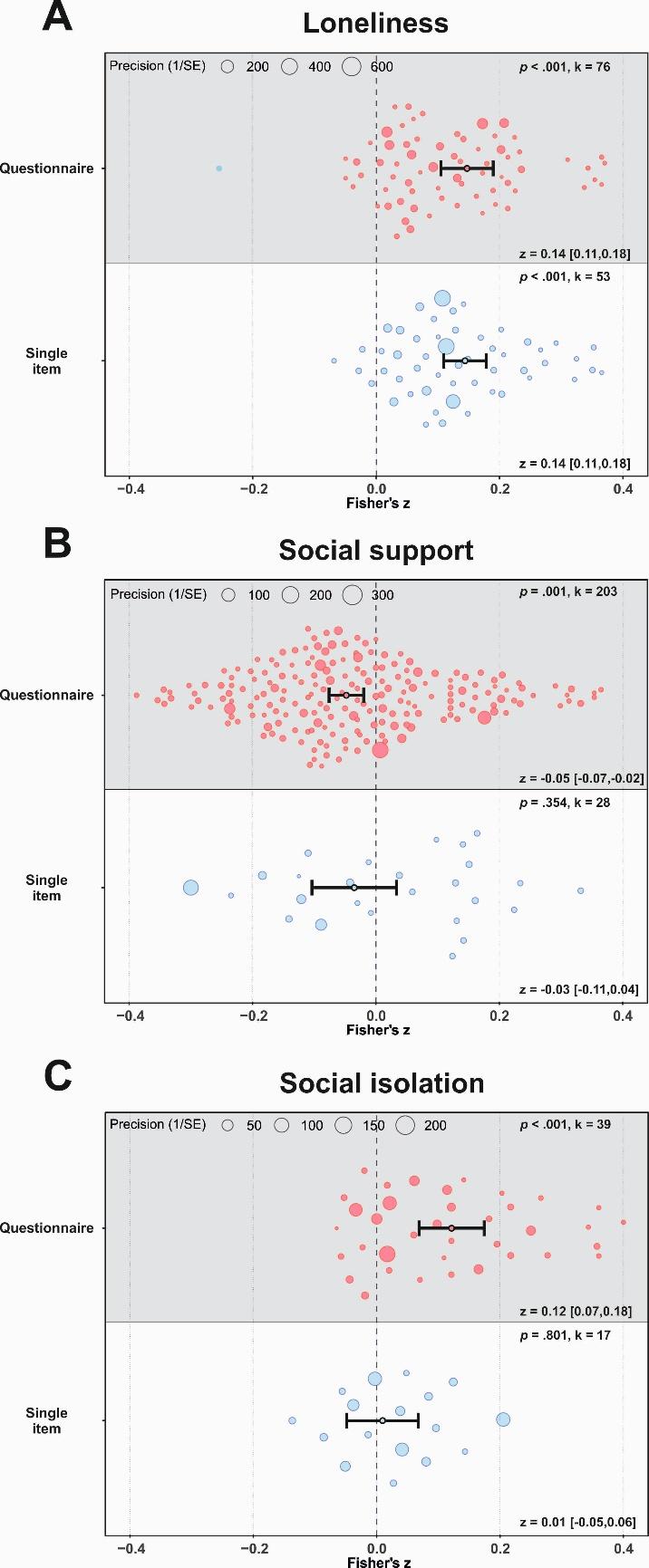

**Supplementary Figure 24.** Orchard plot illustrating associations between **(A)** loneliness, **(B)** social support, and **(C)** social isolation and pain for single-item versus multi-item assessments of the respective social constructs. Each dot represents a cohort-level effect size, with dot size reflecting precision. The number of included effect sizes (k) is shown in the upper right of each panel. Pooled effects with 95% confidence intervals are displayed in the lower right and indicated by black dots and error bars.

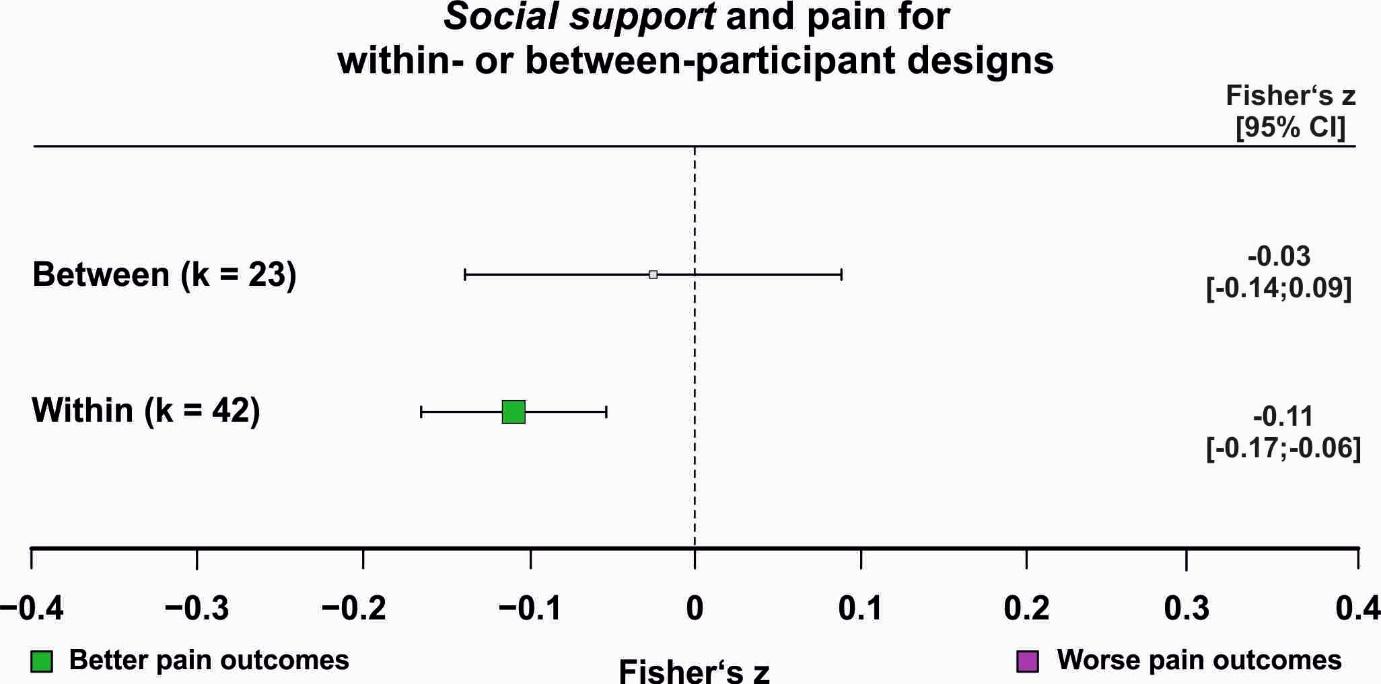

**Supplementary Figure 25.** Association between pain and social support in within-participant versus between-participant study designs. The number of contributing effect sizes (k) for each design is shown; only designs with at least ten effect sizes were analysed. Pooled effect estimates with 95% confidence intervals are reported numerically. Purple squares indicate significantly worse pain outcomes, green squares indicate significantly better pain outcomes, and grey squares indicate non-significant associations. Significant post hoc comparisons (two-sided *t* tests) are indicated. The corresponding orchard plot is provided in Supplementary Figure 35.* = p < .05, ** = p < .01, *** = p < .001

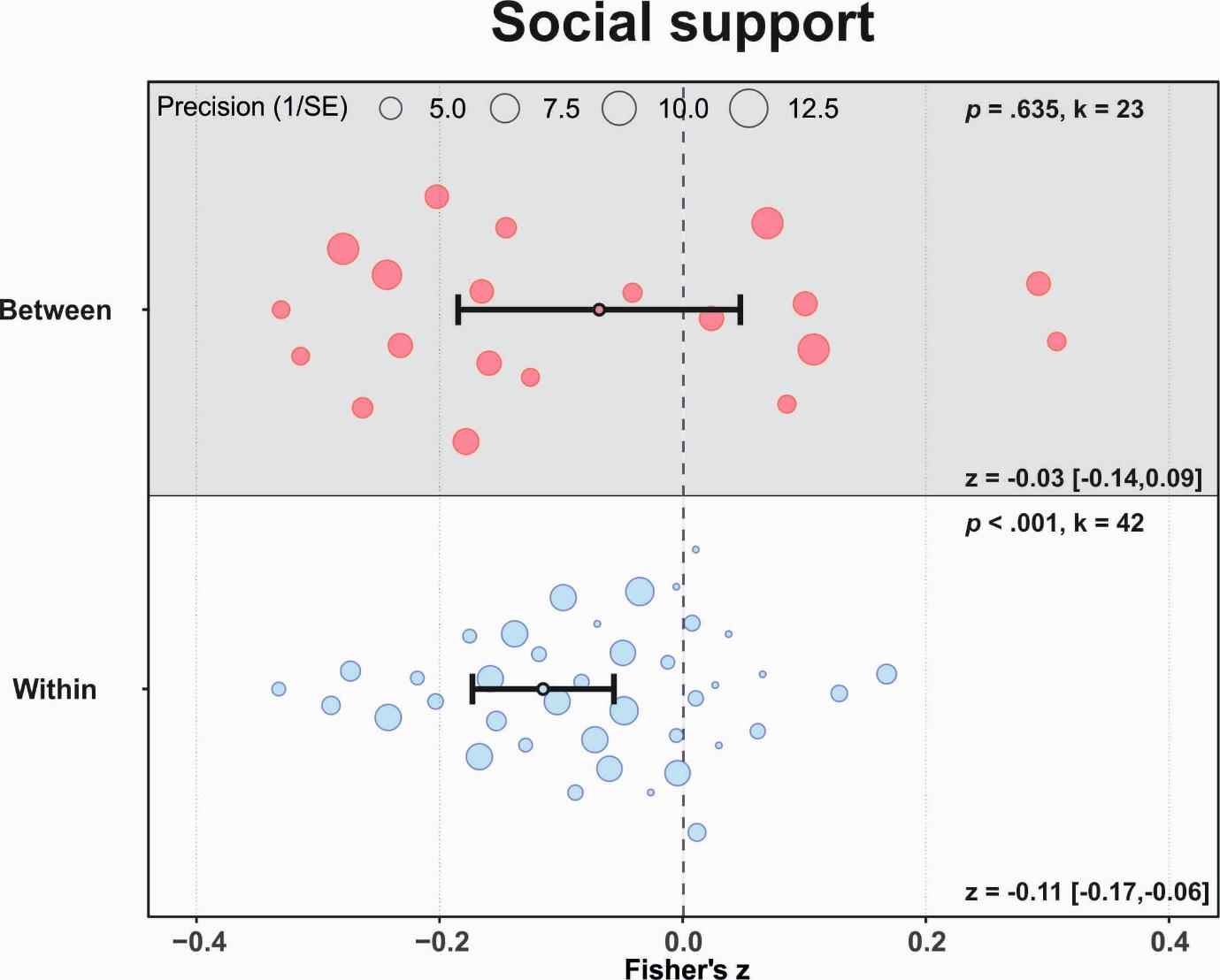

**Supplementary Figure 26.** Orchard plot illustrating associations between social support and pain in within-participant versus between-participant study designs. Each dot represents a cohort-level effect size, with dot size reflecting precision. The number of included effect sizes (k) is shown in the upper right of each panel. Pooled effects with 95% confidence intervals are displayed in the lower right and indicated by black dots and error bars.

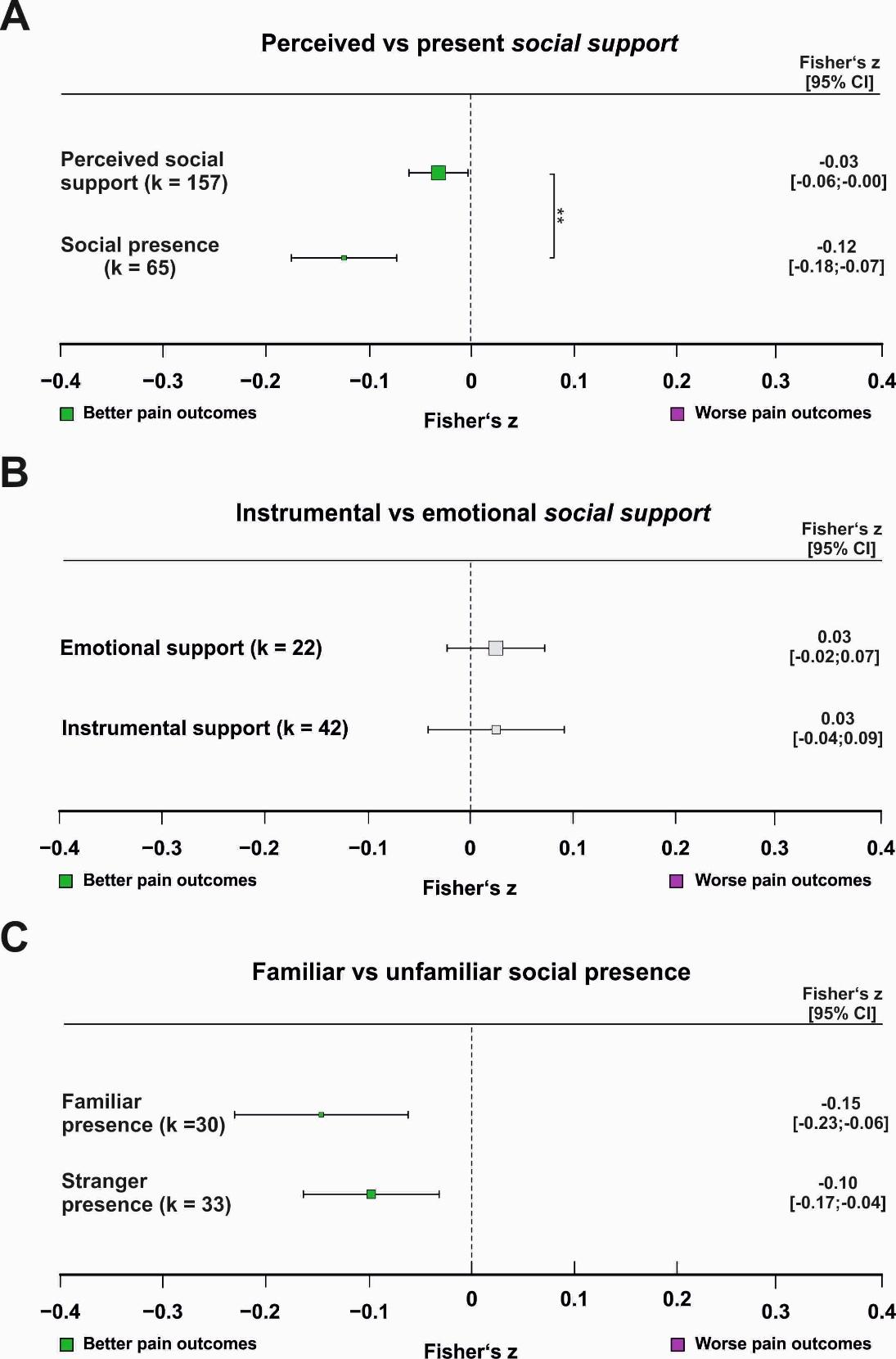

**Supplementary Figure 27.** Associations between pain and social support across different operationalisations: **(A)** perceived versus present support, **(B)** emotional versus instrumental support, and **(C)** presence of a familiar versus unfamiliar individual. The number of contributing effect sizes (k) for each subgroup is shown; only subgroups with at least ten effect sizes were analysed. Pooled effect estimates with 95% confidence intervals are reported numerically. Purple squares indicate significantly worse pain outcomes, green squares indicate significantly better pain outcomes, and grey squares indicate non-significant associations. Significant post hoc comparisons (two-sided *t* tests) are indicated. The corresponding orchard plot is provided in Supplementary Figure 26.

* = *p* < .05, ** = *p* < .01, *** = *p* < .001

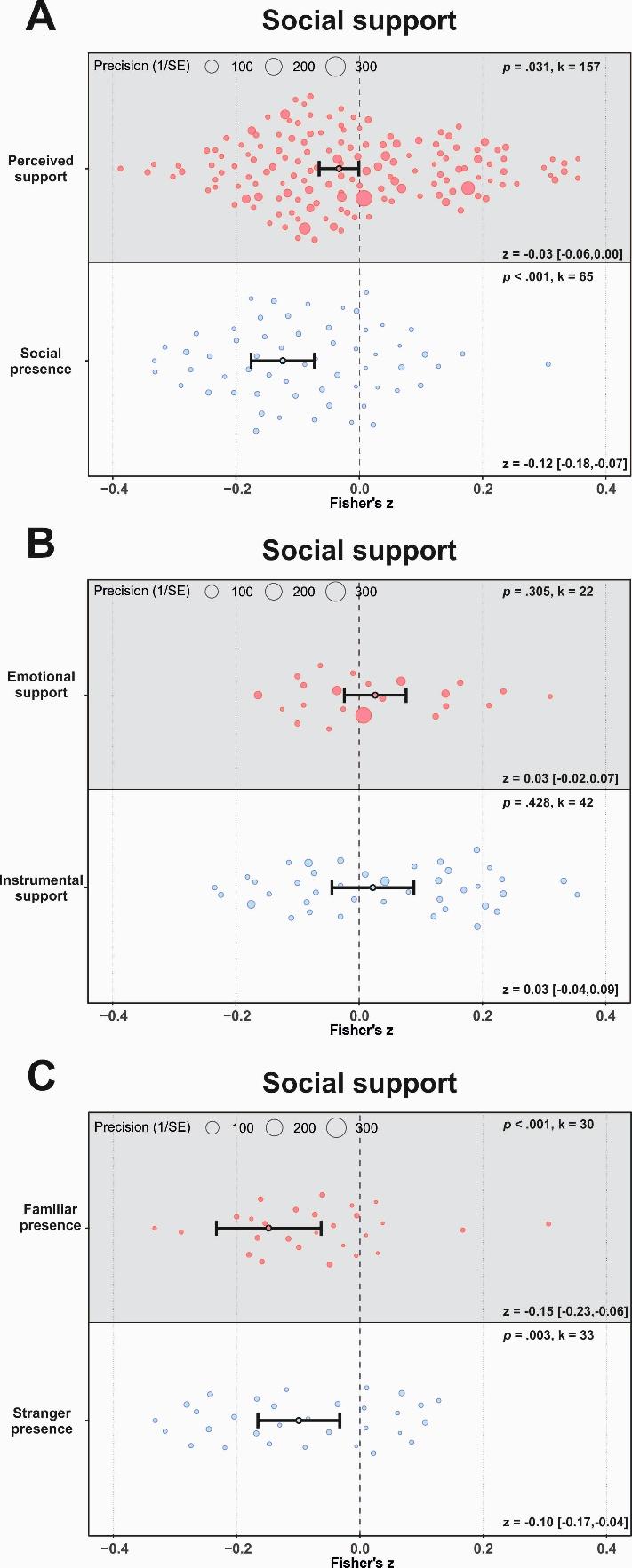

**Supplementary Figure 28.** Orchard plot illustrating associations between social support and pain across different operationalisations: **(A)** perceived support versus social presence, **(B)** emotional versus instrumental support, and **(C)** familiar versus unfamiliar presence. Each dot represents a cohort-level effect size, with dot size reflecting precision. The number of included effect sizes (k) is shown in the upper right of each panel. Pooled effects with 95% confidence intervals are displayed in the lower right and indicated by black dots and error bars.

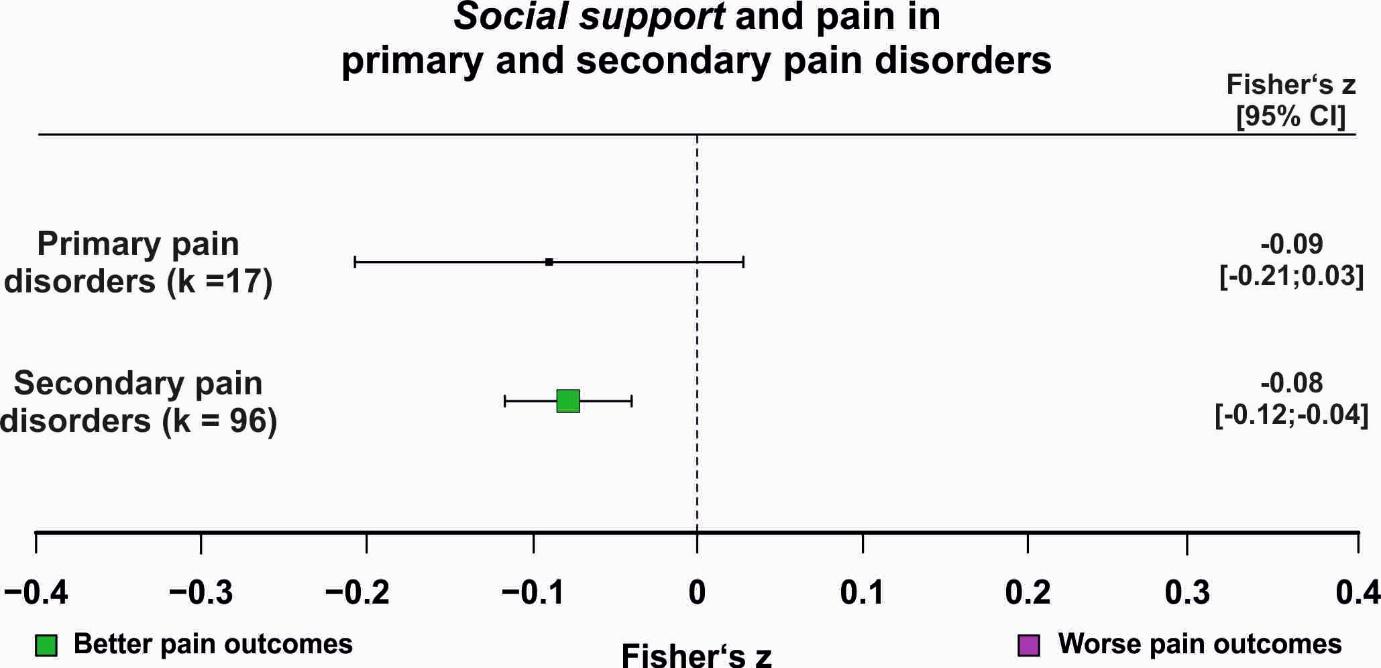

**Supplementary Figure 29.** Association between pain and social support in primary and secondary pain disorders. The number of contributing effect sizes (k) for each subgroup is shown; only subgroups with at least ten effect sizes were analysed. Pooled effect estimates with 95% confidence intervals are reported numerically. Purple squares indicate significantly worse pain outcomes, green squares indicate significantly better pain outcomes, and grey squares indicate non-significant associations. Significant post hoc comparisons (two-sided *t* tests) are indicated. The corresponding orchard plot is provided in Supplementary Figure 30. * = *p* < .05, ** = *p* < .01, *** = *p* < .001

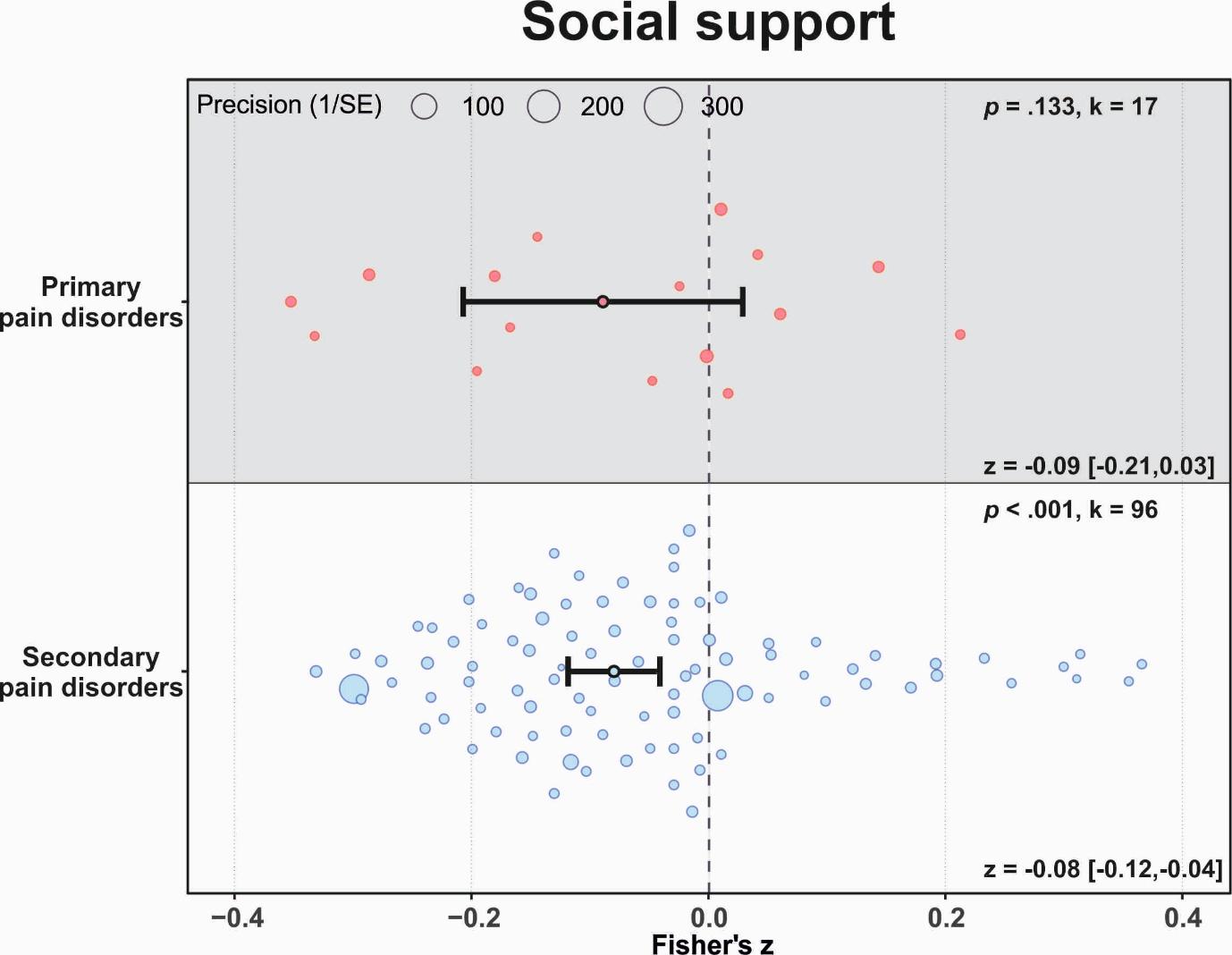

**Supplementary Figure 30.** Orchard plot illustrating associations between social support and pain in primary and secondary pain disorders. Each dot represents a cohort-level effect size, with dot size reflecting precision. The number of included effect sizes (k) is shown in the upper right of each panel. Pooled effects with 95% confidence intervals are displayed in the lower right and indicated by black dots and error bars.

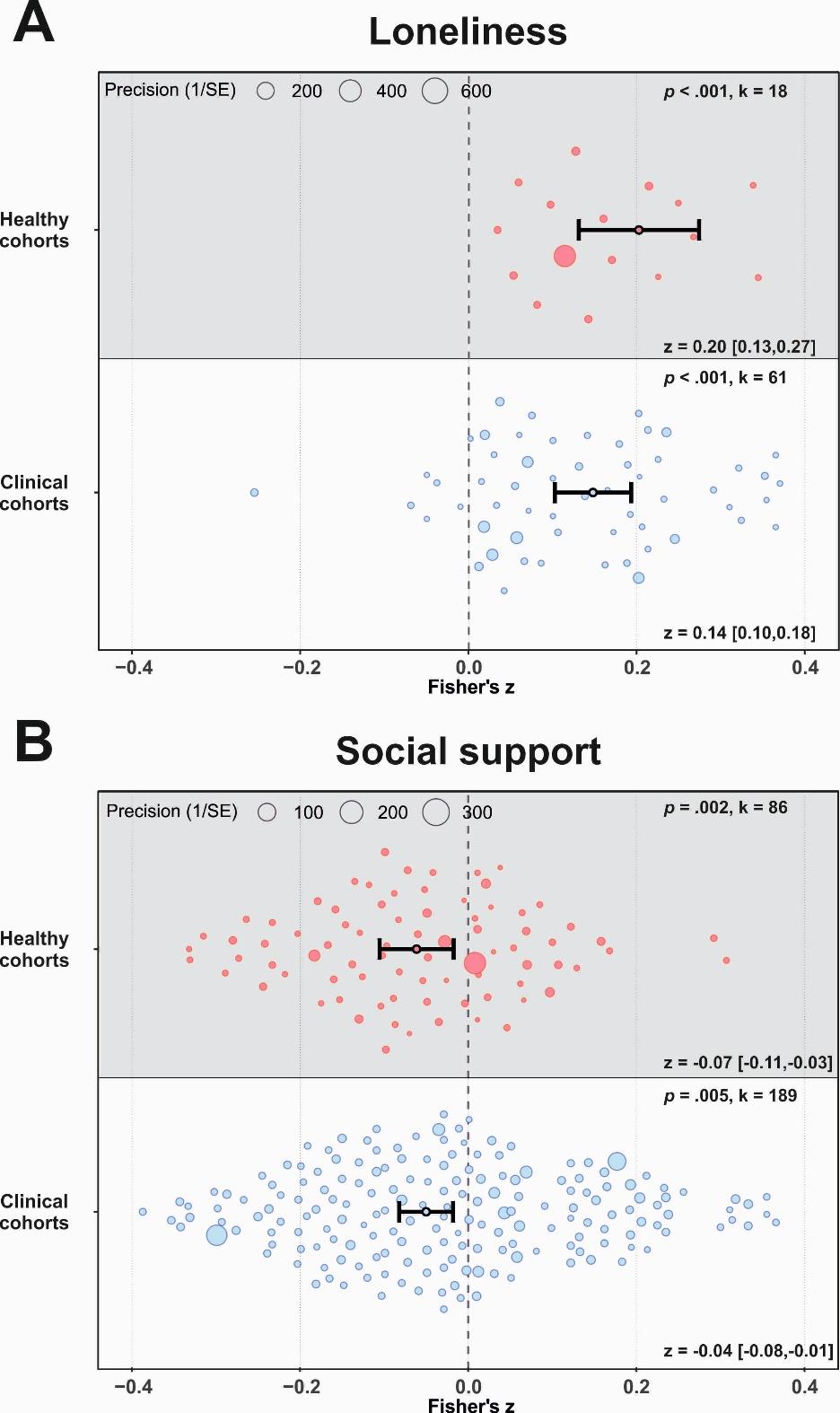

**Supplementary Figure 31.** Orchard plot illustrating associations between loneliness and social support and pain in clinical and non-clinical cohorts. Each dot represents a cohort-level effect size, with dot size reflecting precision. The number of included effect sizes (k) is shown in the upper right of each panel. Pooled effects with 95% confidence intervals are displayed in the lower right and indicated by black dots and error bars.

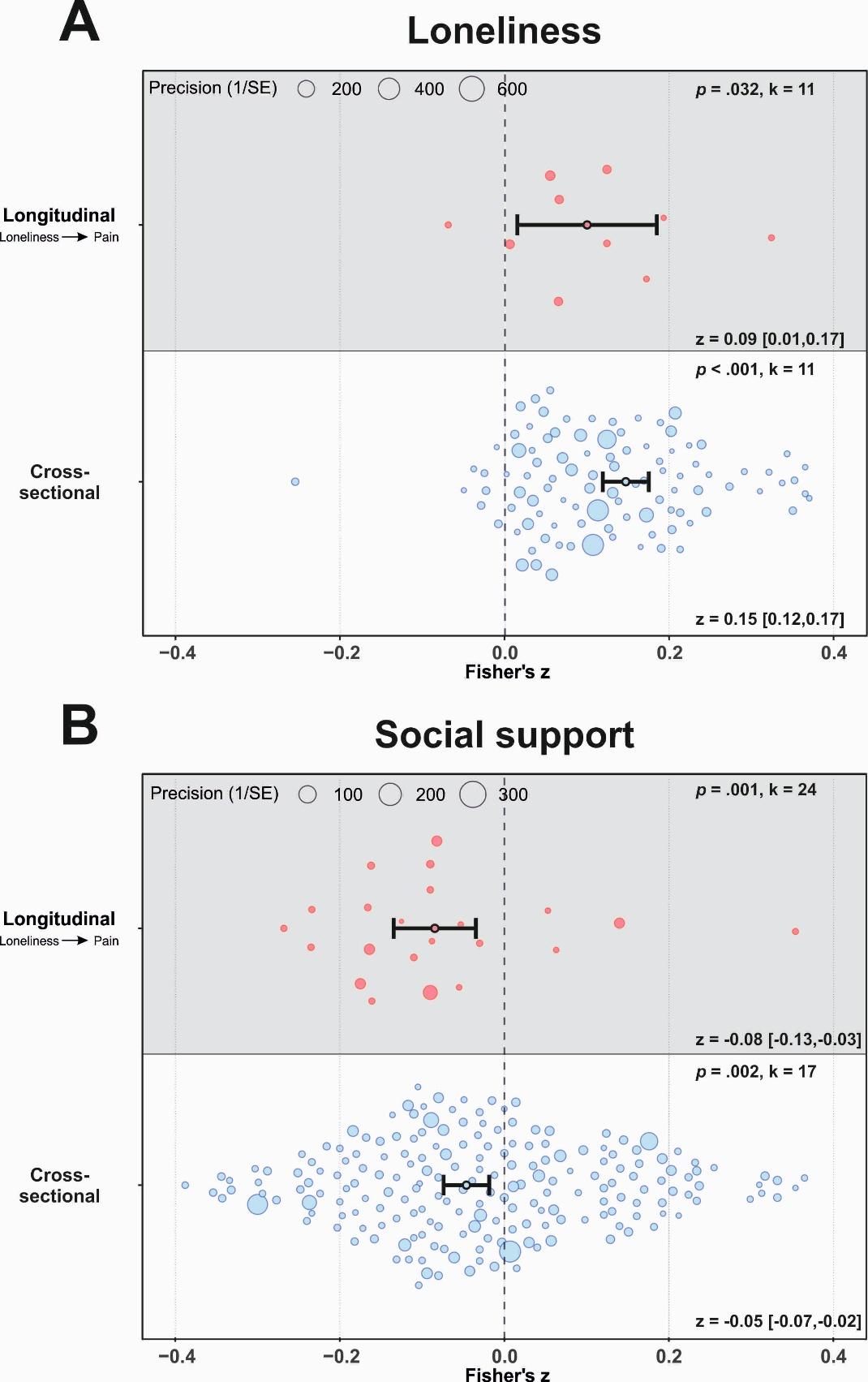

**Supplementary Figure 32.** Orchard plot illustrating associations between loneliness and social support and pain in cross-sectional and longitudinal datasets. Each dot represents a cohort-level effect size, with dot size reflecting precision. The number of included effect sizes (k) is shown in the upper right of each panel. Pooled effects with 95% confidence intervals are displayed in the lower right and indicated by black dots and error bars.

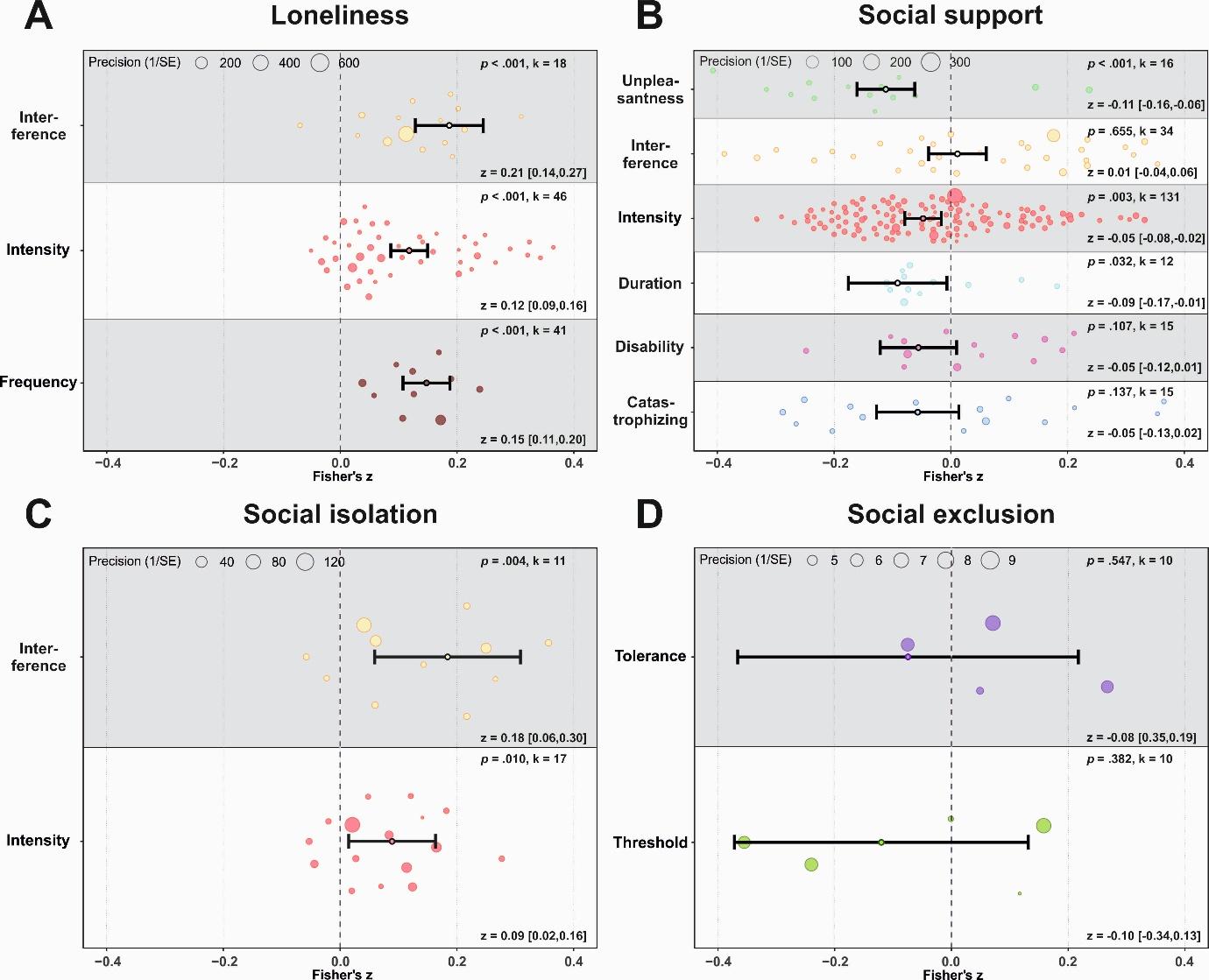

**Supplementary Figure 33.** Orchard plot illustrating associations between dimensions of social connectedness and specific pain outcomes. Each dot represents a cohort-level effect size, with dot size reflecting precision. The number of included effect sizes (k) is shown in the upper right of each panel. Pooled effects with 95% confidence intervals are displayed in the lower right and indicated by black dots and error bars.

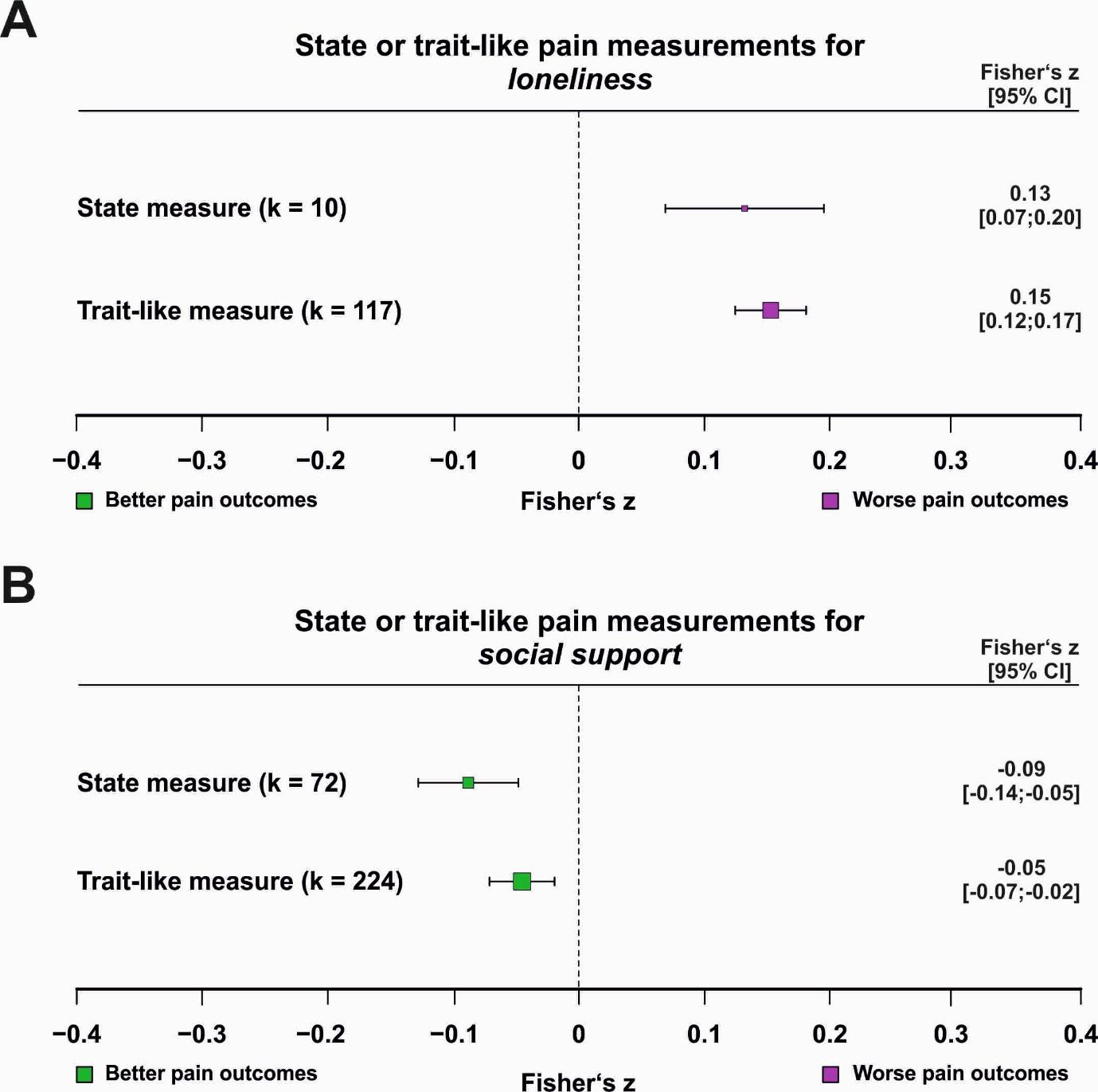

**Supplementary Figure 34.** Associations between pain and **(A)** loneliness and **(B)** social support for state versus trait-like pain assessments. The number of contributing effect sizes (k) for each subgroup is shown; only subgroups with at least ten effect sizes were analysed. Pooled effect estimates with 95% confidence intervals are reported numerically. Purple squares indicate significantly worse pain outcomes, green squares indicate significantly better pain outcomes, and grey squares indicate non-significant associations. Significant post hoc comparisons (two-sided *t* tests) are indicated. The corresponding orchard plot is provided in Supplementary Figure 28.

* = *p* < .05, ** = *p* < .01, *** = *p* < .001

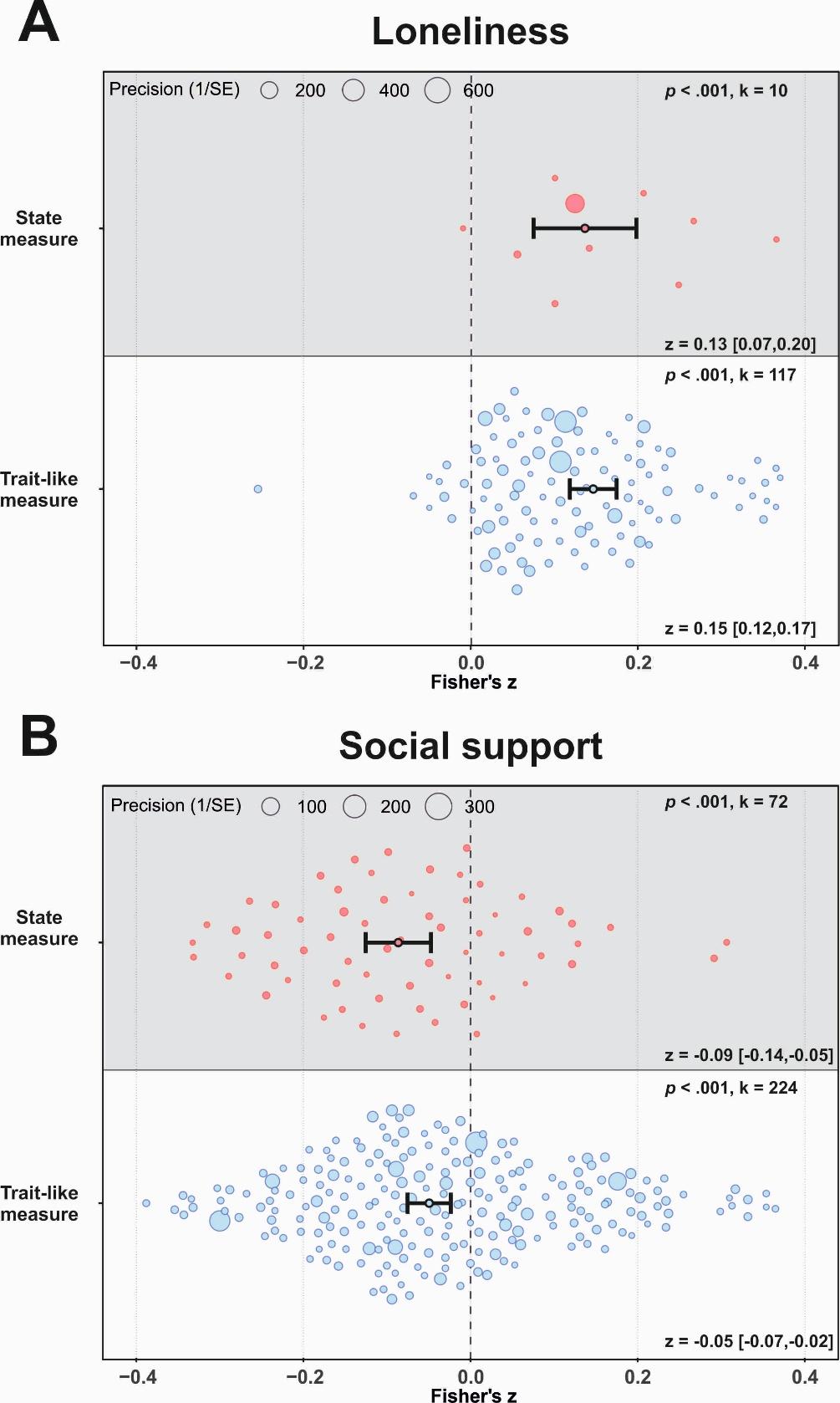

**Supplementary Figure 35.** Orchard plot illustrating associations between **(A)** loneliness and **(B)** social support and pain for state versus trait-like pain assessments. Each dot represents a cohort-level effect size, with dot size reflecting precision. The number of included effect sizes (k) is shown in the upper right of each panel. Pooled effects with 95% confidence intervals are displayed in the lower right and indicated by black dots and error bars.

**Supplementary Table 1**
Study quality assessment ratings for all included studies.

| **Study**  **ID** | **Study**  **name** | **Med. status** | **Co-**  **variates** | **Power analysis** | **Pre-**  **registered** | **Peer review** | **Randomi-**  **zation bias** | **Sequencing bias** | **Blinding bias** | **Attrition bias** |
| --- | --- | --- | --- | --- | --- | --- | --- | --- | --- | --- |
| 1 | Jaremka et al., 2014 | Yes | Yes | No | No | Yes | NA | NA | NA | NA |
| 2 | Loeffler et al., 2021 | No | Yes | No | No | Yes | NA | NA | NA | NA |
| 3 | Allen et al., 2020 | No | Yes | No | No | Yes | NA | NA | NA | NA |
| 4 | Jaremka et al., 2013 | No | Yes | No | No | Yes | NA | NA | NA | NA |
| 5 | Emerson et al., 2017 | No | Yes | No | No | Yes | NA | NA | NA | NA |
| 6 | Wolf et al., 2014 | No | Yes | No | No | Yes | NA | NA | NA | NA |
| 7 | Smith et al., 2019 | No | Yes | No | No | Yes | NA | NA | NA | NA |
| 8 | Yamada et al., 2021 | Yes | Yes | No | No | Yes | NA | NA | NA | NA |
| 9 | Nicolson et al., 2020 | No | Yes | No | No | Yes | NA | NA | NA | NA |
| 10 | Forgeron et al., 2024 | No | Yes | Yes | No | Yes | NA | NA | NA | NA |
| 11 | Boggero et al., 2019 | No | Yes | Yes | No | Yes | NA | NA | NA | NA |
| 12 | Powell et al., 2021 | No | Yes | No | No | Yes | NA | NA | NA | NA |
| 13 | Powell et al., 2022 | No | Yes | No | No | Yes | NA | NA | NA | NA |
| 14 | Zhang, 2024 | No | Yes | No | No | Yes | NA | NA | NA | NA |
| 15 | Wilson et al., 2022b | Yes | Yes | No | No | Yes | NA | NA | NA | NA |
| 16 | Stout et al., 2018 | No | Yes | No | No | Yes | NA | NA | NA | NA |
| 17 | Lutzman et al., 2020 | No | Yes | Yes | No | Yes | NA | NA | NA | NA |
| 18 | Camacho et al., 2024 | No | Yes | No | No | Yes | NA | NA | NA | NA |
| 19 | Noguchi et al., 2024 | No | Yes | No | No | Yes | NA | NA | NA | NA |
| 20 | Lee et al., 2023 | No | Yes | No | No | Yes | NA | NA | NA | NA |
| 21 | Stout et al., 2022 | No | Yes | No | No | Yes | NA | NA | NA | NA |
| 22 | Han et al., 2024 | No | Yes | No | No | No | NA | NA | NA | NA |
| 23 | Chan et al., 2014 | No | Yes | No | No | Yes | NA | NA | NA | NA |
| 24 | Gutiérrez et al., 2022 | No | Yes | No | No | Yes | NA | NA | NA | NA |
| 25 | Stout et al., 2021 | No | Yes | No | No | Yes | NA | NA | NA | NA |
| 26 | Fortuna et al., 2020 | No | Yes | No | No | Yes | NA | NA | NA | NA |
| 27 | Montejo-Carrasco et al., 2022 | Yes | Yes | No | No | Yes | NA | NA | NA | NA |
| 28 | Mikkelsen et al., 2020 | Yes | Yes | No | No | Yes | NA | NA | NA | NA |
| 29 | Mikkelsen et al., 2021 | Yes | Yes | No | No | Yes | NA | NA | NA | NA |
| 30 | Ahola et al., 2024 | No | Yes | No | No | Yes | NA | NA | NA | NA |
| 31 | Suzuki et al., 2024 | No | Yes | No | No | Yes | NA | NA | NA | NA |
| 32 | Mugoya et al., 2018 | No | Yes | No | No | Yes | NA | NA | NA | NA |
| 33 | Yu et al., 2021 | No | Yes | No | No | Yes | NA | NA | NA | NA |
| 34 | Adams et al., 2017 | No | Yes | No | No | Yes | NA | NA | NA | NA |
| 35 | Westergaard et al., 2021 | Yes | Yes | No | No | Yes | NA | NA | NA | NA |
| 36 | Emmungil et al., 2020 | No | Yes | No | No | Yes | NA | NA | NA | NA |
| 37 | Nguyen et al., 2023 | No | Yes | No | No | Yes | NA | NA | NA | NA |
| 38 | Doostdari et al., 2024 | No | Yes | No | No | Yes | NA | NA | NA | NA |
| 39 | Jacobs et al., 2006a | No | Yes | No | No | Yes | NA | NA | NA | NA |
| 40 | van Baarsen, 2008 | No | Yes | No | No | Yes | NA | NA | NA | NA |
| 41 | Akkaya & Kiyak, 2018 | No | Yes | Yes | No | Yes | NA | NA | NA | NA |
| 42 | Rokach et al., 2018 | No | Yes | No | No | Yes | NA | NA | NA | NA |
| 43 | Sahin et al., 2020 | Yes | No | No | No | Yes | NA | NA | NA | NA |
| 44 | Stensland et al., 2014 | No | Yes | No | No | Yes | NA | NA | NA | NA |
| 45 | Broen, 2022 | No | Yes | No | Yes | No | NA | NA | NA | NA |
| 46 | Stickley et al., 2016 | No | Yes | No | No | Yes | NA | NA | NA | NA |
| 47 | Waltz et al., 1998 | No | Yes | No | No | Yes | NA | NA | NA | NA |
| 48 | Christiansen et al., 2021 | No | Yes | No | No | Yes | NA | NA | NA | NA |
| 49 | Karayannis et al., 2018 | No | Yes | No | No | Yes | NA | NA | NA | NA |
| 50 | Bernstein et al., 2011 | No | Yes | No | No | Yes | No | NA | NA | No |
| 51 | Vervoort et al., 2011 | No | Yes | No | No | Yes | NA | Yes | NA | No |
| 52 | Kleck et al., 1976 | No | No | No | No | Yes | NA | No | NA | No |
| 53 | Krahé et al., 2015 | Yes | Yes | No | No | Yes | NA | No | NA | No |
| 54 | Master et al., 2009 | No | No | No | No | Yes | NA | No | NA | No |
| 55 | McClelland & McCubbin, 2008 | Yes | No | Yes | No | Yes | No | NA | NA | No |
| 56 | Vlaeyen et al., 2009 | No | Yes | Yes | No | Yes | No | NA | No | No |
| 57 | Modić Stanke & Ivanec, 2010 | Yes | No | No | No | Yes | No | No | NA | No |
| 58 | Montoya et al., 2004 | Yes | Yes | No | No | Yes | NA | Yes | NA | No |
| 59 | Roberts et al., 2015 | Yes | Yes | No | No | Yes | No | NA | NA | No |
| 60 | Sambo et al.., 2010 | No | Yes | No | No | Yes | NA | No | NA | No |
| 61 | Sullivan et al., 2004 | No | No | No | No | Yes | No | NA | NA | No |
| 62 | Brown et al., 2003 | Yes | Yes | Yes | No | Yes | No | No | NA | No |
| 63 | Edwards et al., 2017 | Yes | Yes | Yes | No | Yes | No | No | NA | No |
| 64 | Gallant & Hadjistavropoulos, 2017 | Yes | No | Yes | No | Yes | NA | No | No | No |
| 65 | Goldstein et al., 2016 | Yes | Yes | Yes | No | Yes | NA | No | NA | No |
| 66 | Jackson et al., 2005 | Yes | No | No | No | Yes | No | NA | No | No |
| 67 | Karmann et al., 2014 | Yes | Yes | No | No | Yes | No | No | NA | No |
| 68 | Borsook & MacDonald, 2010 | Yes | Yes | No | No | Yes | No | NA | NA | No |
| 69 | Baumgartner et al., 2023 | Yes | Yes | Yes | No | Yes | NA | NA | NA | No |
| 70 | DeWall et al., 2006 | No | Yes | No | No | Yes | No | NA | NA | No |
| 71 | Nanavaty et al, 2023 | Yes | No | No | No | Yes | No | NA | No | No |
| 72 | Hawthorne et al., 2013 | No | Yes | Yes | No | Yes | NA | NA | NA | NA |
| 73 | Oliviera et al., 2014 | No | Yes | No | No | Yes | NA | NA | NA | NA |
| 74 | Philpot et al., 2019 | Yes | Yes | No | No | Yes | NA | NA | NA | NA |
| 75 | Bach et al., 2019 | Yes | Yes | No | No | Yes | NA | No | NA | No |
| 76 | Hruschak et al., 2021 | Yes | Yes | Yes | No | Yes | NA | NA | NA | NA |
| 77 | Chen et al., 2014 | No | No | No | No | Yes | No | NA | NA | No |
| 78 | Richmond et al., 2018 | Yes | Yes | No | No | Yes | NA | NA | NA | NA |
| 79 | Yu et al., 2018 | No | Yes | No | No | Yes | No | NA | NA | No |
| 80 | Riva et al., 2011 | No | No | No | No | Yes | No | NA | NA | No |
| 81 | Baker et al., 2022 | No | Yes | No | No | Yes | NA | NA | NA | NA |
| 82 | Pieritz et al., 2017 | No | Yes | No | No | Yes | No | NA | NA | No |
| 83 | Osborne et al., 2007 | No | Yes | No | No | Yes | NA | NA | NA | NA |
| 84 | Boggero et al., 2024 | Yes | Yes | No | No | Yes | NA | NA | NA | NA |
| 85 | Musich et al., 2022 | No | Yes | No | No | Yes | NA | NA | NA | NA |
| 86 | Solé et al., 2020 | No | Yes | Yes | No | Yes | NA | NA | NA | NA |
| 87 | Bungert et al., 2015 | Yes | Yes | No | No | Yes | NA | No | NA | No |
| 88 | Stevens et al., 2020 | No | Yes | No | No | Yes | NA | NA | NA | NA |
| 89 | Martin et al., 2018 | No | No | Yes | No | Yes | NA | NA | NA | NA |
| 90 | Donaghy et al., 2022 | No | Yes | No | No | Yes | NA | NA | NA | NA |
| 91 | Antico et al., 2018 | No | No | Yes | No | Yes | NA | Yes | NA | No |
| 92 | Matthias et al., 2021 | No | Yes | No | No | Yes | NA | NA | NA | NA |
| 93 | Alphonsus et al., 2021 | No | Yes | No | No | Yes | NA | NA | NA | NA |
| 94 | Crockett & Turan, 2018 | No | Yes | No | No | Yes | NA | NA | NA | NA |
| 95 | Weiß et al., 2024a | Yes | Yes | No | No | Yes | NA | NA | NA | NA |
| 96 | Wand et al., 2022 | No | Yes | No | No | Yes | NA | NA | NA | NA |
| 97 | Hughes et al., 2014 | Yes | Yes | No | No | Yes | NA | NA | NA | NA |
| 98 | Fisher et al., 2021 | Yes | Yes | No | No | Yes | NA | NA | NA | NA |
| 99 | Holtzman et al., 2004 | No | Yes | No | No | Yes | NA | NA | NA | NA |
| 100 | Wolf et al., 2015 | No | Yes | No | No | Yes | NA | NA | NA | NA |
| 101 | Croda, 2015 | No | Yes | No | No | No | NA | NA | NA | NA |
| 102 | Reddan et al., 2020 | No | No | No | No | Yes | NA | No | NA | No |
| 103 | Guillory et al., 2015 | Yes | Yes | No | No | Yes | No | NA | Yes | No |
| 104 | Motl et al., 2008 | No | Yes | No | No | Yes | NA | NA | NA | NA |
| 105 | Jump et al., 2005 | Yes | Yes | No | No | Yes | NA | NA | NA | NA |
| 106 | Keefe et al., 2003 | No | No | No | No | Yes | NA | NA | NA | NA |
| 107 | Lee et al., 2015 | No | Yes | No | No | Yes | NA | NA | NA | NA |
| 108 | Li et al., 2017 | Yes | Yes | No | No | Yes | NA | NA | NA | NA |
| 109 | Bjelkarøy et al., 2021 | Yes | Yes | No | No | Yes | NA | NA | NA | NA |
| 110 | Penn et al., 2019 | Yes | Yes | No | No | Yes | NA | NA | NA | NA |
| 111 | Ferreira-Valente et al., 2014 | No | Yes | No | No | Yes | NA | NA | NA | NA |
| 112 | Cohen et al., 2007 | No | No | No | No | Yes | NA | NA | NA | NA |
| 113 | Forgeron et al., 2018 | No | Yes | Yes | No | Yes | NA | NA | NA | NA |
| 114 | Evans et al., 2021 | No | Yes | No | No | Yes | NA | NA | NA | NA |
| 115 | Gulewitsch et al, 2018 | No | No | No | No | Yes | NA | Yes | NA | No |
| 116 | Kreuder et al., 2018 | No | No | No | No | Yes | No | No | No | No |
| 117 | Yamada et al., 2020 | No | Yes | No | No | Yes | NA | NA | NA | NA |
| 118 | Elfering et al., 2002 | No | No | No | No | Yes | NA | NA | NA | NA |
| 119 | LeRoy, 2015 | No | Yes | Yes | No | No | No | NA | NA | No |
| 120 | Oraison & Kennedy, 2019 | No | Yes | No | No | Yes | NA | NA | NA | NA |
| 121 | Driscoll et al., 2015 | Yes | Yes | No | No | Yes | NA | NA | NA | NA |
| 122 | Cho et al, 2011 | Yes | Yes | No | No | Yes | NA | NA | NA | NA |
| 123 | Buenaver et al., 2007 | No | Yes | No | No | Yes | NA | NA | NA | NA |
| 124 | Musich et al., 2019 | Yes | Yes | No | No | Yes | NA | NA | NA | NA |
| 125 | Duschek et al., 2019 | No | No | No | No | Yes | NA | No | NA | No |
| 126 | Khazaeipour et al., 2017 | No | No | No | No | Yes | NA | NA | NA | NA |
| 127 | Samulowitz et al, 2022 | No | Yes | No | No | Yes | NA | NA | NA | NA |
| 128 | Wilson et al., 2022 | No | Yes | No | No | Yes | NA | NA | NA | NA |
| 129 | Neumann et al., 2023 | Yes | No | Yes | Yes | Yes | No | No | NA | No |
| 130 | Rzeszutek et al., 2015 | No | Yes | No | No | Yes | NA | NA | NA | NA |
| 131 | Larice et al., 2020 | No | Yes | No | No | Yes | NA | NA | NA | NA |
| 132 | Matos et al., 2017 | No | Yes | No | No | Yes | NA | NA | NA | NA |
| 133 | Stroud et al., 2006 | No | Yes | No | No | Yes | NA | NA | NA | NA |
| 134 | Gunduz et al., 2018 | No | No | No | No | Yes | NA | NA | NA | NA |
| 135 | Afrashteh & Abbasi, 2023 | Yes | Yes | No | No | Yes | NA | NA | NA | NA |
| 136 | Leung et al., 2015 | Yes | Yes | No | No | Yes | NA | NA | NA | NA |
| 137 | Ghadimi et al., 2023 | Yes | Yes | No | No | Yes | NA | NA | NA | NA |
| 138 | Schmitter et al., 2009 | No | No | Yes | No | Yes | NA | NA | NA | NA |
| 139 | Wang et al., 2018 | No | Yes | No | No | Yes | NA | NA | NA | NA |
| 140 | Gebhardt et al., 2021 | No | Yes | No | No | Yes | NA | NA | NA | NA |
| 141 | Shim et al., 2017 | No | Yes | No | No | Yes | NA | NA | NA | NA |
| 142 | Tse et al., 2013 | Yes | No | No | No | Yes | NA | NA | NA | NA |
| 143 | Baur et al., 2016 | No | Yes | No | No | Yes | NA | NA | NA | NA |
| 144 | Miró et al., 2017 | No | Yes | No | No | Yes | NA | NA | NA | NA |
| 145 | Ren et al., 2021 | No | Yes | No | No | Yes | NA | NA | NA | NA |
| 146 | Hanssen et al., 2014 | Yes | Yes | Yes | No | Yes | NA | NA | NA | NA |
| 147 | von Mohr et al., 2018 | No | Yes | No | No | Yes | NA | No | NA | No |
| 148 | Sturgeon et al., 2016 | No | Yes | No | No | Yes | NA | NA | NA | NA |
| 149 | Alonso et al., 2001 | No | No | No | No | Yes | NA | NA | NA | NA |
| 150 | Hirsh et al., 2010 | Yes | Yes | No | No | Yes | NA | NA | NA | NA |
| 151 | Jablonska et al., 2006 | Yes | Yes | No | No | Yes | NA | NA | NA | NA |
| 152 | Zeng et al., 2015 | No | Yes | No | No | Yes | NA | NA | NA | NA |
| 153 | Gaffey et al., 2018 | No | Yes | No | No | Yes | NA | NA | NA | NA |
| 154 | Bean et al., 2022 | Yes | Yes | Yes | No | Yes | NA | NA | NA | NA |
| 155 | Riem et al., 2021 | Yes | Yes | Yes | No | Yes | No | NA | No | No |
| 156 | Yang et al., 2022 | No | Yes | No | No | Yes | NA | NA | NA | NA |
| 157 | Åkerblom et al., 2015 | Yes | Yes | Yes | No | Yes | NA | NA | NA | NA |
| 158 | Hagglund et al., 1995 | No | Yes | No | No | Yes | NA | NA | NA | NA |
| 159 | Evans et al., 2005 | No | Yes | No | No | Yes | NA | NA | NA | NA |
| 160 | Mitchell et al., 2016 | Yes | Yes | No | No | Yes | NA | NA | NA | NA |
| 161 | Chen et al., 2020 | No | Yes | No | No | Yes | NA | NA | NA | NA |
| 162 | Mallon et al., 2021 | Yes | Yes | No | No | Yes | NA | NA | NA | NA |
| 163 | Larbig et al., 2019 | Yes | Yes | No | No | Yes | NA | NA | NA | NA |
| 164 | Fallon et al., 2021 | Yes | Yes | No | No | Yes | NA | NA | NA | NA |
| 165 | Hung et al., 2016 | Yes | Yes | No | No | Yes | NA | NA | NA | NA |
| 166 | Kindt et al., 2018 | No | Yes | No | No | Yes | NA | NA | NA | NA |
| 167 | Serbic et al., 2020 | No | Yes | No | No | Yes | NA | NA | NA | NA |
| 168 | Moye et al., 2014 | No | Yes | No | No | Yes | NA | NA | NA | NA |
| 169 | Beugnot, 2001 | No | Yes | No | No | No | NA | NA | NA | NA |
| 170 | Nieto et al., 2020 | Yes | No | No | No | Yes | NA | NA | NA | NA |
| 171 | Wiesmann et al., 2014 | No | Yes | No | No | Yes | NA | NA | NA | NA |
| 172 | Tse et al., 2011 | Yes | No | No | No | Yes | NA | NA | NA | NA |
| 173 | Expósito-Vizcaíno et al., 2019 | No | Yes | No | No | Yes | NA | NA | NA | NA |
| 174 | Taal et al., 1993 | Yes | No | No | No | Yes | NA | NA | NA | NA |
| 175 | Almeida et al., 2019 | No | No | No | No | Yes | NA | NA | NA | NA |
| 176 | Tsai et al., 2003 | No | Yes | Yes | No | Yes | NA | NA | NA | NA |
| 177 | Perez et al., 2008 | Yes | Yes | No | No | Yes | NA | NA | NA | NA |
| 178 | Law et al., 2015 | No | Yes | No | No | Yes | NA | NA | NA | NA |
| 179 | Leung et al., 2014 | No | Yes | No | No | Yes | NA | NA | NA | NA |
| 180 | László et al., 2008 | No | Yes | No | No | Yes | NA | NA | NA | NA |
| 181 | Yamada et al., 2016 | No | Yes | No | No | Yes | NA | NA | NA | NA |
| 182 | Blozik et al., 2009 | No | Yes | No | No | Yes | NA | NA | NA | NA |
| 183 | Osborne et al, 2006 | No | Yes | No | No | Yes | NA | NA | NA | NA |
| 184 | Roberts et al., 2008 | No | No | No | No | Yes | NA | NA | NA | NA |
| 185 | Merz et al., 2016 | Yes | Yes | No | No | Yes | NA | NA | NA | NA |
| 186 | Johnsen et al., 2023 | No | Yes | Yes | No | Yes | NA | NA | NA | NA |
| 187 | Hermann et al., 2008 | No | Yes | No | No | Yes | NA | NA | NA | NA |
| 188 | Rogers et al., 2015 | Yes | Yes | No | No | Yes | NA | NA | NA | NA |
| 189 | Wilson & Simpson, 2016 | Yes | Yes | No | No | Yes | NA | NA | NA | NA |
| 190 | Jensen et al., 2002 | No | Yes | No | No | Yes | NA | NA | NA | NA |
| 191 | Evers et al., 2003 | Yes | Yes | No | No | Yes | NA | NA | NA | NA |
| 192 | Stefaniak et al., 2012 | Yes | No | No | No | Yes | NA | NA | NA | NA |
| 193 | Cici et al., 2023 | Yes | Yes | No | No | Yes | NA | NA | NA | NA |
| 194 | Jahre et al., 2021 | No | Yes | No | No | Yes | NA | NA | NA | NA |
| 195 | Boggero et al., 2015 | No | Yes | No | No | Yes | NA | NA | NA | NA |
| 196 | Ruben et al., 2016 | No | No | Yes | No | Yes | No | NA | No | No |
| 197 | Grov et al., 2015 | Yes | Yes | No | No | Yes | NA | NA | NA | NA |
| 198 | Gerdle et al., 2023 | Yes | Yes | No | No | Yes | NA | NA | NA | NA |
| 199 | Tripp et al., 2006 | No | Yes | No | No | Yes | NA | NA | NA | NA |
| 200 | Stén et al., 2014 | Yes | Yes | No | No | Yes | NA | NA | NA | NA |
| 201 | Fillingim et al., 2003 | Yes | No | No | No | Yes | NA | NA | NA | NA |
| 202 | Blanco-Hungría et al., 2012 | Yes | Yes | No | No | Yes | NA | NA | NA | NA |
| 203 | Munk et al., 2023 | Yes | Yes | No | No | Yes | NA | NA | NA | NA |
| 204 | Berlit et al., 2018 | No | No | No | No | Yes | NA | NA | NA | NA |
| 205 | Dorner et al., 2018 | No | Yes | No | No | Yes | NA | NA | NA | NA |
| 206 | Hurwitz et al., 2009 | No | Yes | No | No | Yes | NA | NA | NA | NA |
| 207 | López-Martínez et al., 2008 | Yes | Yes | No | No | Yes | NA | NA | NA | NA |
| 208 | Kerns et al., 2002 | Yes | Yes | No | No | Yes | NA | NA | NA | NA |
| 209 | Flor et al., 1995 | No | No | No | No | Yes | NA | NA | NA | NA |
| 210 | Koopman et al., 1998 | No | Yes | No | No | Yes | NA | NA | NA | NA |
| 211 | Kelsen et al., 1995 | Yes | No | No | No | Yes | NA | NA | NA | NA |
| 212 | Willey and Silliman, 1990 | No | Yes | No | No | Yes | NA | NA | NA | NA |
| 213 | Isacsson et al., 1995 | No | Yes | No | No | Yes | NA | NA | NA | NA |
| 214 | Takeyachi et al., 2003 | No | Yes | No | No | Yes | NA | NA | NA | NA |
| 215 | Schneider et al., 2005 | No | Yes | No | No | Yes | NA | NA | NA | NA |
| 216 | Siviero et al., 2020 | Yes | Yes | No | No | Yes | NA | NA | NA | NA |
| 217 | Mandl et al., 2024 | No | Yes | No | No | Yes | NA | NA | NA | NA |
| 218 | Shi et al., 2024 | No | Yes | No | No | Yes | NA | NA | NA | NA |
| 219 | Lynch-Jordan et al., 2015 | No | Yes | Yes | No | Yes | NA | NA | NA | NA |
| 220 | Leonti et al., 2025 | No | Yes | Yes | No | Yes | NA | NA | NA | NA |
| 221 | Falck et al., 2025 | No | No | No | No | Yes | NA | NA | NA | NA |
| 222 | Bloomberg et al., 2025 | No | Yes | No | No | Yes | NA | NA | NA | NA |
| 223 | Ponce et al., 2025 | No | Yes | No | No | Yes | NA | NA | NA | NA |
| 224 | Rendón et al., 2025 | Yes | Yes | Yes | No | Yes | NA | NA | NA | NA |
| 225 | Forgeron et al., 2025 | No | Yes | Yes | No | Yes | NA | NA | NA | NA |
| 226 | Vestergaard et al., 2025 | No | Yes | No | No | Yes | NA | NA | NA | NA |
| 227 | HaGani et al., 2025 | No | Yes | No | No | Yes | NA | NA | NA | NA |
| 228 | Robinson-Whelen et al., 2025 | No | Yes | Yes | No | Yes | NA | NA | NA | NA |
| 229 | Ikedo et al., 2025 | No | Yes | No | No | No | NA | NA | NA | NA |
| 230 | Harsted et al., 2025 | No | Yes | No | No | Yes | NA | NA | NA | NA |
| 231 | Eshkevar-Faraji et al., 2025 | No | Yes | Yes | No | Yes | NA | NA | NA | NA |
| 232 | Scorsone et al., 2025 | Yes | No | No | No | Yes | NA | NA | NA | NA |
| 233 | McCurry et al., 2025 | No | Yes | No | No | Yes | NA | NA | NA | NA |
| 234 | Johnstone et al., 2025 | Yes | Yes | Yes | No | Yes | NA | NA | NA | NA |
| 235 | Codden et al., 2025 | Yes | Yes | No | No | Yes | NA | NA | NA | NA |
| 236 | Lieber et al., 2025 | Yes | Yes | No | No | Yes | NA | NA | NA | NA |
| 237 | Macchia & Fett, 2025 | No | Yes | No | No | Yes | NA | NA | NA | NA |
| 238 | Che et al., 2018 | Yes | No | No | No | Yes | No | No | NA | No |
| 239 | Ekhlom et al., 2025 | No | Yes | No | No | Yes | NA | NA | NA | NA |

**Supplementary Table 2**
Studies excluded from the meta-analysis with reasons for exclusion.

| **Study number** | **Study name** | **Reason for exclusion** |
| --- | --- | --- |
| 1 | Abrahamsson et al., 2025 | No measure of social connectedness |
| 2 | Amiel et al., 2016 | No measure of social connectedness |
| 3 | Ang et al., 2018 | No measure of social connectedness |
| 4 | Atoyebi & Wister, 2017 | No reported effect |
| 5 | Backe et al., 2018 | No reported effect |
| 6 | Bahl et al., 2025 | No measure of social connectedness |
| 7 | Bannon et al., 2021 | No reported effect |
| 8 | Barad et al., 2021 | No reported effect |
| 9 | Batley et al., 2018 | No reported effect |
| 10 | Baumgartner et al., 2023 | Effect size already used |
| 11 | Bdair et al., 2025 | No measure of social connectedness |
| 12 | Bernardes et al., 2024 | No measure of social connectedness |
| 13 | Birnie et al., 2016 | No reported effect |
| 14 | Block et al., 2018 | No reported effect |
| 15 | Boggero et al., 2016 | Effect size already used |
| 16 | Boggero et al., 2016 | Effect size already used |
| 17 | Boggero et al., 2016 | Effect size already used |
| 18 | Brandstetter et al., 2017 | No reported effect |
| 19 | Broekman et al., 2025 | No reported effect |
| 20 | Brown et al., 2018 | No measure of social connectedness |
| 21 | Brown et al., 2020 | No reported effect |
| 22 | Burns et al., 2013 | No measure of social connectedness |
| 23 | Casey et al., 2025 | No reported effect |
| 24 | Castarlenas et al, 2023 | No reported effect |
| 25 | Castarlenas et al, 2025 | No reported effect |
| 26 | Chaidez et al., 2014 | No reported effect |
| 27 | Chambers et al., 2002 | No control condition |
| 28 | Che et al, 2019 | No measure of social connectedness |
| 29 | Cheatle et al., 2014 | No reported effect |
| 30 | Chen et al., 2025 | No reported effect |
| 31 | Cogan & Spinato, 1987 | No measure of pain |
| 32 | Correa et al., 2025 | No measure of pain |
| 33 | Crooks et al., 2025 | No reported effect |
| 34 | Davis et al., 2025 | No measure of pain |
| 35 | De Ruddere et al., 2016 | No reported effect |
| 36 | de Sola et al., 2016 | No reported effect |
| 37 | De Vellis et al., 1986 | No reported effect |
| 38 | De Vellis et al., 1986 | No reported effect |
| 39 | Dehghan et al., 2021 | No reported effect |
| 40 | Demmelmaier et al., 2008 | No control condition |
| 41 | Deng et al., 2025 | No measure of social connectedness |
| 42 | Dickinson, 2009 | No measure of social connectedness |
| 43 | Dijkstra et al., 2007 | No measure of social connectedness |
| 44 | DiLorenzo et al., 2017 | No reported effect |
| 45 | Docking et al., 2014 | No reported effect |
| 46 | Dysvik et al., 2004 | No measure of pain |
| 47 | Eccleston et al., 2004 | No measure of social connectedness |
| 48 | Edusei et al., 2017 | No access |
| 49 | Eisenberger et al., 2011 | No measure of social connectedness |
| 50 | Eisenberger et al., 2011 | No measure of social connectedness |
| 51 | Emerson et al., 2017 | Effect size already used |
| 52 | Fales & Noel, 2020 | No reported effect |
| 53 | Fan et al., 2021 | No measure of pain |
| 54 | Faucett & Levine, 1991 | No reported effect |
| 55 | Fernández-Peña et al., 2018 | No reported effect |
| 56 | Fernández-Peña et al., 2020 | No reported effect |
| 57 | Fishman et al., 1995 | No control condition |
| 58 | Flowers et al, 2021 | No reported effect |
| 59 | Flowers et al., 2023 | No reported effect |
| 60 | Floyd, 2016 | No measure of pain |
| 61 | Freedman & Sternberg, 2009 | No reported effect |
| 62 | Freij et al., 2025 | No access |
| 63 | Galloway et al., 2019 | No reported effect |
| 64 | Gen et al., 2025 | No control condition |
| 65 | Gil et al., 1987 | No measure of pain |
| 66 | Gil et al., 1990 | No reported effect |
| 67 | Gyasi et al., 2025 | No reported effect |
| 68 | Hanley et al., 2008 | No reported effect |
| 69 | Hassinger et al., 1999 | No measure of social connectedness |
| 70 | Hayes & Wolf, 1984 | No reported effect |
| 71 | Hechler et al., 2010 | No measure of social connectedness |
| 72 | Helmhout et al., 2010 | No measure of pain |
| 73 | Helmhout et al., 2010 | No measure of pain |
| 74 | Henoch et al., 2007 | No reported effect |
| 75 | Ho et al., 2016 | No measure of social connectedness |
| 76 | Hsu et al., 2025 | No measure of social connectedness |
| 77 | Hughes et al., 2001 | No reported effect |
| 78 | Iliffe et al., 2009 | No reported effect |
| 79 | Jacobs et al., 2006b | No reported effect |
| 80 | Jahn et al., 2025 | No control condition |
| 81 | Johnson & Dunbar, 2016 | No measure of pain |
| 82 | Johnson et al., 2025 | No reported effect |
| 83 | Jollife & Nicholas, 2004 | No measure of social connectedness |
| 84 | Kaminsky et al., 2006 | No reported effect |
| 85 | Keefe et al., 1999 | No measure of social connectedness |
| 86 | Kent et al., 2014 | No reported effect |
| 87 | Kim et al., 2015 | No reported effect |
| 88 | Kim et al., 2022 | No reported effect |
| 89 | Kishimoto et al., 2016 | No reported effect |
| 90 | Kittel et al., 20225 | No reported effect |
| 91 | Klapow et al., 1995 | No control condition |
| 92 | Kleck et al., 1976 | Effect size already used |
| 93 | Koebner et al., 2018 | No reported effect |
| 94 | Kong et al., 2023 | No access |
| 95 | Korff & Simon, 1996 | No reported effect |
| 96 | Krahé et al., 2015 | Effect size already used |
| 97 | Kühner et al., 2025 | No reported effect |
| 98 | Kulik & Mahler, 1989 | No measure of pain |
| 99 | Lam & Vuolo, 2023 | No measure of pain |
| 100 | Latifi et al., 2019 | No accessible language |
| 101 | Lauver & Johnson, 1997 | No measure of pain |
| 102 | Lengerud et al., 2018 | No reported effect |
| 103 | Leung et al., 2015 | No measure of social connectedness |
| 104 | Liang et al., 2025 | No reported effect |
| 105 | Madani et al., 2025 | No reported effect |
| 106 | Maité et al., 2025 | No reported effect |
| 107 | Martin et al., 2015 | No reported effect |
| 108 | Marttinen et al., 2019 | No measure of social connectedness |
| 109 | Matos & Bernardes, 2013 | No measure of social connectedness |
| 110 | Matos et al., 2015 | No reported effect |
| 111 | Matos et al., 2016 | Effect size already used |
| 112 | Matos et al., 2017 | No measure of social connectedness |
| 113 | Matsuda et al. 2025 | No reported effect |
| 114 | Matsuda et al., 2025 | No measure of social connectedness |
| 115 | Mayo et al., 2025 | No reported effect |
| 116 | Mazza et al., 2023 | No control condition |
| 117 | McKillop et al., 2016 | No reported effect |
| 118 | McWilliams et al., 2014 | No measure of social connectedness |
| 119 | Miaskowski et al., 2012 | No measure of social connectedness |
| 120 | Miki et al., 2024 | No reported effect |
| 121 | Mikkelsen et al., 2022 | No reported effect |
| 122 | Miller, 1985 | No reported effect |
| 123 | Mitchinson et al., 2008 | Unreliable data |
| 124 | Montejo et al., 2020 | No reported effect |
| 125 | Montejo-Carrasco et al., 2020 | No reported effect |
| 126 | Nakae et al., 2025 | No measure of social connectedness |
| 127 | Nguyen et al., 2012 | No measure of pain |
| 128 | Nguyen et al., 2025 | No access |
| 129 | Nixon et al., 2025 | No reported effect |
| 130 | Nixon, 1994 | No reported effect |
| 131 | Novembre et al., 2014 | No reported effect |
| 132 | Nye, 2022 | Review article |
| 133 | Ogliari et al., 2023 | No reported effect |
| 134 | Ojala et al., 2013 | No measure of pain |
| 135 | Olawa et al., 2025 | No reported effect |
| 136 | Ong et al., 2021 | No reported effect |
| 137 | Orlando et al., 2025 | No reported effect |
| 138 | Pang et al., 2009 | No reported effect |
| 139 | Park et al., 2016 | No reported effect |
| 140 | Park et al., 2025 | No reported effect |
| 141 | Passchier et al., 1996 | No control condition |
| 142 | Peeters & Vlaeyen, 2011 | No measure of social connectedness |
| 143 | Pivodic et al., 2024 | No reported effect |
| 144 | Platow et al., 2006 | No reported effect |
| 145 | Polański et al., 2022 | No reported effect |
| 146 | Poon et al., 2020 | No measure of social connectedness |
| 147 | Ptacek et al., 1995 | No measure of social connectedness |
| 148 | Qualter et al., 2020 | No reported effect |
| 149 | Rao et al., 2022 | No measure of pain |
| 150 | Reinke et al., 2025 | No reported effect |
| 151 | Riva et al., 2011 | No measure of social connectedness |
| 152 | Riva et al., 2014 | No reported effect |
| 153 | Roberts et al., 1996 | No reported effect |
| 154 | Roberts-West et al., 2023 | No reported effect |
| 155 | Rohde et al., 2024 | No reported effect |
| 156 | Rokach et al., 2016 | Effect size already used |
| 157 | Rokach et al., 2017 | No measure of pain |
| 158 | Romano et al., 2000 | No measure of pain |
| 159 | Rosenberg et al., 2025 | No access |
| 160 | Rummans et al., 1998 | No reported effect |
| 161 | Rzeszutek et al., 2015 | No reported effect |
| 162 | Salafia et all., 2025 | No reported effect |
| 163 | Şan et al., 2021 | No measure of social connectedness |
| 164 | Saravanan et al., 2021 | No access |
| 165 | Sato et al., 2025 | No reported effect |
| 166 | Schulz-Kindermann et al., 2002 | No reported effect |
| 167 | Schwab et al., 2021 | No reported effect |
| 168 | Scott et al., 2022 | No reported effect |
| 169 | Scott et al., 2022 | Effect size already used |
| 170 | Shade et al., 2025 | No measure of social connectedness |
| 171 | Şimşek et al., 2025 | No measure of social connectedness |
| 172 | Smedley et al., 2015 | No reported effect |
| 173 | Smite et al., 2012 | No measure of social connectedness |
| 174 | Smith et al., 2025 | No measure of social connectedness |
| 175 | Spiegel et al., 1994 | No reported effect |
| 176 | Staes et al., 2007 | No reported effect |
| 177 | Strang and Qvarner, 1990 | No access |
| 178 | Strang, 1992 | No reported effect |
| 179 | Tagliaferri et al., 2022 | No reported effect |
| 180 | Tan et al., 2022 | No reported effect |
| 181 | Tanaka et al. 2017 | No measure of pain |
| 182 | Tandon et al., 2024 | No reported effect |
| 183 | Telli & Akkus, 2025 | No access |
| 184 | Thoresen et al., 2018 | No measure of pain |
| 185 | Tomás et al., 2025 | No measure of pain |
| 186 | Tree, 2009 | No reported effect |
| 187 | Tse et al., 2010 | No reported effect |
| 188 | Tse et al., 2012 | No reported effect |
| 189 | Tse et al., 2014 | No control condition |
| 190 | Turner et al., 1987 | No reported effect |
| 191 | Umeda & Kim, 2025 | No measure of social connectedness |
| 192 | Van Der Lugt et al., 2011 | No reported effect |
| 193 | Vervoort et al., 2008 | No control condition |
| 194 | Vervoort et al., 2011 | Effect size already used |
| 195 | Vigil et al., 2013 | No reported effect |
| 196 | Vigil et al., 2014 | No measure of social connectedness |
| 197 | Villumsen et al., 2016 | No reported effect |
| 198 | Vlaeyen et al., 2009 | Effect size already used |
| 199 | von Mohr et al., 2017 | No control condition |
| 200 | Vosvick et al., 2004 | No reported effect |
| 201 | Wang et al., 2020 | No measure of social connectedness |
| 202 | Wei et al., 2025 | No measure of pain |
| 203 | Weinstein et al., 2016 | No measure of social connectedness |
| 204 | Weiß et al., 2024b | Review article |
| 205 | Wernicke et al., 2015 | No measure of pain |
| 206 | Widerström-Noga et al., 2007 | No reported effect |
| 207 | Williams & Cano, 2014 | No measure of social connectedness |
| 208 | Wilson & Ruben, 2011 | No measure of social connectedness |
| 209 | Wilson et al., 2022b | No reported effect |
| 210 | Wong et al., 2024 | No reported effect |
| 211 | Yao et al., 2020 | No reported effect |
| 212 | Yates et al., 2023 | No reported effect |
| 213 | Yim et al., 2016 | No reported effect |
| 214 | Younger et al., 2010 | No measure of social connectedness |
| 215 | Yu et al., 2022 | No measure of pain |
| 216 | Zhou et al., 2025 | No measure of pain |
| 217 | Zulfiqar et al., 2025 | No measure of pain |
| 218 | Zwolinski, 2023a | No access |
| 219 | Zwolinski, 2023b | No reported effect |
| 220 | Zwolinski, 2025 | No access |

**Supplementary Table 3**

Bayes Factor analysis for overall effect.

| Model | Moderator level | σ = 0.05 | σ = 0.10 | σ = 0.15 |
| --- | --- | --- | --- | --- |
|  |  | BF10 | BF10 | BF10 |
| Overall | N/A | > 100 | > 100 | > 100 |

**Supplementary Table 4**

Bayes Factor analysis by social connectedness dimension.

| Model | Moderator level | σ = 0.05 | σ = 0.10 | σ = 0.15 |
| --- | --- | --- | --- | --- |
|  |  | BF10 | BF10 | BF10 |
| Social Connected-  ness | Loneliness | > 100 | > 100 | > 100 |
|  | Social Exclusion | 0.93 | 0.70 | 0.54 |
|  | Social Isolation | >100 | > 100 | > 100 |
|  | Social Support | > 100 | > 100 | 72.70 |

**Supplementary Table 5**

Bayes Factor analysis for social connectedness dimension by pain type interaction.

| Model | Moderator level | σ = 0.05 | σ = 0.10 | σ = 0.15 |
| --- | --- | --- | --- | --- |
|  |  | BF10 | BF10 | BF10 |
| Social Connected-  ness  *Pain Types | Loneliness Frequency | 22.00 | > 100 | > 100 |
|  | Loneliness Intensity | > 100 | > 100 | > 100 |
|  | Loneliness Interference | > 100 | > 100 | > 100 |
|  | Social Exclusion Threshold | 0.94 | 0.87 | 0.72 |
|  | Social Exclusion Tolerance | 0.96 | 0.81 | 0.72 |
|  | Social Isolation Intensity | 2.82 | 6.24 | 6.49 |
|  | Social Isolation Interference | 10.62 | 44.45 | > 100 |
|  | Social Support Catastrophizing | 0.91 | 0.70 | 0.49 |
|  | Social Support Disability | 0.74 | 0.75 | 0.56 |
|  | Social Support Duration | 1.61 | 1.96 | 1.71 |
|  | Social Support Intensity | 2.99 | 4.96 | 4.96 |
|  | Social Support Interference | 0.53 | 0.29 | 0.19 |
|  | Social Support Unpleasantness | 6.35 | 10.35 | 12.24 |

**Supplementary Table 6**

Bayes Factor analysis for perceived vs. present social support.

| Model | Moderator level | σ = 0.05 | σ = 0.10 | σ = 0.15 |
| --- | --- | --- | --- | --- |
|  |  | BF10 | BF10 | BF10 |
| Social Support  Perceived vs Present | Perceived Social Support | 1.81 | 1.13 | 1.12 |
|  | Present Social Support | > 100 | > 100 | > 100 |

**Supplementary Table 7**

Bayes Factor analysis for social support types.

| Model | Moderator level | σ = 0.05 | σ = 0.10 | σ = 0.15 |
| --- | --- | --- | --- | --- |
|  |  | BF10 | BF10 | BF10 |
| Social Support  Types | Emotional Support | 0.62 | 0.36 | 0.26 |
|  | Instrumental Support | 0.72 | 0.44 | 0.32 |
|  | Familiar Social Presence | 7.03 | 14.76 | 10.99 |
|  | Unfamiliar Social Presence | 10.82 | 25.63 | 16.20 |

**Supplementary Table 8**

Bayes Factor analysis for social connectedness by clinical sample interaction.

| Model | Moderator level | σ = 0.05 | σ = 0.10 | σ = 0.15 |
| --- | --- | --- | --- | --- |
|  |  | BF10 | BF10 | BF10 |
| Social Connectedness  *Clinical Sample | Loneliness Healthy | > 100 | > 100 | > 100 |
|  | Loneliness Clinical | > 100 | > 100 | > 100 |
|  | Social Exclusion Healthy | 0.88 | 0.66 | 0.52 |
|  | Social Isolation Clinical | 92.97 | > 100 | > 100 |
|  | Social Support Healthy | 9.57 | 9.51 | 5.13 |
|  | Social Support Clinical | 16.40 | 7.8 | 8.51 |

**Supplementary Table 9**

Bayes Factor analysis for social connectedness by pain classification interaction.

| Model | Moderator level | σ = 0.05 | σ = 0.10 | σ = 0.15 |
| --- | --- | --- | --- | --- |
|  |  | BF10 | BF10 | BF10 |
| Social Connectedness  *Pain Classification | Loneliness Secondary Pain | 45.68 | > 100 | > 100 |
|  | Social Isolation Secondary Pain | 5.29 | 6.00 | 6.84 |
|  | Social Support Primary Pain | 1.58 | 1.46 | 1.30 |
|  | Social Support Secondary Pain | > 100 | 43.88 | > 100 |

**Supplementary Table 10**

Bayes Factor analysis for social connectedness by association type interaction.

| Model | Moderator level | σ = 0.05 | σ = 0.10 | σ = 0.15 |
| --- | --- | --- | --- | --- |
|  |  | BF10 | BF10 | BF10 |
| Social Connectedness  *Association Type | Loneliness Cross-Sectional | > 100 | > 100 | > 100 |
|  | Loneliness Longitudinal | 2.53 | 3.48 | 3.08 |
|  | Social Isolation Cross-Sectional | 68.83 | > 100 | 50.88 |
|  | Social Support Cross-Sectional | 19.48 | 10.44 | 10.78 |
|  | Social Support Longitudinal | 1.93 | 2.29 | 1.93 |

**Supplementary Table 11**

Bayes Factor analysis for social connectedness by study design interaction.

| Model | Moderator level | σ = 0.05 | σ = 0.10 | σ = 0.15 |
| --- | --- | --- | --- | --- |
|  |  | BF10 | BF10 | BF10 |
| Social Connectedness  *Study Design | Social Exclusion Between-Person | 1.02 | 0.94 | 0.79 |
|  | Social Support Between-Person | 0.88 | 0.67 | 0.51 |
|  | Social Support Within-Person | 15.53 | 46.67 | 16.54 |

**Supplementary Table 12**

Bayes Factor analysis for social connectedness by pain measurement interaction.

| Model | Moderator level | σ = 0.05 | σ = 0.10 | σ = 0.15 |
| --- | --- | --- | --- | --- |
|  |  | BF10 | BF10 | BF10 |
| Social Connectedness  *Pain Measurement | Loneliness Questionnaire | > 100 | > 100 | > 100 |
|  | Loneliness Single Item | > 100 | > 100 | > 100 |
|  | Social Exclusion Single Item | 0.93 | 0.82 | 0.64 |
|  | Social Isolation Questionnaire | 4.06 | 12.27 | 9.58 |
|  | Social Isolation Single Item | 9.14 | 9.98 | 8.21 |
|  | Social Support Questionnaire | 0.46 | 0.24 | 0.17 |
|  | Social Support Single Item | > 100 | > 100 | > 100 |

**Supplementary Table 13**

Bayes Factor analysis for social connectedness by social measurement interaction.

| Model | Moderator level | σ = 0.05 | σ = 0.10 | σ = 0.15 |
| --- | --- | --- | --- | --- |
|  |  | BF10 | BF10 | BF10 |
| Social Connectedness  *Social Measurement | Loneliness Questionnaire | > 100 | > 100 | > 100 |
|  | Loneliness Single Item | > 100 | > 100 | > 100 |
|  | Social Isolation Questionnaire | > 100 | > 100 | > 100 |
|  | Social Isolation Single Item | 0.86 | 0.55 | 0.38 |
|  | Social Support Questionnaire | 17.99 | 13.85 | 7.50 |
|  | Social Support Single Item | 0.78 | 0.53 | 0.36 |

**Supplementary Table 14**

Bayes Factor analysis for social connectedness by trait/state pain interaction.

| Model | Moderator level | σ = 0.05 | σ = 0.10 | σ = 0.15 |
| --- | --- | --- | --- | --- |
|  |  | BF10 | BF10 | BF10 |
| Social Connectedness  *Trait/State Pain | Loneliness State | 1.62 | 2.50 | 1.96 |
|  | Loneliness Trait | > 100 | > 100 | > 100 |
|  | Social Exclusion State | 0.93 | 0.66 | 0.60 |
|  | Social Isolation Trait | > 100 | > 100 | 79.35 |
|  | Social Support State | 5.43 | 6.53 | 4.80 |
|  | Social Support Trait | 39.98 | 16.91 | 11.52 |

**Supplementary Table 15**

Bayes Factor analysis for social connectedness by trait/state social measurement interaction.

| Model | Moderator level | σ = 0.05 | σ = 0.10 | σ = 0.15 |
| --- | --- | --- | --- | --- |
|  |  | BF10 | BF10 | BF10 |
| Social Connectness  *Trait/State Social | Loneliness Trait | > 100 | > 100 | > 100 |
|  | Social Exclusion State | 0.86 | 0.71 | 0.54 |
|  | Social Isolation Trait | > 100 | 99.23 | > 100 |
|  | Social Support State | 4.53 | 8.72 | 9.57 |
|  | Social Support Trait | 15.63 | 23.82 | 11.03 |

**Supplementary Table 16**

Bayes Factor analysis for publication bias tests and continuous moderators.

| Model | Moderator level | σ = 0.05 | σ = 0.1 | σ = 0.15 |
| --- | --- | --- | --- | --- |
|  |  | BF10 | BF10 | BF10 |
| PET | Loneliness | > 100 | 88.23 | > 100 |
|  | Social Support | 0.82 | 0.54 | 0.34 |
|  | Social Isolation | 3.77 | 2.42 | 1.73 |
|  | Social Exclusion | 0.91 | 0.77 | 0.63 |
| Age | Loneliness | 0.06 | 0.03 | 0.01 |
|  | Social Support | 0.10 | 0.05 | 0.03 |
|  | Social Isolation | 0.08 | 0.04 | 0.02 |
|  | Social Exclusion | 0.38 | 0.24 | 0.08 |
| Gender Ratio | Loneliness | 1.13 | 0.79 | 0.72 |
|  | Social Support | 1.87 | 1.81 | 1.56 |
|  | Social Isolation | 1.29 | 1.77 | 1.62 |
|  | Social Exclusion | 1.06 | 0.86 | 0.97 |
| Study Quality | Loneliness | 0.92 | 0.95 | 0.75 |
|  | Social Support | 0.95 | 0.82 | 0.71 |
|  | Social Isolation | 1.12 | 1.17 | 1.12 |
|  | Social Exclusion | 0.98 | 1.02 | 0.94 |

**Supplementary Table 17**

Sensitivity analysis for overall effect across different correlation assumptions.

| Model | Moderator level | ρ = 0.25 | | | ρ = 0.5 | | | ρ = 0.75 | | |
| --- | --- | --- | --- | --- | --- | --- | --- | --- | --- | --- |
|  |  | z | 95% CI LB | 95% CI UB | z | 95% CI LB | 95% CI UB | z | 95% CI LB | 95% CI UB |
| Overall | N/A | -0.09 | -0.11 | -0.07 | -0.09 | -0.11 | -0.07 | -0.09 | -0.11 | -0.07 |

**Supplementary Table 18**

Sensitivity analysis by social connectedness dimension across different correlation assumptions.

| Model | Moderator level | ρ = 0.25 | | | ρ = 0.5 | | | ρ = 0.75 | | |
| --- | --- | --- | --- | --- | --- | --- | --- | --- | --- | --- |
|  |  | z | 95% CI LB | 95% CI UB | z | 95% CI LB | 95% CI UB | z | 95% CI LB | 95% CI UB |
| Social Connected-ness | Loneliness | 0.14 | 0.12 | 0.17 | 0.14 | 0.11 | 0.17 | 0.14 | 0.11 | 0.16 |
|  | Social Exclusion | -0.03 | -0.15 | 0.09 | -0.03 | -0.14 | 0.09 | -0.02 | -0.13 | 0.09 |
|  | Social Isolation | 0.09 | 0.05 | 0.13 | 0.09 | 0.05 | 0.13 | 0.08 | 0.04 | 0.13 |
|  | Social Support | -0.05 | -0.07 | -0.03 | -0.05 | -0.08 | -0.03 | -0.05 | -0.08 | -0.03 |

**Supplementary Table 19**

Sensitivity analysis for social connectedness by pain type interaction across different correlation assumptions.

| Model | Moderator level | ρ = 0.25 | | | ρ = 0.5 | | | ρ = 0.75 | | |
| --- | --- | --- | --- | --- | --- | --- | --- | --- | --- | --- |
|  |  | z | 95% CI LB | 95% CI UB | z | 95% CI LB | 95% CI UB | z | 95% CI LB | 95% CI UB |
| Social Connected-  ness*  Pain Types | Loneliness  Frequency | 0.15 | 0.11 | 0.20 | 0.15 | 0.11 | 0.20 | 0.15 | 0.10 | 0.21 |
|  | Loneliness  Intensity | 0.13 | 0.09 | 0.16 | 0.12 | 0.09 | 0.16 | 0.12 | 0.09 | 0.16 |
|  | Loneliness  Interference | 0.21 | 0.14 | 0.27 | 0.21 | 0.14 | 0.28 | 0.21 | 0.14 | 0.28 |
|  | Social Exclusion  Threshold | -0.10 | -0.34 | 0.13 | -0.10 | -0.34 | 0.13 | -0.10 | -0.33 | 0.13 |
|  | Social Exclusion  Tolerance | -0.08 | -0.35 | 0.19 | -0.08 | -0.35 | 0.18 | -0.08 | -0.34 | 0.19 |
|  | Social Isolation  Intensity | 0.09 | 0.02 | 0.16 | 0.09 | 0.02 | 0.15 | 0.10 | 0.03 | 0.16 |
|  | Social Isolation  Interference | 0.18 | 0.06 | 0.30 | 0.18 | 0.06 | 0.32 | 0.18 | 0.06 | 0.29 |
|  | Social Support  Catastro-  phizing | -0.05 | -0.13 | 0.02 | -0.05 | -0.13 | 0.01 | -0.05 | -0.13 | 0.03 |
|  | Social Support  Disability | -0.05 | -0.12 | 0.01 | -0.05 | -0.12 | 0.01 | -0.05 | -0.12 | 0.01 |
|  | Social Support  Duration | -0.09 | -0.17 | -0.01 | -0.09 | -0.17 | -0.01 | -0.08 | -0.16 | -0.01 |
|  | Social Support  Intensity | -0.05 | -0.08 | -0.02 | -0.05 | -0.08 | -0.02 | -0.05 | -0.08 | -0.02 |
|  | Social Support  Interference | 0.01 | -0.04 | 0.06 | 0.01 | -0.04 | 0.06 | 0.01 | -0.04 | 0.06 |
|  | Social Support  Unpleasant-ness | -0.11 | -0.16 | -0.06 | -0.11 | -0.16 | -0.06 | -0.11 | -0.16 | -0.05 |

**Supplementary Table 20**

Sensitivity analysis for perceived vs. present social support across different correlation assumptions.

| Model | Moderator level | ρ = 0.25 | | | ρ = 0.5 | | | ρ = 0.75 | | |
| --- | --- | --- | --- | --- | --- | --- | --- | --- | --- | --- |
|  |  | z | 95% CI LB | 95% CI UB | z | 95% CI LB | 95% CI UB | z | 95% CI LB | 95% CI UB |
| Social Support  Perceived vs Present | Perceived Social Support | -0.03 | -0.06 | 0.00 | -0.03 | -0.06 | 0.00 | -0.03 | -0.06 | 0.00 |
|  | Present  Social Support | -0.13 | -0.18 | -0.07 | -0.12 | -0.18 | -0.07 | -0.12 | -0.17 | -0.07 |

**Supplementary Table 21**

Sensitivity analysis for social support types across different correlation assumptions.

| Model | Moderator level | ρ = 0.25 | | | ρ = 0.5 | | | ρ = 0.75 | | |
| --- | --- | --- | --- | --- | --- | --- | --- | --- | --- | --- |
|  |  | z | 95% CI LB | 95% CI UB | z | 95% CI LB | 95% CI UB | z | 95% CI LB | 95% CI UB |
| Social Support  Types | Emotional Support | 0.02 | -0.03 | 0.07 | 0.03 | -0.02 | 0.07 | 0.03 | -0.02 | 0.08 |
|  | Instrumental Support | 0.03 | -0.04 | 0.10 | 0.04 | -0.04 | 0.09 | 0.02 | -0.05 | 0.09 |
|  | Familiar Social Presence | -0.14 | -0.22 | -0.06 | -0.15 | -0.23 | -0.06 | -0.15 | -0.24 | -0.07 |
|  | Unfamiliar Social Presence | -0.11 | -0.18 | -0.05 | -0.10 | -0.17 | -0.04 | -0.09 | -0.16 | -0.02 |

**Supplementary Table 22**

Sensitivity analysis for social connectedness by clinical sample interaction across different correlation assumptions.

| Model | Moderator level | ρ = 0.25 | | | ρ = 0.5 | | | ρ = 0.75 | | |
| --- | --- | --- | --- | --- | --- | --- | --- | --- | --- | --- |
|  |  | z | 95% CI LB | 95% CI UB | z | 95% CI LB | 95% CI UB | z | 95% CI LB | 95% CI UB |
| Social Connected-  ness  *Clinical Sample | Loneliness Healthy | 0.20 | 0.13 | 0.27 | 0.20 | 0.13 | 0.27 | 0.20 | 0.13 | 0.27 |
|  | Loneliness Clinical | 0.14 | 0.09 | 0.17 | 0.14 | 0.10 | 0.18 | 0.14 | 0.10 | 0.18 |
|  | Social Exclusion Healthy | -0.03 | -0.18 | 0.13 | -0.02 | -0.17 | 0.13 | -0.02 | -0.16 | 0.13 |
|  | Social Isolation Clinical | 0.12 | 0.06 | 0.17 | 0.12 | 0.07 | 0.18 | 0.13 | 0.07 | 0.19 |
|  | Social Support Healthy | -0.07 | -0.12 | -0.03 | -0.07 | -0.11 | -0.03 | -0.07 | -0.11 | -0.02 |
|  | Social Support Clinical | -0.04 | -0.07 | -0.01 | -0.04 | -0.08 | -0.01 | -0.04 | -0.08 | -0.01 |

**Supplementary Table 23**

Sensitivity analysis for social connectedness by pain classification interaction across different correlation assumptions.

| Model | Moderator level | ρ = 0.25 | | | ρ = 0.5 | | | ρ = 0.75 | | |
| --- | --- | --- | --- | --- | --- | --- | --- | --- | --- | --- |
|  |  | z | 95% CI LB | 95% CI UB | z | 95% CI LB | 95% CI UB | z | 95% CI LB | 95% CI UB |
| Social Connectness  *Pain Classification | Loneliness Secondary Pain | 0.14 | 0.07 | 0.21 | 0.14 | 0.07 | 0.21 | 0.14 | 0.07 | 0.21 |
|  | Social Isolation Secondary Pain | 0.10 | 0.04 | 0.17 | 0.11 | 0.04 | 0.17 | 0.11 | 0.05 | 0.17 |
|  | Social Support Primary Pain | -0.09 | -0.21 | 0.03 | -0.09 | -0.21 | 0.03 | -0.09 | -0.21 | 0.03 |
|  | Social Support Secondary Pain | -0.08 | -0.11 | -0.04 | -0.08 | -0.12 | -0.04 | -0.08 | -0.12 | -0.04 |

**Supplementary Table 24**

Sensitivity analysis for social connectedness by chronicity interaction across different correlation assumptions.

| Model | Moderator level | ρ = 0.25 | | | ρ = 0.5 | | | ρ = 0.75 | | |
| --- | --- | --- | --- | --- | --- | --- | --- | --- | --- | --- |
|  |  | z | 95% CI LB | 95% CI UB | z | 95% CI LB | 95% CI UB | z | 95% CI LB | 95% CI UB |
| Social Connectness  *Chronicity | Loneliness Cross-Sectional | 0.15 | 0.12 | 0.17 | 0.14 | 0.12 | 0.17 | 0.14 | 0.12 | 0.17 |
|  | Loneliness Longitudinal | 0.09 | 0.01 | 0.17 | 0.09 | 0.01 | 0.17 | 0.09 | 0.01 | 0.17 |
|  | Social Isolation Cross-Sectional | 0.09 | 0.05 | 0.13 | 0.09 | 0.05 | 0.13 | 0.09 | 0.04 | 0.13 |
|  | Social Support  Cross-Sectional | -0.05 | -0.07 | -0.02 | -0.05 | -0.07 | -0.02 | -0.05 | -0.07 | -0.02 |
|  | Social Support  Longitudinal | -0.08 | -0.13 | -0.03 | -0.08 | -0.13 | -0.03 | -0.07 | -0.12 | -0.02 |

**Supplementary Table 25**

Sensitivity analysis for social connectedness by study design interaction across different correlation assumptions.

| Model | Moderator level | ρ = 0.25 | | | ρ = 0.5 | | | ρ = 0.75 | | |
| --- | --- | --- | --- | --- | --- | --- | --- | --- | --- | --- |
|  |  | z | 95% CI LB | 95% CI UB | z | 95% CI LB | 95% CI UB | z | 95% CI LB | 95% CI UB |
| Social Connectness  *Study Design | Social Exclusion Between-Person | -0.11 | -0.31 | 0.09 | -0.11 | -0.31 | 0.10 | -0.10 | -0.30 | 0.10 |
|  | Social Support Between-Person | -0.03 | -0.14 | 0.09 | -0.03 | -0.14 | 0.09 | -0.03 | -0.14 | 0.09 |
|  | Social Support Within-Person | -0.11 | -0.17 | -0.06 | -0.11 | -0.17 | -0.06 | -0.11 | -0.17 | -0.06 |

**Supplementary Table 26**

Sensitivity analysis for social connectedness by pain measurement interaction across different correlation assumptions.

| Model | Moderator level | ρ = 0.25 | | | ρ = 0.5 | | | ρ = 0.75 | | |
| --- | --- | --- | --- | --- | --- | --- | --- | --- | --- | --- |
|  |  | z | 95% CI LB | 95% CI UB | z | 95% CI LB | 95% CI UB | z | 95% CI LB | 95% CI UB |
| Social Connectness  *Pain Measurement | Loneliness Questionnaire | 0.20 | 0.15 | 0.25 | 0.20 | 0.14 | 0.25 | 0.19 | 0.14 | 0.25 |
|  | Loneliness Single Item | 0.13 | 0.10 | 0.15 | 0.12 | 0.10 | 0.15 | 0.12 | 0.10 | 0.15 |
|  | Social Exclusion Single Item | -0.07 | -0.21 | 0.08 | -0.06 | -0.21 | 0.08 | -0.06 | -0.20 | 0.08 |
|  | Social Isolation Questionnaire | 0.09 | 0.05 | 0.14 | 0.09 | 0.04 | 0.14 | 0.09 | 0.04 | 0.14 |
|  | Social Isolation Single Item | 0.07 | 0.02 | 0.12 | 0.07 | 0.02 | 0.12 | 0.07 | 0.02 | 0.12 |
|  | Social Support Questionnaire | -0.02 | -0.05 | 0.02 | -0.02 | -0.05 | 0.02 | -0.02 | -0.05 | 0.02 |
|  | Social Support Single Item | -0.08 | -0.11 | -0.05 | -0.08 | -0.11 | -0.05 | -0.08 | -0.11 | -0.05 |

**Supplementary Table 27**

Sensitivity analysis for social connectedness by social measurement interaction across different correlation assumptions.

| Model | Moderator level | ρ = 0.25 | | | ρ = 0.5 | | | ρ = 0.75 | | |
| --- | --- | --- | --- | --- | --- | --- | --- | --- | --- | --- |
|  |  | z | 95% CI LB | 95% CI UB | z | 95% CI LB | 95% CI UB | z | 95% CI LB | 95% CI UB |
| Social Connectness  *Social Measurement | Loneliness Questionnaire | 0.14 | 0.11 | 0.18 | 0.14 | 0.11 | 0.18 | 0.14 | 0.10 | 0.18 |
|  | Loneliness Single Item | 0.14 | 0.10 | 0.18 | 0.14 | 0.10 | 0.17 | 0.14 | 0.10 | 0.17 |
|  | Social Isolation Questionnaire | 0.12 | 0.07 | 0.17 | 0.12 | 0.07 | 0.18 | 0.12 | 0.07 | 0.18 |
|  | Social Isolation Single Item | 0.01 | -0.05 | 0.06 | 0.01 | -0.05 | 0.06 | 0.01 | -0.05 | 0.06 |
|  | Social Support Questionnaire | -0.05 | -0.07 | -0.02 | -0.05 | -0.07 | -0.02 | -0.05 | -0.07 | -0.02 |
|  | Social Support Single Item | -0.04 | -0.11 | 0.04 | -0.03 | -0.11 | 0.04 | -0.03 | -0.10 | 0.04 |

**Supplementary Table 28**

Sensitivity analysis for social connectedness by trait/state pain interaction across different correlation assumptions.

| Model | Moderator level | ρ = 0.25 | | | ρ = 0.5 | | | ρ = 0.75 | | |
| --- | --- | --- | --- | --- | --- | --- | --- | --- | --- | --- |
|  |  | z | 95% CI LB | 95% CI UB | z | 95% CI LB | 95% CI UB | z | 95% CI LB | 95% CI UB |
| Social Connectness  *Trait/State Pain | Loneliness State | 0.14 | 0.07 | 0.20 | 0.13 | 0.07 | 0.20 | 0.13 | 0.06 | 0.20 |
|  | Loneliness Trait | 0.15 | 0.12 | 0.18 | 0.15 | 0.12 | 0.17 | 0.14 | 0.12 | 0.17 |
|  | Social Exclusion State | -0.06 | -0.23 | 0.11 | -0.05 | -0.22 | 0.11 | -0.05 | -0.21 | 0.12 |
|  | Social Isolation Trait | 0.09 | 0.05 | 0.13 | 0.09 | 0.05 | 0.13 | 0.09 | 0.05 | 0.13 |
|  | Social Support State | -0.10 | -0.14 | -0.05 | -0.09 | -0.14 | -0.05 | -0.09 | -0.13 | -0.04 |
|  | Social Support Trait | -0.05 | -0.07 | -0.02 | -0.05 | -0.07 | -0.02 | -0.05 | -0.07 | -0.02 |

**Supplementary Table 29**

Sensitivity analysis for social connectedness by trait/state social measurement interaction across different correlation assumptions.

| Model | Moderator level | ρ = 0.25 | | | ρ = 0.5 | | | ρ = 0.75 | | |
| --- | --- | --- | --- | --- | --- | --- | --- | --- | --- | --- |
|  |  | z | 95% CI LB | 95% CI UB | z | 95% CI LB | 95% CI UB | z | 95% CI LB | 95% CI UB |
| Social Connectness*  Trait/State Social | Loneliness Trait | 0.14 | 0.12 | 0.17 | 0.14 | 0.11 | 0.17 | 0.14 | 0.11 | 0.17 |
|  | Social Exclusion State | -0.05 | -0.22 | 0.11 | -0.05 | -0.21 | 0.11 | -0.05 | -0.21 | 0.12 |
|  | Social Isolation Trait | 0.09 | 0.05 | 0.13 | 0.09 | 0.05 | 0.13 | 0.09 | 0.05 | 0.13 |
|  | Social Support State | -0.10 | -0.15 | -0.06 | -0.10 | -0.14 | -0.05 | -0.09 | -0.14 | -0.05 |
|  | Social Support Trait | -0.05 | -0.07 | -0.02 | -0.05 | -0.07 | -0.02 | -0.05 | -0.07 | -0.02 |

**Supplementary Table 30**

Sensitivity analysis for publication bias tests and continuous moderators across different correlation assumptions.

| Model | Moderator level | ρ = 0.25 | | | ρ = 0.5 | | | ρ = 0.75 | | |
| --- | --- | --- | --- | --- | --- | --- | --- | --- | --- | --- |
|  |  | B | 95% CI LB | 95% CI UB | B | 95% CI LB | 95% CI UB | B | 95% CI LB | 95% CI UB |
| PET | Loneliness | 1.63 | 0.94 | 2.32 | 1.64 | 0.96 | 2.33 | 1.66 | 0.97 | 2.34 |
|  | Social Support | -0.30 | -0.72 | 0.13 | -0.30 | -0.73 | 0.12 | -0.31 | -0.74 | 0.12 |
|  | Social Isolation | 1.31 | 0.51 | 2.11 | 1.30 | 0.50 | 2.10 | 1.30 | 0.50 | 2.10 |
|  | Social Exclusion | -0.76 | -1.91 | 0.40 | -0.73 | -1.88 | 0.41 | -0.69 | -1.82 | 0.44 |
| Age | Loneliness | -0.00 | -0.00 | 0.00 | -0.00 | -0.00 | 0.00 | -0.00 | 0.00 | 0.00 |
|  | Social Support | 0.00 | 0.00 | 0.00 | 0.00 | -0.00 | 0.00 | 0.00 | -0.00 | 0.00 |
|  | Social Isolation | -0.00 | -0.01 | 0.00 | -0.00 | -0.01 | 0.00 | -0.00 | -0.01 | 0.00 |
|  | Social Exclusion | 0.00 | -0.03 | 0.03 | -0.00 | -0.03 | 0.03 | 0.00 | -0.03 | 0.03 |
| Gender Ratio | Loneliness | 0.05 | -0.07 | 0.19 | 0.05 | -0.08 | 0.18 | 0.05 | -0.07 | 0.18 |
|  | Social Support | -0.07 | -0.17 | 0.04 | -0.07 | -0.17 | 0.03 | -0.07 | -0.17 | 0.03 |
|  | Social Isolation | 0.15 | 0.02 | 0.27 | 0.15 | 0.02 | 0.27 | 0.15 | 0.02 | 0.28 |
|  | Social Exclusion | 0.15 | -0.48 | 0.78 | 0.15 | -0.48 | 0.78 | 0.15 | -0.47 | 0.77 |
| Study Quality | Loneliness | 0.09 | -0.09 | 0.27 | 0.09 | -0.09 | 0.27 | 0.09 | -0.09 | 0.26 |
|  | Social Support | -0.07 | -0.24 | 0.09 | -0.07 | -0.23 | 0.10 | -0.06 | -0.23 | 0.10 |
|  | Social Isolation | 0.20 | -0.08 | 0.49 | 0.21 | -0.08 | 0.49 | 0.21 | -0.08 | 0.49 |
|  | Social Exclusion | -0.11 | -0.85 | 0.62 | -0.11 | -0.84 | 0.62 | -0.10 | -0.82 | 0.62 |

**Supplementary Table 31**

Heterogeneity estimates for all models.

| Model | Significance | Heterogeneity  estimate | Study  heterogeneity (σ²) | Cohort  heterogeneity (σ²) | Effect  Heterogeneity (σ²) |
| --- | --- | --- | --- | --- | --- |
| Overall | No moderator included | Q(519) = 32706.21, p < .001 | 0.011 | 0.003 | 0.008 |
| Social Connectedness | F(4,280) = 39.50, p < .001 | Q(516) = 15118.66, p < .001 | 0.01 | 0.002 | 0.007 |
| Social Connectedness  *Pain Types | F(13,179) = 9.52, p < .001 | Q(333) = 2245.43, p < .001 | 0.01 | 0.006 | 0.006 |
| Social Support  Perceived vs Present | F(2,122) = 8.75, p < .001 | Q(220) = 2454.78, p < .001 | 0.01 | 0.00 | 0.008 |
| Social Support  Types | F(4,62) = 5.97, p < .001 | Q(123) = 503.66, p < .001 | 0.006 | 0.003 | 0.006 |
| Social Connectedness  *Clinical Sample | F(6,226) = 19.86, p < .001 | Q(415) = 9278.57, p < .001 | 0.016 | 0.00 | 0.008 |
| Social Connectedness  *Pain Classification | F(4,84) = 14.84, p < .001 | Q(148) = 5234.58, p < .001 | 0.008 | 0.00 | 0.01 |
| Social Connectedness  *Association Type | F(5,211) = 26.33, p < .001 | Q(388) = 14586.69, p < .001 | 0.009 | 0.00 | 0.007 |
| Social Connectedness  *Study Design | F(3,52) = 0.97, p = .002 | Q(89) = 285.41, p < .001 | 0.027 | 0.035 | 0.006 |
| Social Connectedness  *Pain Measurement | F(7,252) = 22.50, p < .001 | Q(473) = 11350.39, p < .001 | 0.008 | 0.003 | 0.007 |
| Social Connectedness  *Social Measurement | F(6,218) = 20.22, p < .001 | Q(410) = 10080.18, p < .001 | 0.008 | 0.002 | 0.008 |
| Social Connectness  *Trait/State Pain | F(5,272) = 26.11, p < .001 | Q(504) = 14977.67, p < .001 | 0.01 | 0.003 | 0.007 |
| Social Connectness  *Trait/State Social | F(4,274) = 31.91, p < .001 | Q(504) = 14986.96, p < .001 | 0.01 | 0.003 | 0.007 |

**Supplementary Results**

***Bias Assessment***

To test the robustness of the associations between dimensions of social connectedness and pain, we visually inspected funnel plots and performed PET-PEESE analysis to test for and adjust small study bias. For loneliness (F(1,90) = 22.56, p < .001, BF10 = 93.72, Supplementary Figure 3) and social isolation (F(1,39) = 10.81, p = .002, BF10 = 2.55, Supplementary Figure 5), smaller studies tended to report larger effects, indicating the presence of publication bias. After bias adjustment, the association between loneliness and pain (z = 0.10 [0.07;0.12], t(90) = 7.64, p < .001, BF10 > 100), as well as social isolation and pain (z = 0.07 [0.03;0.13], t(39) = 3.26, p = .002, BF10 = 1.15) remained significant, indicating robustness despite bias. No evidence of small-study bias was observed for social support or social exclusion (Supplementary Figures 4 and 6). Study quality was examined as an additional source of bias. Study quality did not moderate the observed associations (Supplementary Figures 7-10). Studies using covariates in their analyses did not show any different effects compared to studies without covariate use.

***Moderator analyses of methodological factors***

In a final set of moderator analyses, we examined methodological factors, specifically construct assessment and experimental design. For construct assessment, we compared single-item measures with multi-item instruments (typically questionnaires) for both pain and social connectedness. For pain assessment, loneliness showed positive associations with pain irrespective of measurement approach (multiple items: z = 0.20 [0.14;0.25], t(252) = 7.40, p < .001, BF10 > 100; single item: z = 0.12 [0.10;0.15], t(252) = 9.91, p < .001, BF10 > 100; Supplementary Figures 21A and 22A), with stronger associations observed for multi-item measures (t(252) = 2.86, p = .005, BF10 = 9.83). In contrast, social support was negatively associated with pain only when assessed using single-item measures (z = -0.08 [-0.11;-0.05], t(252) = 5.77, p < .001, BF10 > 100), whereas questionnaire-based measures showed no significant association (z = -0.02 [-0.05;0.02], t(252) =1.00, p = .319, BF10 = 0.23; Supplementary Figures 21B and 22B). Social isolation exhibited positive associations with pain for both multi-item (z = 0.09 [0.04;0.14], t(252) = 3.68, p < .001, BF10 = 10.67) and single-item assessments (z = 0.07 [0.02;0.12], t(252) = 2.89, p = .004, BF10 = 9.77; Supplementary Figures 21C and 22C). When examining the assessment of social connectedness, loneliness was positively associated with pain regardless of whether multi-item (z = 0.14 [0.11;0.18], t(218) = 7.78, p < .001, BF10 > 100) or single-item measures (z = 0.14 [0.10;0.17], t(218) = 7.84, p < .001, BF10 > 100; Supplementary Figure 23A and 24A) were used. In contrast, significant associations for social isolation and social support were observed only when these constructs were assessed using multi-item instruments (social isolation: z = 0.12 [0.07;0.18], t(218) = 4.60, p < .001, BF10 > 100; social support: z = -0.05 [-0.07;-0.02], t(218) = 3.23, p = .001, BF10 = 13.34); single-item measures yielded no reliable associations (social isolation: z = 0.01 [-0.05;0.06], t(218) = 0.25, p = .801, BF10 = 0.57; social support: z = -0.03 [-0.11;0.04], t(218) = 0.93, p = .354, BF10 = 0.57; Supplementary Figure 23B/C and 24B/C). Finally, we examined experimental design effects for social support by comparing within-participant and between-participant studies. A significant negative association between social support and pain was observed in within-participant designs (z = -0.11 [-0.17;-0.06], t(52) = 4.02, p < .001, BF10 = 44.84), whereas no association was detected in between-participant designs (z = -0.03 [-0.14;0.09], t(52) = 0.48, p = .635, BF10 = 0.69; Supplementary Figures 25 and 26), indicating greater sensitivity of within-participant paradigms.
